# Machine learning-optimized perinatal depression screening: Maximum impact, minimal burden

**DOI:** 10.1101/2025.10.13.25337771

**Authors:** Eric G. Hurwitz, Caroline Shell, Kritika Chugh, Veerle Bergink, Rena C. Patel, Melissa A. Haendel, N3C consortium

**Affiliations:** Department of Genetics, University of North Carolina at Chapel Hill, 120 Mason Farm Road, Chapel Hill, NC, 27599, United States; Department of OBGYN, Sinai Hospital of Baltimore, 2435 W. Belvedere Ave. Suite 33 Baltimore, MD, 21215, United States; Department of Psychiatry, University of North Carolina at Chapel Hill, 101 Manning Dr #1, Chapel Hill, NC, 27514, United States; Department of Psychiatry, Icahn School of Medicine at Mount Sinai, One Gustave L. Levy Place, New York, NY, 10029, United States; Department of Medicine, University of Alabama at Birmingham, 510 20th Street South Birmingham, AL, 35294, United States

**Author notes:** **Corresponding author:** Eric Hurwitz, 120 Mason Farm Road, Chapel Hill, NC, 27599. email addresses: Caroline Shell, Kritika Chugh, Veerle Bergink, Rena C. Patel, Melissa Haendel).

**Keywords:** perinatal depression, machine learning, Edinburgh Postnatal Depression Scale, screening, maternal mental health

## Abstract

**Background:** Perinatal depression affects up to 30% of pregnant and postpartum women and represents a leading cause of maternal morbidity and mortality. Despite clinical guidelines recommending routine screening with tools like the Edinburgh Postnatal Depression Scale (EPDS), implementation rates remain less than 20% in clinical practice. Assessment burden and administration time are key barriers to widespread screening adoption. Current methods to abbreviate screening instruments rely on traditional psychometric approaches that may not optimize predictive accuracy. Machine learning methods offer potential to develop briefer screening tools while maintaining clinical performance.

**Objective:** To develop and validate a machine learning framework for abbreviating the EPDS from 10 to 2 questions and evaluate its clinical utility compared to traditional additive scoring methods.

**Study Design:** Cross-sectional, retrospective, observational study using electronic health record data (01/2018-10/2024) with external validation in an independent prospective cohort (01/2017-01/2020). We evaluated all possible 2-question through 5-question EPDS combinations using linear regression to predict remaining scores from question subsets, which combined with the known subset scores reconstructed full 10-item EPDS total scores. Binary classification models assessed screening accuracy for positive depression screens using the clinical threshold of 13. Decision curve analysis evaluated net benefit across threshold probabilities. For methodological validation, we applied the framework to PHQ-9 data. Study setting included the National Clinical Cohort Collaborative (N3C), a multi-site electronic health record database of clinical sites across the United States, and Washington University in St. Louis longitudinal prospective pregnancy cohort. Participants included women with complete item-level EPDS assessments in N3C (n=22,924 total; 7,750 postpartum-specific subset), external validation cohort of pregnant women (n=1,217), and individuals with PHQ-9 data for methodological validation (n=398,606). Statistical analyses included coefficient of determination (R^2^), root mean square error, mean absolute error, sensitivity, specificity, F1 score, and decision curve analysis.

**Results:** Among 22,924 women (median age 32 years, 48.8% White non-Hispanic [n=11,190], 11.5% positive screens [n=2,626]), optimal 2-question combinations were Q4:anxious plus Q8:sad and Q5:scared plus Q8:sad, both achieving R^2^=0.70 (root mean square error=2.17-2.23). Binary classification demonstrated sensitivity 68-72% and specificity 98-99% for identifying positive screens. Performance remained consistent in postpartum subset (R^2^=0.70) and external pregnancy cohort (R^2^=0.64). Decision curve analysis showed machine learning approaches provided superior net benefit (0.01-0.03) compared to traditional additive scoring across clinically relevant threshold probabilities. Methodological validation using PHQ-9 confirmed framework generalizability (R^2^=0.63). Adding demographic or clinical covariates did not improve predictive performance.

**Conclusion:** A machine learning framework identified 2-question EPDS subsets that maintain predictive accuracy while reducing assessment burden by 80%. This approach demonstrates superior net benefit compared to traditional abbreviated scoring methods and could enhance perinatal depression screening implementation in resource-constrained settings, obstetric telehealth, and routine clinical workflows. With ∼3.6 million annual births in the United States, widespread adoption could substantially improve identification of women with perinatal depression.

**Condensation page:** *Tweetable statement:* Machine learning identifies two EPDS questions that maintain screening accuracy while reducing assessment burden by 80%, potentially improving perinatal depression detection across diverse clinical settings.

**AJOG at a glance:**

a. Why was this study conducted?

- Perinatal depression affects up to 30% of women but screening implementation remains below 20% despite clinical guidelines
- Assessment burden is a key barrier to widespread screening adoption
- Current abbreviation methods rely on traditional psychometric approaches rather than machine learning
b. What are the key findings?

- Machine learning identified two-question Edinburgh Postnatal Depression Scale combinations that maintained predictive accuracy (R^2^=0.70) while reducing assessment burden by 80%
- Decision curve analysis showed superior net benefit versus traditional additive scoring
- Model performance remained strong across validation cohorts
c. What does this study add to what is already known?

- First application of machine learning to Edinburgh Postnatal Depression Scale abbreviation, with the largest sample size to date
- Machine learning-weighted scoring outperforms traditional scoring for shortened screening tools
- Provides a generalizable machine learning framework to shorten other mental health assessments

## Introduction

Standardized mental health assessments are essential screening tools, but longer assessments have lower response rates and quality^1–3^. In perinatal care specifically, provider time constraints and patient burden are among the most commonly cited barriers to screening, contributing to low screening rates despite guideline recommendations. Abbreviated versions like the PHQ-2 (a 2-item subset of the PHQ-9) are widely used to reduce burden while maintaining validity^4^. Development of such tools typically relies on psychometric methods including item response theory, computerized adaptive testing, or classical test theory^5–9^.

The Edinburgh Postnatal Depression Scale (EPDS) is the most commonly used assessment for screening perinatal mental health disorders, one of the most frequent complications during pregnancy and postpartum^10^. This 10-item self-administered questionnaire uses responses ranging 0-3 per question, with total scores ≥13 indicating positive screening for perinatal depression^11^. Untreated perinatal mental health disorders contribute significantly to maternal morbidity and mortality, with suicide representing a leading cause of pregnancy-related death^12^. Perinatal depression is diagnosed in approximately 9% of women, yet symptom-based screening identifies elevated depressive symptoms in up to 30% during the COVID-19 pandemic. This discrepancy reflects both the burden of the condition and the urgent need for more precise screening approaches^13,14^.

Despite guidelines from the American College of Obstetricians and Gynecologists and American Academy of Pediatrics recommending screening at multiple pregnancy and postpartum timepoints, screening rates remain below 20%^15–17^.

This gap highlights the need for efficient screening tools that minimize burden while maintaining accuracy. Previous efforts to shorten the EPDS using various statistical approaches have yielded subsets ranging from 2 to 8 items^18–23^. Harel et al. applied item response theory to create a 5-item version demonstrating sensitivity of 0.83 and specificity of 0.86^24^. However, these studies have key limitations: small to moderate sample sizes, limited focus on 2-item subsets despite their clinical utility, and use of pre-pandemic data that may not reflect current symptom patterns^18–24^.

We developed a generalizable machine learning (ML) framework to identify optimal abbreviated subsets of the EPDS for perinatal depression screening, using data from the National Clinical Cohort Collaborative (N3C), a large electronic health record dataset^25^. This study aimed to: 1) identify the optimal EPDS question subsets that best predict full-scale scores; 2) evaluate the net benefit of ML-weighted scoring against traditional additive scoring approaches; 3) validate findings across multiple cohorts and time periods to assess generalizability and temporal stability; and 4) confirm methodological generalizability by applying the same framework to the PHQ-9. This approach offers significant potential to reduce perinatal mental health screening burden while improving detection rates across clinical and research settings.

## Materials and methods

### Study population

The National Clinical Cohort Collaborative (N3C) contains harmonized electronic health record data from clinical sites across the United States from January 2018 onwards with weekly updates. Data in this study from the Enclave (a secure, centralized analytic computing environment) uses release up to December 16, 2024 (v187 with 85 data partner sites) using a Limited Data Set (de-identified health records except for dates and geographic information at the area-level, in accordance with HIPAA privacy standards). The NIH IRB approved N3C, and this project (RP-E39D65) was approved by the N3C Data Access Committee. UNC study access was approved by the IRB (21-0309).

The temporal scope of N3C data captures both pre-pandemic baseline periods and substantial shifts in perinatal mental health following COVID-19 onset in March 2020. Studies have documented increased symptom severity and greater reliance on screening tools during this period, underscoring the need for brief, efficient screening instruments^26–28^.

### EPDS and study cohort

Item-level EPDS responses were obtained from the N3C Observation table using standardized OMOP concepts (Table S-I). Each EPDS question was assigned a symptom label, consistent with that by Harel et al. (e.g., Q1=funny, Q4=anxiety); PHQ-9 symptoms were labeled according to DSM-5 criteria for major depressive disorder (Table S-I)^24,29^. We included individuals with female sex at birth who completed all 10 EPDS items on the same date. Total scores were calculated by summing item-level responses. Given the known skew towards lower scores, when multiple assessments existed for the same individual, we selected the assessment with the highest total score as one approach to partially mitigate this imbalance and improve representation of individuals with elevated symptom scores (i.e., only 1 row of EPDS data per participant in the analytic dataset).

### ML method for assessment abbreviation

We developed a ML approach to identify which subset of EPDS questions could predict the total score. Each model applied linear regression using 2-question responses to predict the remaining score, which could be added to the known 2-question responses for a predicted total EPDS score (Figure S1). Model performance for the question-combination search was estimated using repeated 10-fold cross-validation (10 folds, 3 repeats) rather than a separate train/test split. This was repeated for all 2-, 3-, 4-, and 5-question combinations to characterize how predictive performance scales with subset length. We focused primarily on 2-question combinations for clinical implementation, as evidence demonstrates that longer assessments are associated with lower response rates and completion, and established ultra-brief screeners such as the PHQ-2 demonstrate that 2-item subsets can minimize burden while maintaining clinical validity^4^. Full results for 3-, 4-, and 5-question combinations are reported to contextualize the performance tradeoff between brevity and predictive accuracy.

Performance was assessed using R^2^, root mean square error (RMSE), and mean absolute error (MAE)^30^. We then extended this approach to binary classification using logistic regression with the clinical cutoff of ≥13 for positive screening, which is consistent with prior work^11,31^. Binary classification models were evaluated using 10-fold cross-validation; cross-validated fold-level performance was compared against full-sample performance to assess overfitting. Model performance metrics included accuracy (95% confidence interval [CI]), kappa, sensitivity, specificity, positive predictive value, negative predictive value, prevalence, detection rate, detection prevalence, balanced accuracy, precision, recall, and F1 score^32^. Of note, linear/logistic regression was deliberately chosen to prioritize a simpler and more interpretable ML framework as opposed to other more complex algorithms, such as random forest or XGBoost.

### Validation cohorts

#### Postpartum cohort

To evaluate performance in postpartum women specifically, we identified women who completed the EPDS 4 weeks to 12 months postpartum using the Hierarchy and rule-based pregnancy episode Inference integrated with Pregnancy Progression Signatures (HIPPS) algorithm developed in N3C^33^. We excluded assessments before 4 weeks to avoid confounding by postpartum blues, which affect up to 85% of new mothers^34^.

#### External validation cohort

For external validation, we used data from 1,217 pregnant women enrolled in a prospective study conducted by Washington University in St. Louis (2017-2020)^35,36^. This cohort differed from N3C in its prospective design with active longitudinal follow-up and was collected entirely pre-pandemic, allowing assessment of temporal generalizability.

### Methodological validation using PHQ-9

To confirm our framework’s generalizability beyond the EPDS, we applied the same approach to the PHQ-9, a 9-item depression screening tool^1^. Item-level PHQ-9 responses were obtained from N3C using standardized concepts (Table S-I), and we identified optimal 2-question combinations using the same methods. The PHQ-9 provided methodological validation because: 1) large sample size enabled robust testing, and 2) the established PHQ-2 abbreviation provided a benchmark comparison^4^.

### Comparison of ML and simple addition approaches

We compared our ML approach against simple additive scoring, which sums responses to two questions (each rated 0–3, total range 0–6) and classifies scores ≥3 as positive, analogous to the PHQ-2 scoring convention. We evaluated four strategies: 1) simple addition with ≥3 threshold, 2) ML classification, 3) ensemble OR logic (positive if either method positive), and 4) ensemble AND logic (positive if both methods positive).

Performance was compared using F1 score, kappa, precision, and recall.

### Decision curve analysis

We conducted decision curve analysis to evaluate the net benefit of each screening strategy across a range of clinical scenarios. In this framework, the ‘threshold probability’ represents the minimum probability of depression at which a clinician would act (for example, refer a patient for further evaluation). A clinician in a well-resourced setting might act at a low threshold (referring patients even with modest risk), while one in a resource-constrained setting might require a higher threshold before referring. Net benefit was calculated by weighing the benefit of correctly identifying true positive screens against the harm of unnecessary referrals from false positives, across threshold probabilities from 0.01 to 0.99. Each screening strategy was compared against two reference strategies: ‘screen all’ (refer every patient regardless of score) and ‘screen none’ (refer no patients). A strategy was considered clinically useful if its net benefit exceeded both reference strategies, meaning it outperformed both blanket referral and no screening across a given range of thresholds^37,38^.

### Statistical analysis

Demographic comparisons between cohorts used Chi-square tests for categorical variables and unpaired t-tests for continuous variables^39^. Linear regression assessed relationships between individual question responses and total scores, with Bonferroni correction for multiple comparisons^40^. All analyses used R with the Caret package for ML^41^.

## Results

### Demographics of EPDS study participants

After filtering for individuals with complete item-level EPDS responses on the same date who were female sex at birth, the N3C cohort included 22,924 women (143 pre-COVID [0.6%], 22,781 post-COVID [99.4%]), of whom 2,626 (11.5%) screened positive (total score ≥13). The majority were White non-Hispanic (11,190 [48.8%]), with higher representation of Black non-Hispanic women among positive screens (708 [27.0%]) compared to negative screens (4,443 [21.9%]). Median age was 32 years (IQR 27-37) (Table I).

**Table 1:**
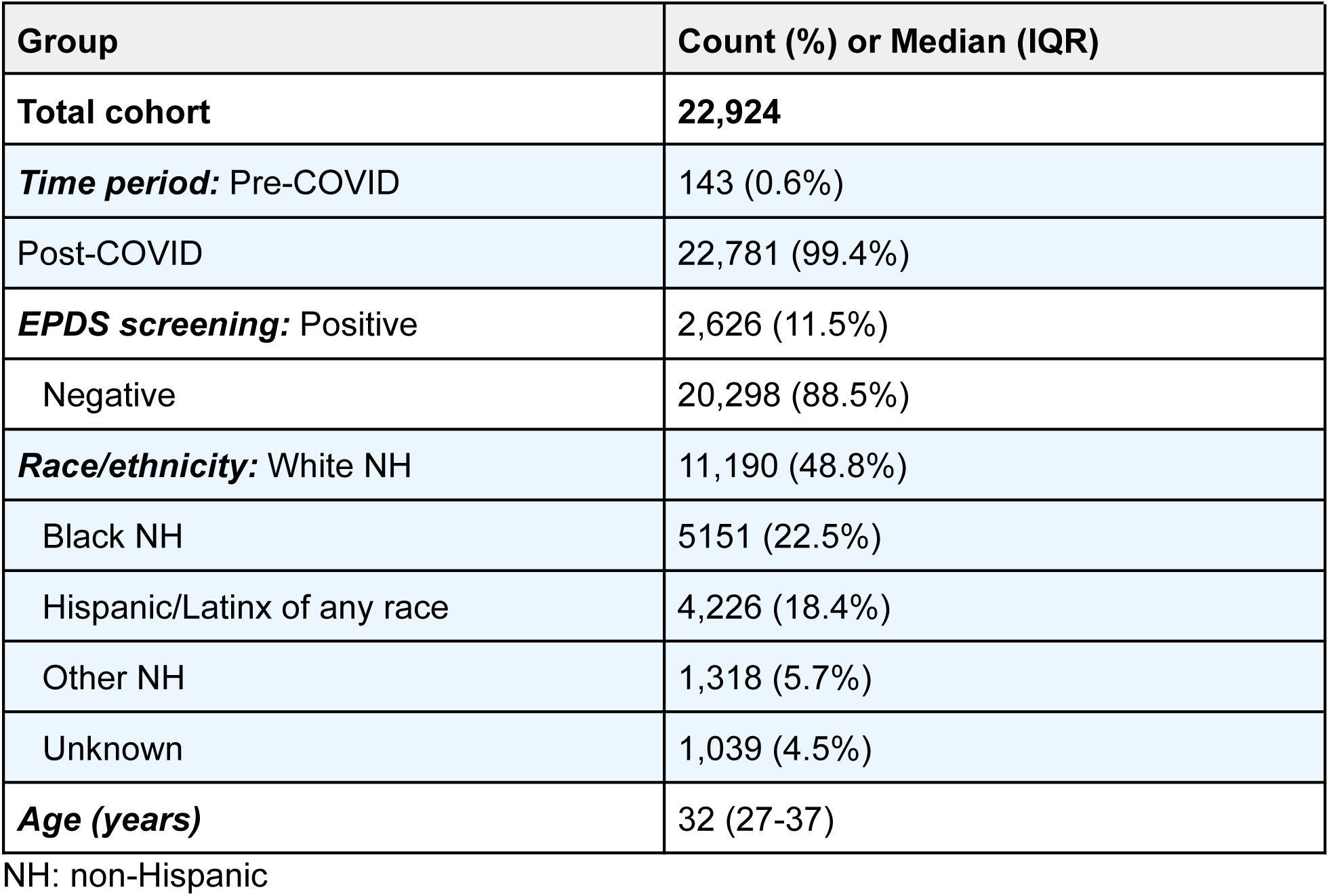
Demographics of the N3C cohort with any EPDS data.

| Group | Count (%) or Median (IQR) |
| --- | --- |
| **Total cohort** | **22,924** |
| ***Time period:*** Pre-COVID | 143 (0.6%) |
| Post-COVID | 22,781 (99.4%) |
| ***EPDS screening:*** Positive | 2,626 (11.5%) |
| Negative | 20,298 (88.5%) |
| ***Race/ethnicity:*** White NH | 11,190 (48.8%) |
| Black NH | 5151 (22.5%) |
| Hispanic/Latinx of any race | 4,226 (18.4%) |
| Other NH | 1,318 (5.7%) |
| Unknown | 1,039 (4.5%) |
| ***Age (years)*** | 32 (27-37) |

NH: non-Hispanic

### We developed a generalizable ML method to shorten psychometric assessments applied to the EPDS

We conceptualized shortening the EPDS by predicting total scores from question subsets rather than applying new cutoff scores. Individual EPDS items showed strong associations with total scores, with more severe responses corresponding to higher totals (*P*<0.001; Figure S3). We developed linear regression models testing all possible 2-question combinations to predict total scores. Q4:anxious + Q8:sad demonstrated highest performance (R^2^=0.70, RMSE=2.17, MAE=1.61), followed closely by Q5:scared + Q8:sad (R^2^=0.70, RMSE=2.23, MAE=1.72) (Figure 1). Performance varied substantially across question pairs (lowest R^2^=0.37; Table S-III), confirming top combinations were identified based on predictive value. Binary classification using the clinical threshold ≥13 demonstrated high performance for both pairs (Table S-V).

**Figure 1:**
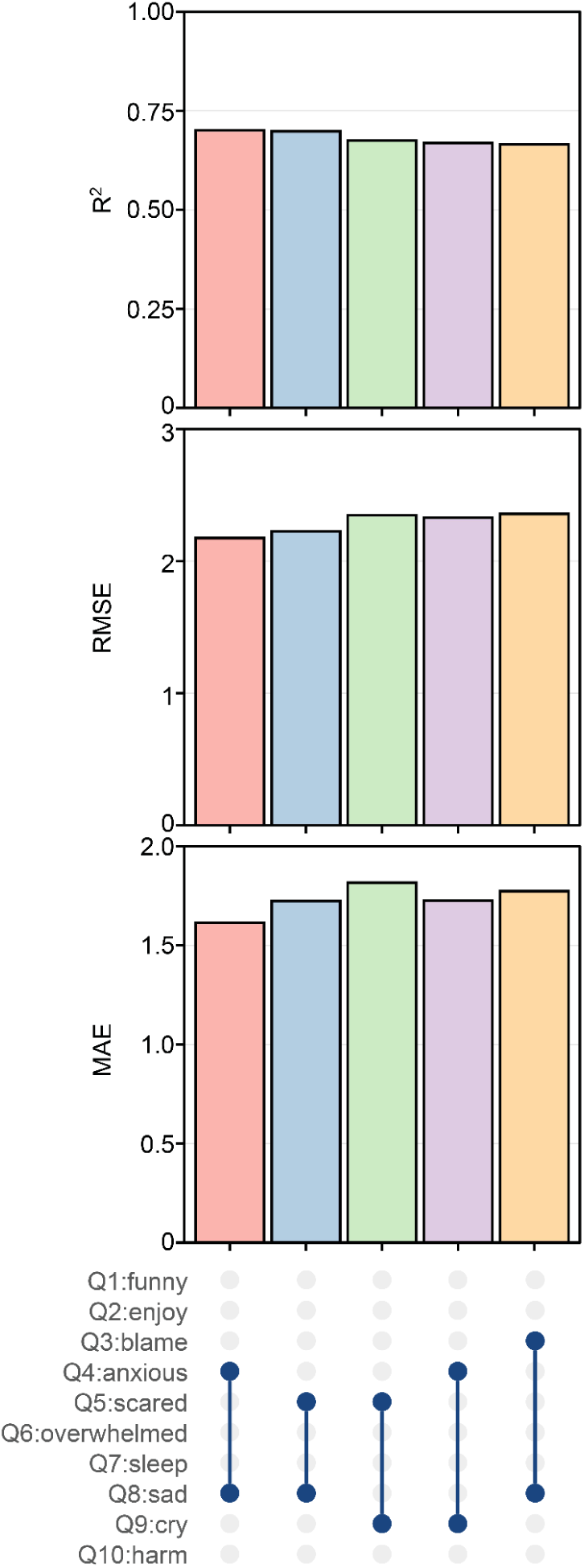
Q4:anxious + Q8:sad and Q5:scared + Q8:sad models displayed the highest performance for predicting the remaining EPDS total score. Model performance metrics across the top five different models (represented by colored bars) using 2-item EPDS question subsets to predict the EPDS total score from the entire cohort of women who took the EPDS in N3C. The top panel displays R^2^ values (coefficient of determination), the middle panel shows RMSE, while the bottom panel shows MAE. The dot-and-line diagram at the bottom indicates which variables (Q1-Q10) were included in each model configuration, with blue dots representing included variables connected by vertical lines.

### Our method shortening the EPDS remained stable for women in the postpartum period and an external pregnancy cohort

We validated models in two cohorts: a postpartum-specific N3C subset (n=7,750) and an independent external cohort of pregnant women from Washington University (n=1,217)^35,36^. The N3C postpartum cohort showed slightly higher positive screening (13.6% vs 11.5%) but similar demographics (Table S-VI). The Wash U cohort was demographically distinct, with higher Black non-Hispanic representation (55.2% vs 22.5%, *P*<0.001), younger median age (28 vs 32 years, *P*<0.001), and lower positive screening (7.6% vs 11.5%, *P*<0.001) (Table S-VI). Both validation cohorts demonstrated consistent performance of top question pairs despite demographic differences and varying COVID-19 exposure periods. Q4:anxious + Q8:sad and Q5:scared + Q8:sad maintained high predictive accuracy across all cohorts with minimal performance differences, confirming robustness across diverse populations (Figure 2).

**Figure 2:**
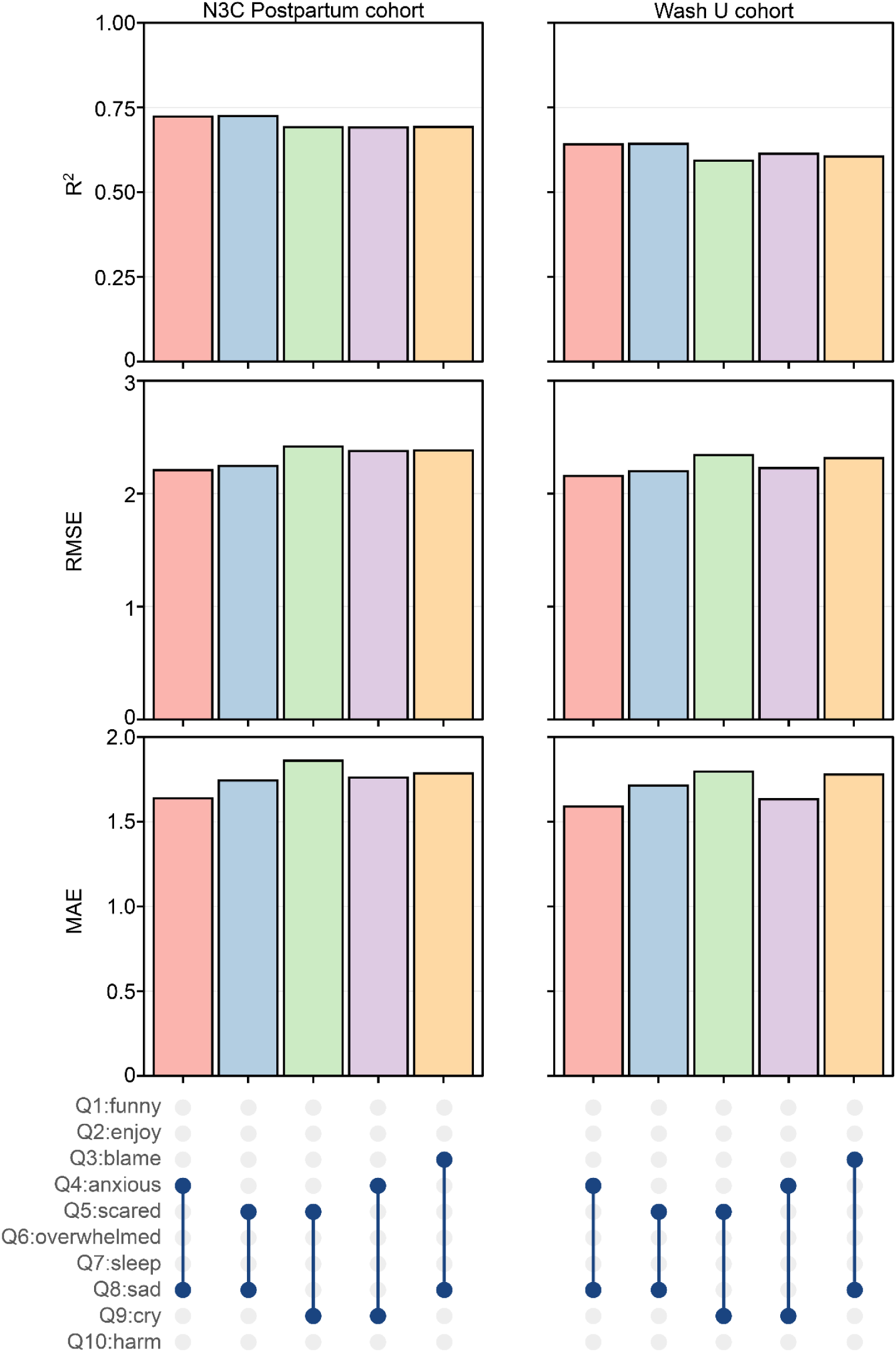
Q4:anxious + Q8:sad and Q5:scared + Q8:sad models displayed the highest performance for predicting the remaining EPDS total score in a postpartum cohort. Model performance metrics across the top five different models (represented by colored bars) using 2-item EPDS question subsets to predict the EPDS total score among women in the postpartum period in N3C (left) and Wash U cohort (right). The top panel displays R^2^ values (coefficient of determination), the middle panel shows RMSE, while the bottom panel shows MAE. The dot-and-line diagram at the bottom indicates which variables (Q1-Q10) were included in each model configuration, with blue dots representing included variables connected by vertical lines.

### Our ML method generalized to an assessment beyond the EPDS

To confirm framework generalizability, we applied it to the PHQ-9, a 9-item depression screening tool with an established 2-question abbreviation (PHQ-2)^4^. In 398,606 individuals (37.9% screening, positive), individual question responses showed similar dose-response relationships with total scores (Table S-VII and Figure S5). Our method identified Q2:sad + Q7:concentration as optimal, differing from the established PHQ-2 (Q1:enjoy + Q2:sad) (Figure S6). Performance varied substantially across question pairs (lowest R^2^=0.40; Table S-VIII), confirming top combinations reflected specific predictive value. Binary classification (threshold ≥10) showed comparable performance between approaches, with slightly better results for our ML-identified combination (Table S-IX).

### Model performance improved with additional questions included

To evaluate the trade-off between reducing assessment burden and maintaining predictive accuracy, we examined performance using 2, 3, 4, or 5 questions on the EPDS and in the PHQ-9 for validation and compared performance by R^2^, RMSE, and MAE. As expected, there was a dose-dependent relationship where including additional questions improved model performance (higher R^2^, lower RMSE and MAE), with the exception that R^2^ improvement from 4 to 5 questions in the PHQ-9 was not statistically significant (Figure 3 and Table S-X).

**Figure 3:**
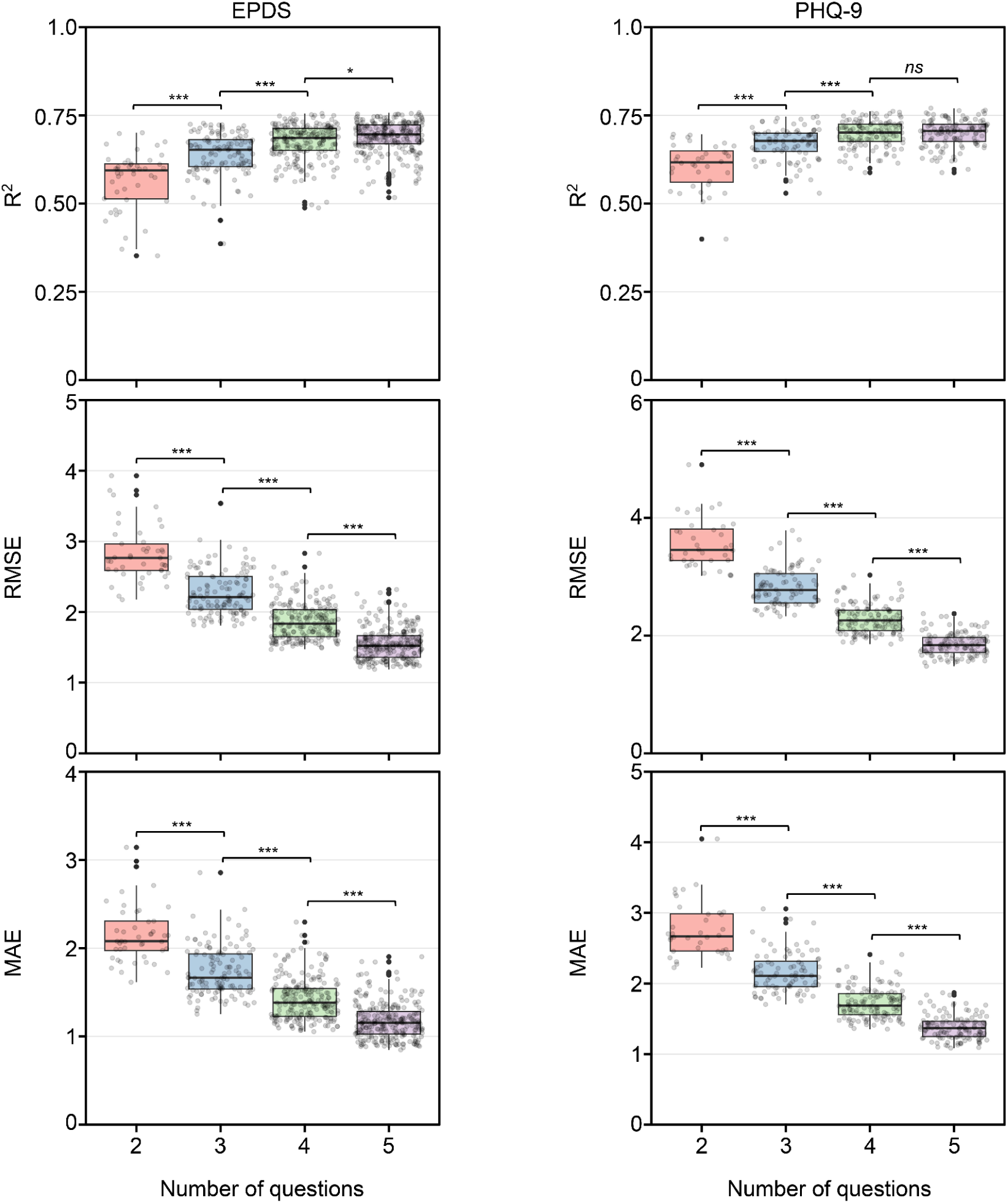
Adding additional questions improved model performance predicting the remaining total score of psychometric assessments. Box-plots showing model performance metrics for predicting the total scores using shortened versions of the EPDS (left) and PHQ-9 (right) questionnaires (represented by colored bars) from women who took the EPDS and all individuals who took the PHQ-9 in N3C. Each gray dot represents an individual ML model testing a different combination of 2, 3, 4, or 5 questions to predict the total score for the EPDS and PHQ-9. The top panel displays R^2^ values (coefficient of determination), the middle panel shows RMSE, while the bottom panel shows MAE. Overall, both EPDS and PHQ-9 assessments demonstrated improved model performance with additional questions, evidenced by higher R^2^ values and lower RMSE and MAE. * = P < 0.05, ** = P < 0.01, *** = P < 0.001 after Bonferroni correction

### Clinical trade-offs between ML and other screening approaches

We compared our ML approach against simple additive scoring, where three combinations were selected a priori: Q4:anxious + Q8:sad and Q5:scared + Q8:sad (top-performing ML combinations) and Q2:enjoy + Q8:sad (EPDS equivalent of the PHQ-2, included as an established benchmark). We evaluated four strategies: 1) simple addition, 2) ML classification, 3) ensemble OR logic (positive if either the simple addition or ML classification method is positive), and 4) ensemble AND logic (positive if both the simple addition and ML classification methods are positive). Performance was compared using F1 score, kappa, precision, and recall.

A key challenge in implementing screening is balancing identification of true cases (sensitivity/recall) versus avoiding unnecessary follow-ups from false positives (precision). Traditional additive scoring demonstrated high sensitivity (84-91%) but lower precision (54-77%), functioning as a “wide net” that minimizes missed cases but may burden healthcare systems with unnecessary evaluations. The ML approach achieved higher precision (80-86%) with moderate sensitivity (68-73%). Ensemble approaches amplified these trade-offs: OR logic maximized case detection (recall 92-97%) but had the lowest precision (39-54%), while AND logic matched ML performance (Figure 4A).

**Figure 4:**
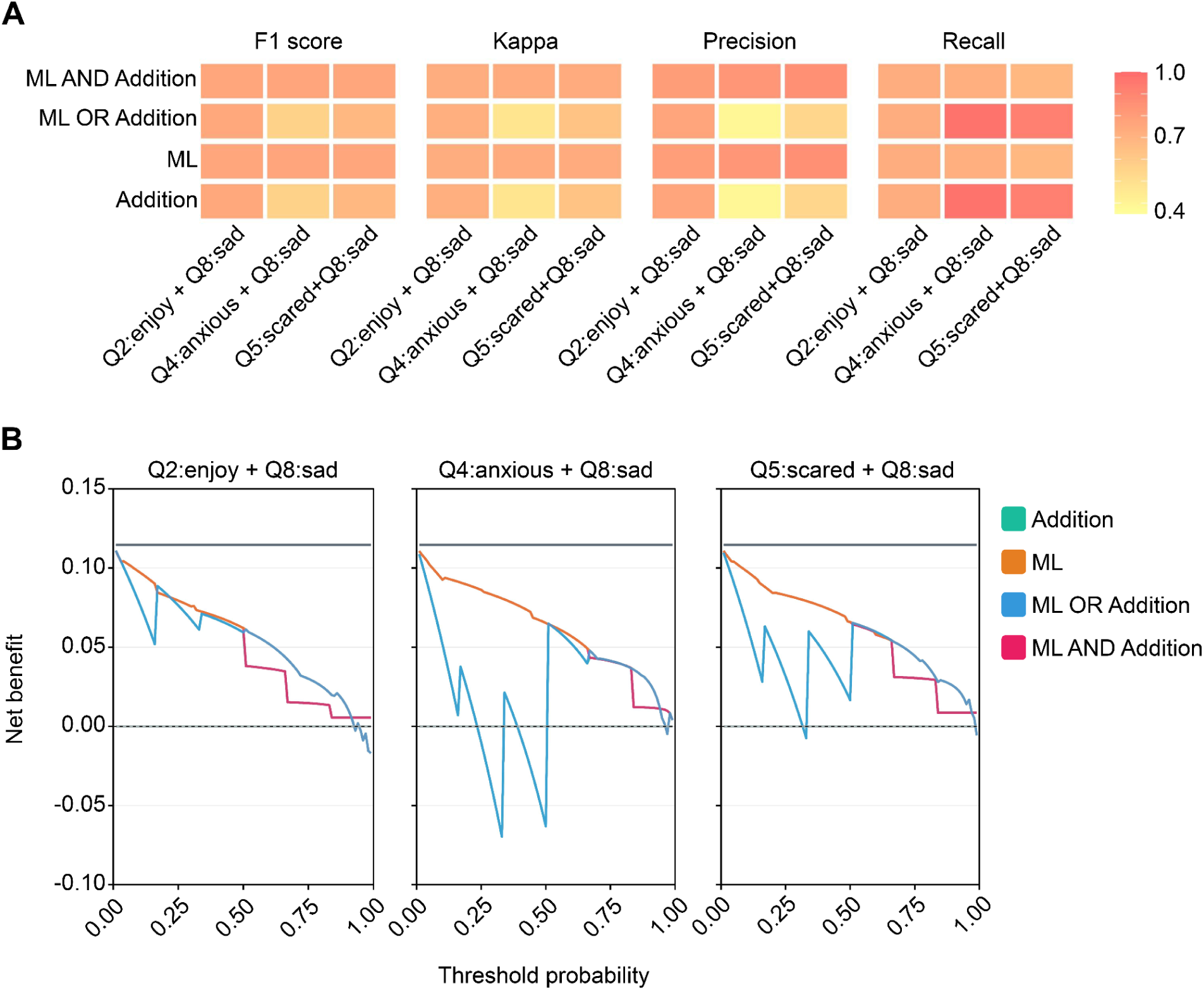
Our ML approach demonstrated improved net benefit across question combinations for PPD screening. **A.** Performance comparison of EPDS question combinations using different screening methods. ML-based approaches achieved higher F1 scores, kappa values, and precision compared to the other methods. **B.** Decision curve analysis evaluating net benefit of different EPDS question combinations and screening approaches. Our ML method demonstrated improved net benefit over traditional approaches, with the greatest advantage observed at lower threshold probabilities. Decision curves for individual methods are also presented in Figure S8 for improved visibility.

Decision curve analysis evaluates net benefit by accounting for the fact that different clinical settings have different thresholds for acting on a positive screen (some clinicians may refer patients at lower suspicion, others only at high suspicion). Across clinically relevant thresholds (0–80%; representing the range of minimum predicted depression probabilities at which a clinician would refer for further evaluation), the ML approach provided greater net benefit compared to simple additive scoring, meaning it identified more true cases while generating fewer unnecessary referrals regardless of the clinical setting’s risk tolerance (Figure 4B and Figure S8). The ensemble OR approach (positive if either the simple addition or ML classification method is positive), despite its high sensitivity, frequently produced negative net benefit, meaning it performed worse than simply screening no one, particularly for Q4:anxious + Q8:sad. The ensemble AND approach (positive if both the simple addition and ML classification methods are positive) showed minimal net benefit across most thresholds, offering little clinical advantage over the reference strategies..

## Discussion

### Principal findings

We developed a generalizable ML framework that accurately predicts full-length EPDS scores from two questions with improved net benefit compared to traditional additive scoring. Applied to >23,000 women across multiple cohorts, our method consistently identified Q4:anxious + Q8:sad and Q5:scared + Q8:sad as optimal pairs that maintained robust predictive performance while reducing burden. Decision curve analysis revealed our ML approach provided consistently improved net benefit across clinically relevant treatment thresholds. Validated across demographically diverse populations including postpartum and pregnant women, this work addresses the clinical urgency underscored by COVID-19’s lasting impact on perinatal mental health, with depression prevalence reaching 30% compared to pre-pandemic estimates of 10-20%, alongside challenges including workforce shortages and reliance on remote care^13,42,43^.

### Results in the context of what is known

Shortened psychometric assessments reduce participant burden in research and clinical practice^3^. Methods have primarily used psychometric approaches such as item-response theory, computerized adaptive testing, and classical test theory, though ML approaches have occasionally been applied with the goal of maximizing prediction accuracy^5–9^. Because the EPDS contains only 10 questions, we evaluated all possible combinations while prioritizing 2-question subsets for practical implementation.

Compared to prior EPDS abbreviation efforts, our study had the largest sample size of nearly 8,000 more individuals than the next largest study^18–23^. Interestingly, Q5:scared + Q8:sad was also identified by Choi et al.^18^ In previous literature, Q4:anxious appeared in 2 of 8 shortened versions, Q5:scared in 3 of 8, and Q8:sad in 6 of 8. Q8’s consistent presence reflects sadness as a core depressive symptom^18–23^. The selection of Q4:anxious or Q5:scared alongside sadness reflects the EPDS’s dual focus on depressive and anxiety symptomatology, mirroring clinical practice and high prevalence of perinatal anxiety symptoms^44,45^. While 3-question subsets showed incrementally better performance on average, our top 2-question combinations (Q4+Q8, Q5+Q8) performed comparably to the best 3-question combination (R^2^=0.70 vs. 0.73), supporting a 2-question recommendation given evidence that longer questionnaires are associated with significantly lower response rates. Our study was the only one using ML to select optimal 2-question EPDS subsets. While ML in psychometrics has gained attention in other fields, its use in mental health assessments remains limited^3,9,46^.

### Clinical implications

The most clinically relevant finding was that our ML approach demonstrated improved net benefit in decision curve analysis, which evaluates screening utility by weighing the benefit of identifying true cases against the cost of unnecessary referrals across a range of clinical risk thresholds^37,38,47^. This optimization became particularly important during COVID-19, when healthcare systems faced unprecedented strain with limited resources^42,43,48^. Our ML approach provides net benefit across diverse clinical decision thresholds, enabling implementation from well-resourced academic centers to community settings with limited infrastructure, which is essential for healthcare resilience during future emergencies^49,50^. Beyond improving detection efficiency, earlier identification of perinatal depression enables timely treatment initiation, which is associated with better maternal and infant outcomes; notably, screening itself (independent of downstream treatment) has been associated with reduced depression risk through decreased stigma and increased awareness^51^.

While the net benefit improvement appears modest (0.01-0.03), this translates to 1-3 additional positive screens per 100 screened, which is highly impactful at population scale. With approximately 3.6 million annual U.S. births, theoretical widespread implementation could identify up to 100,000 additional positive perinatal depression screens annually compared to simple additive scoring, where every detected case matters for maternal and infant outcomes^52^. This enhanced detection is critical in the post-pandemic era, where elevated depression prevalence and mental health workforce shortages necessitate maximally efficient screening^13,53,54^.

Furthermore, our abbreviated ML-based approach would enable improved longitudinal screening, overcoming the limitation that a positive result during a single screen may reflect transient psychological distress rather than clinically significant depression. A prospective Danish cohort study found that nearly 30% of women who initially screened positive on the EPDS showed spontaneous, clinically meaningful symptom resolution within 1-4 weeks without intervention, and a two-stage screening approach reduced unnecessary referrals by 24% while preserving detection of enduring cases^55^. The abbreviated ML-based 2-question format developed would be particularly well-suited to support such serial screening strategies: by substantially reducing respondent burden, it enables administration of the EPDS at multiple timepoints across the perinatal period, facilitating a more dynamic picture of symptom trajectory.

Consistent positive screens across two administrations, rather than a single threshold crossing, may represent a more clinically meaningful threshold for referral, serving as an approach that directly addresses the over-pathologization concern while preserving sensitivity for women with persistent depression.

Increased positive screens does raise concerns given existing clinician shortages. However, COVID-19 catalyzed rapid expansion of digital health interventions for perinatal mental health, including digital cognitive behavioral therapy, smartphone monitoring, telepsychiatry, and peer support platforms that have demonstrated efficacy while increasing access^26,53,56,57^. Our ML-optimized brief screening (with multiple positive screens) would be well-suited for integration into these digital ecosystems, enabling efficient remote assessment that can trigger automated care pathways or just-in-time interventions^58,59^. Pandemic-accelerated telehealth acceptance also created an opportune moment for implementing scalable, technology-enabled screening programs that reach underserved populations^60^.

### Research Implications

Future research should prospectively validate this abbreviated screening in real-world implementation to assess impact on screening rates and detection of perinatal depression. While we used linear and logistic regression for interpretability, exploration of ensemble methods like random forest or gradient boosting may further optimize performance^61,62^. For longer assessments, genetic algorithms could efficiently identify optimal question subsets^3^. Extension of this framework to other perinatal mental health measures warrants investigation. Finally, implementation studies examining integration of ML-enhanced brief screening with digital health interventions would inform scalable deployment strategies in resource-constrained settings.

### Strengths and limitations

Key strengths include our generalizable ML framework applicable to any standardized assessment, the largest EPDS abbreviation study to date, and the first application of ML methods to EPDS shortening. We validated performance across multiple independent cohorts including postpartum and pregnant populations, and confirmed methodology generalizability using PHQ-9 data. Our study limitations include EPDS data from only four N3C data partner sites, which may have variability in EPDS administration, documentation practices, and workflow differences, though our cohort demonstrates racial and ethnic diversity. We used linear and logistic regression models prioritizing simplicity and clinical interpretability over complex ensemble methods. While these models demonstrated strong performance, more sophisticated approaches may yield incremental improvements. Additionally, our study predicted total EPDS scores rather than clinical diagnoses, which represents an important limitation; future studies should validate abbreviated screening performance against structured clinical interviews. While we used the standard cutoff of ≥13, this threshold is not universally optimal, a large individual patient data meta-analysis by Levis et al. demonstrated that optimal EPDS cutoffs vary across populations and settings, and different thresholds may influence model performance. Importantly, this framework is flexible and can be retrained using alternative thresholds (e.g., ≥10) or question combinations to suit the clinical context, though prospective validation would be needed for any such adaptation. Lastly, because EPDS total scores are bounded (0–30) and derived from ordinal item-level responses, our use of linear regression makes distributional assumptions, which may not always hold true. In practice, predicted scores fell within the plausible range and performance metrics were consistent across cohorts, but future work could explore ordinal or bounded-outcome regression approaches (e.g., ordinal logistic regression, beta regression) to formally account for these properties.

## Conclusions

Beyond immediate application to perinatal depression screening, this generalizable framework provides a systematic approach to optimize standardized mental health instruments. Successful application to PHQ-9 demonstrates broader applicability. The ability to predict full assessment scores from two questions enables efficient longitudinal monitoring and reduces patient fatigue. The brief format offers particular advantages during healthcare system strain^48–50^. The increased telehealth use since COVID-19 presents opportunities to combine shortened assessments with ML approaches to improve remote screening^60^. Most importantly, by making effective screening more accessible and less burdensome, this framework addresses the persistent gap between clinical guidelines recommending routine screening and real-world implementation, offering a scalable solution that could improve perinatal mental health outcomes across diverse clinical settings and populations.

### N3C Attribution

The analyses described in this publication were conducted with data or tools accessed through the NCATS N3C Data Enclave https://covid.cd2h.org and N3C Attribution & Publication Policy v1.2-2020-08-25b supported by NCATS Contract No. 75N95023D00001 and the Center for Data to Health U24TR002306. This research was possible because of the patients whose information is included within the data and the organizations (https://ncats.nih.gov/n3c/resources/data-contribution/data-transfer-agreement-signatories) and scientists who have contributed to the on-going development of this community resource [https://doi.org/10.1093/jamia/ocaa196].

Disclaimer

The N3C Publication committee confirmed that this [manuscript/abstract/poster/presentation] msid:2655.082 is in accordance with N3C data use and attribution policies; however, this content is solely the responsibility of the authors and does not necessarily represent the official views of the National Institutes of Health or the N3C program.

### IRB

The N3C data transfer to NCATS is performed under a Johns Hopkins University Reliance Protocol # IRB00249128 or individual site agreements with NIH. The N3C Data Enclave is managed under the authority of the NIH; information can be found at https://ncats.nih.gov/n3c/resources.

### Individual Acknowledgements For Core Contributors

We gratefully acknowledge the following core contributors to N3C: Adam B. Wilcox, Adam M. Lee, Alexis Graves, Alfred (Jerrod) Anzalone, Amin Manna, Amit Saha, Amy Olex, Andrea Zhou, Andrew E. Williams, Andrew M. Southerland, Andrew T. Girvin, Anita Walden, Anjali Sharathkumar, Benjamin Amor, Benjamin Bates, Brian Hendricks, Brijesh Patel, G. Caleb Alexander, Carolyn T. Bramante, Cavin Ward-Caviness, Charisse Madlock-Brown, Christine Suver, Christopher G. Chute, Christopher Dillon, Chunlei Wu, Clare Schmitt, Cliff Takemoto, Dan Housman, Davera Gabriel, David A. Eichmann, Diego Mazzotti, Donald E. Brown, Eilis Boudreau, Elaine L. Hill, Emily Carlson Marti, Emily R. Pfaff, Evan French, Farrukh M Koraishy, Federico Mariona, Fred Prior, George Sokos, Greg Martin, Harold P. Lehmann, Heidi Spratt, Hemalkumar B. Mehta, J.W. Awori Hayanga, Jami Pincavitch, Jaylyn Clark, Jeremy Richard Harper, Jessica Yasmine Islam, Jin Ge, Joel Gagnier, Johanna J. Loomba, John B. Buse, Jomol Mathew, Joni L. Rutter, Julie A. McMurry, Justin Guinney, Justin Starren, Karen Crowley, Katie Rebecca Bradwell, Kellie M. Walters, Ken Wilkins, Kenneth R. Gersing, Kenrick Cato, Kimberly Murray, Kristin Kostka, Lavance Northington, Lee Pyles, Lesley Cottrell, Lili M. Portilla, Mariam Deacy, Mark M. Bissell, Marshall Clark, Mary Emmett, Matvey B. Palchuk, Melissa A. Haendel, Meredith Adams, Meredith Temple-O’Connor, Michael G. Kurilla, Michele Morris, Nasia Safdar, Nicole Garbarini, Noha Sharafeldin, Ofer Sadan, Patricia A. Francis, Penny Wung Burgoon, Philip R.O. Payne, Randeep Jawa, Rebecca Erwin-Cohen, Rena C. Patel, Richard A. Moffitt, Richard L. Zhu, Rishikesan Kamaleswaran, Robert Hurley, Robert T. Miller, Saiju Pyarajan, Sam G. Michael, Samuel Bozzette, Sandeep K. Mallipattu, Satyanarayana Vedula, Scott Chapman, Shawn T. O’Neil, Soko Setoguchi, Stephanie S. Hong, Steven G. Johnson, Tellen D. Bennett, Tiffany J. Callahan, Umit Topaloglu, Valery Gordon, Vignesh Subbian, Warren A. Kibbe, Wenndy Hernandez, Will Beasley, Will Cooper, William Hillegass, Xiaohan Tanner Zhang. Details of contributions available at covid.cd2h.org/core-contributors.

### Data Partners with Released Data

The following institutions whose data is released or pending:

Available: Advocate Health Care Network — UL1TR002389: The Institute for Translational Medicine (ITM) • Aurora Health Care Inc — UL1TR002373: Wisconsin Network For Health Research • Boston University Medical Campus — UL1TR001430: Boston University Clinical and Translational Science Institute • Brown University — U54GM115677: Advance Clinical Translational Research (Advance-CTR) • Carilion Clinic — UL1TR003015: iTHRIV Integrated Translational health Research Institute of Virginia • Case Western Reserve University — UL1TR002548: The Clinical & Translational Science Collaborative of Cleveland (CTSC) • Charleston Area Medical Center — U54GM104942: West Virginia Clinical and Translational Science Institute (WVCTSI) • Children’s Hospital Colorado — UL1TR002535: Colorado Clinical and Translational Sciences Institute • Columbia University Irving Medical Center — UL1TR001873: Irving Institute for Clinical and Translational Research • Dartmouth College — None (Voluntary) Duke University — UL1TR002553: Duke Clinical and Translational Science Institute • George Washington Children’s Research Institute — UL1TR001876: Clinical and Translational Science Institute at Children’s National (CTSA-CN) • George Washington University — UL1TR001876: Clinical and Translational Science Institute at Children’s National (CTSA-CN) • Harvard Medical School — UL1TR002541: Harvard Catalyst • Indiana University School of Medicine — UL1TR002529: Indiana Clinical and Translational Science Institute • Johns Hopkins University — UL1TR003098: Johns Hopkins Institute for Clinical and Translational Research • Louisiana Public Health Institute — None (Voluntary) • Loyola Medicine — Loyola University Medical Center • Loyola University Medical Center — UL1TR002389: The Institute for Translational Medicine (ITM) • Maine Medical Center — U54GM115516: Northern New England Clinical & Translational Research (NNE-CTR) Network • Mary Hitchcock Memorial Hospital & Dartmouth Hitchcock Clinic — None (Voluntary) • Massachusetts General Brigham — UL1TR002541: Harvard Catalyst • Mayo Clinic Rochester — UL1TR002377: Mayo Clinic Center for Clinical and Translational Science (CCaTS) • Medical University of South Carolina — UL1TR001450: South Carolina Clinical & Translational Research Institute (SCTR) • MITRE Corporation — None (Voluntary) • Montefiore Medical Center — UL1TR002556: Institute for Clinical and Translational Research at Einstein and Montefiore • Nemours — U54GM104941: Delaware CTR ACCEL Program • NorthShore University HealthSystem — UL1TR002389: The Institute for Translational Medicine (ITM) • Northwestern University at Chicago — UL1TR001422: Northwestern University Clinical and Translational Science Institute (NUCATS) • OCHIN — INV-018455: Bill and Melinda Gates Foundation grant to Sage Bionetworks • Oregon Health & Science University — UL1TR002369: Oregon Clinical and Translational Research Institute • Penn State Health Milton S. Hershey Medical Center — UL1TR002014: Penn State Clinical and Translational Science Institute • Rush University Medical Center — UL1TR002389: The Institute for Translational Medicine (ITM) • Rutgers, The State University of New Jersey — UL1TR003017: New Jersey Alliance for Clinical and Translational Science • Stony Brook University — U24TR002306 • The Alliance at the University of Puerto Rico, Medical Sciences Campus — U54GM133807: Hispanic Alliance for Clinical and Translational Research (The Alliance) • The Ohio State University — UL1TR002733: Center for Clinical and Translational Science • The State University of New York at Buffalo — UL1TR001412: Clinical and Translational Science Institute • The University of Chicago — UL1TR002389: The Institute for Translational Medicine (ITM) • The University of Iowa — UL1TR002537: Institute for Clinical and Translational Science • The University of Miami Leonard M. Miller School of Medicine — UL1TR002736: University of Miami Clinical and Translational Science Institute • The University of Michigan at Ann Arbor — UL1TR002240: Michigan Institute for Clinical and Health Research • The University of Texas Health Science Center at Houston — UL1TR003167: Center for Clinical and Translational Sciences (CCTS) • The University of Texas Medical Branch at Galveston — UL1TR001439: The Institute for Translational Sciences • The University of Utah — UL1TR002538: Uhealth Center for Clinical and Translational Science • Tufts Medical Center — UL1TR002544: Tufts Clinical and Translational Science Institute • Tulane University — UL1TR003096: Center for Clinical and Translational Science • The Queens Medical Center — None (Voluntary) • University Medical Center New Orleans — U54GM104940: Louisiana Clinical and Translational Science (LA CaTS) Center • University of Alabama at Birmingham — UL1TR003096: Center for Clinical and Translational Science • University of Arkansas for Medical Sciences — UL1TR003107: UAMS Translational Research Institute • University of Cincinnati — UL1TR001425: Center for Clinical and Translational Science and Training • University of Colorado Denver, Anschutz Medical Campus — UL1TR002535: Colorado Clinical and Translational Sciences Institute • University of Illinois at Chicago — UL1TR002003: UIC Center for Clinical and Translational Science • University of Kansas Medical Center — UL1TR002366: Frontiers: University of Kansas Clinical and Translational Science Institute • University of Kentucky — UL1TR001998: UK Center for Clinical and Translational Science • University of Massachusetts Medical School Worcester — UL1TR001453: The UMass Center for Clinical and Translational Science (UMCCTS) • University Medical Center of Southern Nevada — None (voluntary) • University of Minnesota — UL1TR002494: Clinical and Translational Science Institute • University of Mississippi Medical Center — U54GM115428: Mississippi Center for Clinical and Translational Research (CCTR) • University of Nebraska Medical Center — U54GM115458: Great Plains IDeA-Clinical & Translational Research • University of North Carolina at Chapel Hill — UL1TR002489: North Carolina Translational and Clinical Science Institute • University of Oklahoma Health Sciences Center — U54GM104938: Oklahoma Clinical and Translational Science Institute (OCTSI) • University of Pittsburgh — UL1TR001857: The Clinical and Translational Science Institute (CTSI) • University of Pennsylvania — UL1TR001878: Institute for Translational Medicine and Therapeutics • University of Rochester — UL1TR002001: UR Clinical & Translational Science Institute • University of Southern California — UL1TR001855: The Southern California Clinical and Translational Science Institute (SC CTSI) • University of Vermont — U54GM115516: Northern New England Clinical & Translational Research (NNE-CTR) Network • University of Virginia — UL1TR003015: iTHRIV Integrated Translational health Research Institute of Virginia • University of Washington — UL1TR002319: Institute of Translational Health Sciences • University of Wisconsin-Madison — UL1TR002373: UW Institute for Clinical and Translational Research • Vanderbilt University Medical Center — UL1TR002243: Vanderbilt Institute for Clinical and Translational Research • Virginia Commonwealth University — UL1TR002649: C. Kenneth and Dianne Wright Center for Clinical and Translational Research • Wake Forest University Health Sciences — UL1TR001420: Wake Forest Clinical and Translational Science Institute • Washington University in St. Louis — UL1TR002345: Institute of Clinical and Translational Sciences • Weill Medical College of Cornell University — UL1TR002384: Weill Cornell Medicine Clinical and Translational Science Center • West Virginia University — U54GM104942: West Virginia Clinical and Translational Science Institute (WVCTSI) Submitted: Icahn School of Medicine at Mount Sinai — UL1TR001433: ConduITS Institute for Translational Sciences • The University of Texas Health Science Center at Tyler — UL1TR003167: Center for Clinical and Translational Sciences (CCTS) • University of California, Davis — UL1TR001860: UCDavis Health Clinical and Translational Science Center • University of California, Irvine — UL1TR001414: The UC Irvine Institute for Clinical and Translational Science (ICTS) • University of California, Los Angeles — UL1TR001881: UCLA Clinical Translational Science Institute • University of California, San Diego — UL1TR001442: Altman Clinical and Translational Research Institute • University of California, San Francisco — UL1TR001872: UCSF Clinical and Translational Science Institute NYU Langone Health Clinical Science Core, Data Resource Core, and PASC Biorepository Core — OTA-21-015A: Post-Acute Sequelae of SARS-CoV-2 Infection Initiative (RECOVER) Pending: Arkansas Children’s Hospital — UL1TR003107: UAMS Translational Research Institute • Baylor College of Medicine — None (Voluntary) • Children’s Hospital of Philadelphia — UL1TR001878: Institute for Translational Medicine and Therapeutics • Cincinnati Children’s Hospital Medical Center — UL1TR001425: Center for Clinical and Translational Science and Training • Emory University — UL1TR002378: Georgia Clinical and Translational Science Alliance • HonorHealth — None (Voluntary) • Loyola University Chicago — UL1TR002389: The Institute for Translational Medicine (ITM) • Medical College of Wisconsin — UL1TR001436: Clinical and Translational Science Institute of Southeast Wisconsin • MedStar Health Research Institute — None (Voluntary) • Georgetown University — UL1TR001409: The Georgetown-Howard Universities Center for Clinical and Translational Science (GHUCCTS) • MetroHealth — None (Voluntary) • Montana State University — U54GM115371: American Indian/Alaska Native CTR • NYU Langone Medical Center — UL1TR001445: Langone Health’s Clinical and Translational Science Institute • Ochsner Medical Center — U54GM104940: Louisiana Clinical and Translational Science (LA CaTS) Center • Regenstrief Institute — UL1TR002529: Indiana Clinical and Translational Science Institute • Sanford Research — None (Voluntary) • Stanford University — UL1TR003142: Spectrum: The Stanford Center for Clinical and Translational Research and Education • The Rockefeller University — UL1TR001866: Center for Clinical and Translational Science • The Scripps Research Institute — UL1TR002550: Scripps Research Translational Institute • University of Florida — UL1TR001427: UF Clinical and Translational Science Institute • University of New Mexico Health Sciences Center — UL1TR001449: University of New Mexico Clinical and Translational Science Center • University of Texas Health Science Center at San Antonio — UL1TR002645: Institute for Integration of Medicine and Science • Yale New Haven Hospital — UL1TR001863: Yale Center for Clinical Investigation

## Data Availability

The National Clinical Cohort Collaborative (N3C) data transfer to the National Center for Advancing Translational Sciences is performed under a Johns Hopkins University Reliance Protocol (IRB00249128) or individual site agreements with the National Institutes of Health (NIH). The N3C Data Enclave is managed under the authority of the NIH; information can be found. N3C Enclave data are protected and can be accessed for COVID-19-related research with an NIH-approved 1) IRB protocol and 2) institutional Data Use Request (DUR ID: RP-E39D65). A detailed accounting of data protections and access tiers is found at the NCATS website. N3C Enclave and data access instructions can be found at the N3C website; all codes used to produce the analyses in this manuscript are available within the N3C Enclave to users with valid log-in credentials to support reproducibility.

## Data sharing statement

The National Clinical Cohort Collaborative (N3C) data transfer to the National Center for Advancing Translational Sciences is performed under a Johns Hopkins University Reliance Protocol (IRB00249128) or individual site agreements with the National Institutes of Health (NIH). The N3C Data Enclave is managed under the authority of the NIH; information can be found^63^. N3C Enclave data are protected and can be accessed for COVID-19–related research with an NIH-approved 1) IRB protocol and 2) institutional Data Use Request (DUR ID: RP-E39D65). A detailed accounting of data protections and access tiers is found at the NCATS website^63^. N3C Enclave and data access instructions can be found at the N3C website^64^; all codes used to produce the analyses in this manuscript are available within the N3C Enclave to users with valid log-in credentials to support reproducibility.

## Code availability

Code is available for use upon request to the corresponding author (EH).

## Author contributions

EH conceived the study and analyzed the data. EH, CS, KC, VB, CS, and MAH interpreted the results. MAH was responsible for resources. EH was responsible for the original manuscript draft preparation. EH was responsible for visualization. MAH was responsible for supervision and acquired funding. EH, CS, KC, VB, RCP, CS, and MAH were responsible for reviewing and editing the manuscript for publication.

## Use of generative AI

Chat-GPT, developed by Open-AI^65^, and Claude, developed by Anthropic^66^, were used to edit some portions of the manuscript, such as grammar, sentence structure, and synonyms. All content generated from these models were reviewed by authors and were not used for idea generation.

