## Supplementary Material for "Machine learning-optimized perinatal depression screening: Maximum impact, minimal burden"

### Supplementary methods

#### Detailed N3C infrastructure and data governance

The National COVID Cohort Collaborative (N3C) Enclave consists of a limited dataset of COVID-19 cases and controls and includes their electronic health record (EHR) data from clinical sites across the United States[1]. The harmonized data is incorporated in N3C from individuals who are COVID-19 positive and negative from January 1, 2020 to present with weekly updates to include near real-time procedures and clinical visits, with historical data dating back to January 1, 2018[1]. Ingestion of data into N3C matches COVID-19 positive individuals with two COVID-19 negative controls based on up to four sociodemographic variables (age, sex, race, and ethnicity) when available from the data partner site[1]. The National Institutes of Health (NIH) Institutional Review Board (IRB) approved the N3C Enclave. Individual data partner sites maintain their own local IRB-approved data transfer agreements or operate under a Johns Hopkins University Reliance Protocol (IRB00249128). Each investigator with access to the Enclave receives IRB approval from their institution.

#### EPDS and PHQ-9 phenotyping - detailed methods

EPDS Phenotyping

Item-level responses to the EPDS in N3C were obtained using OMOP concepts listed in Table S-I and came from the Observation table. EPDS symptom classifications followed Harel et al., which maps each EPDS question to specific symptom domains including anxiety, depression, and thoughts of self-harm[2]. We included individuals with female sex at birth who completed all 10 EPDS items on the same date. When multiple assessments existed for the same individual, we selected the assessment with the highest total score to optimize representation of individuals with elevated scores, as the cohort was expected to contain a larger proportion of individuals with lower scores.

PHQ-9 phenotyping

Item-level responses to the PHQ-9 in N3C were obtained using OMOP concepts listed in eTable 1 and came from the Observation table. PHQ-9 items were based on the Diagnostic and Statistical Manual of Mental Disorders (DSM)-5 criteria for major depressive disorder[3]. We then filtered for complete assessments where individuals answered all items on the same date and calculated total scores by summing all item-level responses. We did not perform any sex-based exclusion for the PHQ-9 since the goal was for methodological validation rather than creating a shortened PHQ-9 assessment specific to any population. When multiple assessments existed for the same individual, we selected the assessment with the highest total score to optimize representation of individuals with elevated scores.

The availability of item-level EPDS data from the pandemic period is important given documented shifts in perinatal mental health symptom presentation and severity during and after COVID-19[4–6]. The pandemic-driven expansion of digital assessment administration underscores the need for brief, efficient screening tools that minimize patient burden while maintaining diagnostic utility in resource-constrained healthcare environments.

**Table S-I: OMOP concept IDs for item-level responses to the EPDS and PHQ-9 in N3C.**

| Assessment | Question number | OMOP concept ID | OMOP concept name | Symptom captured/abbreviation |
|---|---|---|---|---|
| EPDS | 1 | 42870286 | I have been able to laugh and see the funny side of things in the past 7 days [EPDS] | Funny |
| EPDS | 2 | 42870287 | I have looked forward with enjoyment to things in the past 7 days [EPDS] | Enjoy |
| EPDS | 3 | 42870288 | I have blamed myself unnecessarily when things went wrong in the past 7 days [EPDS] | Blame |
| EPDS | 4 | 42870289 | I have been anxious or worried for no good reason in the past 7 days [EPDS] | Anxious |
| EPDS | 5 | 42870290 | I have felt scared or panicky for no very good reason in the past 7 days [EPDS] | Scared |

| | | | | |
|---|---|---|---|---|
| EPDS | 6 | 42870291 | Things have been getting on top of me in the past 7 days [EPDS] | Overwhelmed |
| EPDS | 7 | 42870292 | I have been so unhappy that I have had difficulty sleeping in the past 7 days [EPDS] | Sleep |
| EPDS | 8 | 42870293 | I have felt sad or miserable in the past 7 days [EPDS] | Sad |
| EPDS | 9 | 42870294 | I have been so unhappy that I have been crying in the past 7 days [EPDS] | Cry |
| EPDS | 10 | 42870295 | The thought of harming myself has occurred to me in the past 7 days [EPDS] | Harm |
| PHQ-9 | 1 | 3042924 | Little interest or pleasure in doing things in last 2 weeks | Enjoy |
| PHQ-9 | 2 | 3045858 | Feeling down, depressed, or hopeless in last 2 weeks | Sad |

| | | | | |
|---|---|---|---|---|
| PHQ-9 | 3 | 3045933 | Trouble falling or staying asleep, or sleeping too much in last 2 weeks [Reported.PHQ] | Sleep |
| PHQ-9 | 4 | 3044964 | Feeling tired or having little energy in last 2 weeks [Reported.PHQ] | Tired |
| PHQ-9 | 5 | 3044098 | Poor appetite or overeating in last 2 weeks [Reported.PHQ] | Appetite |
| PHQ-9 | 6 | 3043801 | Feeling bad about yourself - or that you are a failure or have let yourself or your family down in last 2 weeks [Reported.PHQ] | Guilt |
| PHQ-9 | 7 | 3045019 | Trouble concentrating on things, such as reading the newspaper or watching television in | Concentration |

| | | | | |
|---|---|---|---|---|
| | | | last 2 weeks [Reported.PHQ] | |
| PHQ-9 | 8 | 3043785 | Moving or speaking so slowly that other people could have noticed. Or the opposite - being so fidgety or restless that you have been moving around a lot more than usual in last 2 weeks [Reported.PHQ] | Psychomotor |
| PHQ-9 | 9 | 3043462 | Thoughts that you would be better off dead, or of hurting yourself in some way in last 2 weeks [Reported.PHQ] | Harm |

**Calculating the average total score per question response**

The mean and standard deviation of the total score for individuals who responded 0-3 for each of the individual questions was calculated to determine the relationship between responses to individual questions in the EPDS and PHQ-9 and the total score. To determine if the average score per individual question response was significantly different, we performed linear regression models assessing the relationship between the total score and individual questions. We repeated the analysis using response 0, response 1, and response 2 as the reference at a significance level of 0.05 with Bonferroni correction to account for multiple comparisons[7].

**Detailed machine learning (ML) model development**

Model architecture and rationale

We developed our ML approach to shorten psychometric assessments based on the hypothesis that an individual's responses to a subset of questions could reliably predict their total score. The development of efficient screening approaches is needed in the context of pandemic-related healthcare system strain and the documented increases in perinatal depression prevalence, where healthcare systems must balance thorough assessment with limited clinical time and resources[8,9]. Each model was built using a framework that involved selecting a subset of questions, subtracting the responses to the selected subset of questions from the total score, and then using linear regression to predict the remaining portion of the score (Figure S1)[10]. This process was repeated for every combination of two, three, four, and five questions from the psychometric assessments used in this study, which were the EPDS and PHQ-9. We then identified the top five combinations of two questions with the best predictive performance, assessed by $R^2$, root mean square error (RMSE), and mean absolute error (MAE)[11]. All ML analyses were performed using the Caret package in R[12].

**Figure S1: A schematic displaying our ML method with 2-item subsets.**

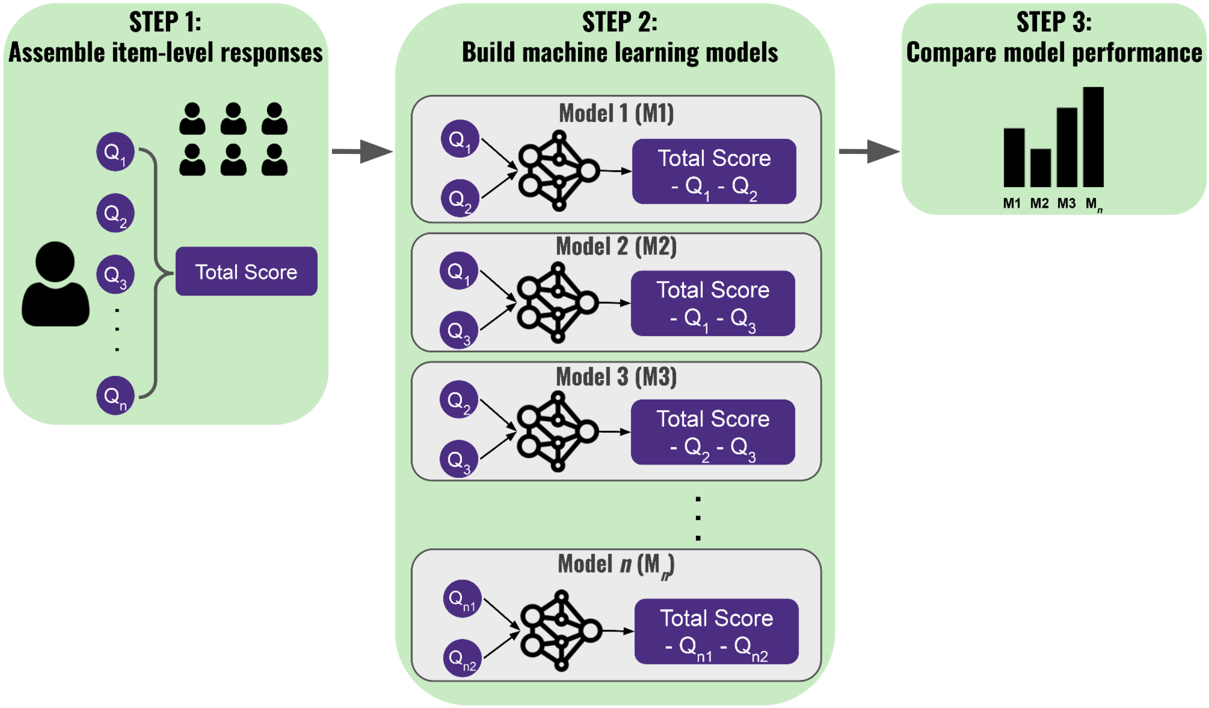


*Our three-step machine learning (ML) framework identifies the most predictive question subsets within mental health assessments (e.g., the EPDS). First, we gather item-level responses to a mental health assessment and calculate the total score by summing the individual responses (Step 1). Next, we develop ML models using each permutation of two items as inputs to predict the remaining score (total score minus the two input items) (Step 2). Finally, we compare performance metrics across all models to identify which two-item combination most accurately predicts the remaining score, effectively determining the optimal brief assessment version (Step 3).*

Binary classification models

After developing our ML approach to predict the remaining EPDS total score as a continuous outcome, we extended the method to binary classification. Binary classification models were constructed using top-performing question pairs identified from the continuous outcome model results or selected a priori based on clinical practice (e.g., using Q2:enjoy + Q8:sad, the EPDS equivalent of PHQ-2's Q1:enjoy + Q2:sad)[13]. Models used logistic regression with clinical cutoffs to predict positive or negative screening results (≥13 for EPDS and ≥10 for PHQ-9)[14,15]. The F1 score, calculated as the harmonic mean of precision and recall, provides a single balanced metric that equally weighs the ability to correctly identify true positive cases (recall/sensitivity) and minimize false positives (precision/positive predictive value), making it particularly useful for comparing overall screening effectiveness when there are trade-offs between these competing priorities[16].

**Sensitivity analysis in the postpartum period - detailed methods**

We aimed to evaluate whether the top-performing question pairs identified from the full EPDS cohort, which included any female in N3C with a recorded EPDS assessment, also performed well among women specifically in the postpartum period. To identify women in the postpartum period, we leveraged the Hierarchy and rule-based pregnancy episode Inference integrated with Pregnancy Progression Signatures (HIPPS) algorithm that has been developed and validated in N3C[17]. We refined our EPDS cohort to include women who completed the assessment between 4 weeks and 12 months postpartum. The 4-week exclusion period was selected to avoid confounding by "postpartum blues" ("baby blues"), which affect up to 85% of new mothers and typically resolve within 2-3 weeks postpartum[18]. Our goal was to capture EPDS testing within the first year postpartum, which represents the standard screening window for postpartum depression. We then reapplied our continuous ML approach, as outlined above, to the postpartum cohort to identify the top five pairs of questions with the highest predictive performance for the remaining total score. Binary classification models were then built using select top-performing pairs of questions.

**External validation cohort - detailed protocol**

For an additional layer of validation and to assess the generalizability of our method, we assessed our ML approach in a cohort external from N3C. The cohort comprised 1,217 pregnant women who participated in a prospective study conducted by Washington University in St. Louis (Wash U) between January 2017 and January 2020, followed by secondary analysis by Stanford University[19,20]. Given the data for this cohort was collected between 2017-2020 (pre-COVID-19) and the majority of the N3C cohort was collected in the COVID-19 pandemic era, we also sought to assess whether the same (or different) subset of questions would emerge as most predictive of the EPDS total score. Briefly, Institutional Review Board approval for this study was obtained from Wash U School of Medicine Human Research Protection Office. The study enrolled English-speaking pregnant individuals aged ≥18 years with singleton pregnancies ≤20 weeks gestation who planned to deliver at Barnes-Jewish Hospital. Participants were seen longitudinally at study visits throughout each trimester, wore actigraphy devices continuously, and completed validated questionnaires about sleep habits and lifestyle.

This cohort differs from N3C in its prospective design with active longitudinal follow-up and standardized data collection protocols, compared to N3C's retrospective EHR data from multiple healthcare systems. Additional details about the cohort can be seen in a prior publication[20]. This cohort provided item-level responses to the EPDS in a different study setting and different demographic distribution compared to the N3C EPDS cohort. We then re-applied our continuous and binary classification ML models as described earlier. Demographic categorical variables between the N3C entire and Wash U cohorts were compared using Chi-square tests[21]. Two separate tests were performed: one comparing EPDS screening rates and one comparing race/ethnicity distributions between cohorts. Age was compared using an unpaired, two-sided t-test.

We re-applied our continuous and binary classification ML models as described earlier, using identical methods to those used in the N3C cohorts to ensure comparability of results.

**Covariate definitions**

Demographics of age and race/ethnicity were obtained using standardized N3C-wide definitions. Age was recorded as a continuous variable in years at the time of EPDS assessment. Race/ethnicity categories included: White non-Hispanic, Black non-Hispanic, Hispanic/Latinx of any race, Other non-Hispanic (which includes Asian, Native American, Pacific Islander, and multiracial individuals), and Unknown (when race/ethnicity data was missing or not recorded). Pregnancy outcomes were identified for each person using the HIPPS algorithm[17]. Prior history of mental disorders was determined using condition diagnoses mapped to phecodes, a standardized system for high-throughput EHR-based phenotyping based on International Classification of Diseases (ICD) codes to identify diseases and conditions[22,23]. Mental health phecodes included in the analysis are listed in Table S-II and encompass major diagnostic categories including depressive disorders, anxiety disorders, bipolar disorder, schizophrenia spectrum disorders, substance use disorders, eating disorders, and other psychiatric conditions. A participant was considered to have prior mental health history if any mental health phecode appeared in their record before their EPDS assessment date.

**Table S-II: Phecodes for mental disorders.**

| Phecode | Description |
|---|---|
| 290.11 | Alzheimer's disease |
| 290.12 | Dementia with cerebral degenerations |
| 290 | Delirium dementia and amnestic and other cognitive disorders |
| 290.1 | Dementias |
| 290.2 | Delirium due to conditions classified elsewhere |
| 290.3 | Other persistent mental disorders due to conditions classified elsewhere |
| 290.13 | Senile dementia |
| 290.16 | Vascular dementia |
| 291.4 | Specific nonpsychotic mental disorders due to brain damage |
| 291.8 | Alteration of consciousness |
| 292 | Neurological disorders |
| 292.1 | Aphasia/speech disturbance |
| 292.11 | Aphasia |
| 292.12 | Symbolic dysfunction |
| 292.2 | Mild cognitive impairment |
| 292.3 | Memory loss |
| 292.4 | Altered mental status |
| 292.5 | Transient alteration of awareness |
| 292.6 | Hallucinations |
| 291 | Other specified nonpsychotic and/or transient mental disorders |
| 291.1 | Transient mental disorders due to conditions classified elsewhere |
| 295 | Schizophrenia and other psychotic disorders |
| 295.1 | Schizophrenia |
| 295.2 | Paranoid disorders |
| 295.3 | Psychosis |
| 296 | Mood disorders |

| | |
|---|---|
| 296.1 | Bipolar |
| 296.2 | Depression |
| 296.22 | Major depressive disorder |
| 297 | Suicidal ideation or attempt |
| 297.1 | Suicidal ideation |
| 297.2 | Suicide or self-inflicted injury |
| 300 | Anxiety disorders |
| 300.1 | Anxiety disorder |
| 300.11 | Generalized anxiety disorder |
| 300.12 | Agorophobia, social phobia, and panic disorder |
| 300.13 | Phobia |
| 300.2 | Generalized anxiety & phobic disorders |
| 300.3 | Obsessive-compulsive disorders |
| 300.4 | Dysthymic disorder |
| 300.8 | Acute reaction to stress |
| 300.9 | Posttraumatic stress disorder |
| 301 | Personality disorders |
| 301.1 | Schizoid personality disorder |
| 301.2 | Antisocial/borderline personality disorder |
| 302 | Sexual and gender identity disorders |
| 302.1 | Decreased libido |
| 303 | Psychogenic and somatoform disorders |
| 303.1 | Dissociative disorder |
| 303.3 | Psychogenic disorder |
| 303.31 | Gastrointestinal malfunction arising from mental factors |
| 303.4 | Somatoform disorder |
| 304 | Adjustment reaction |
| 305 | Disorders associated with eating |
| 305.2 | Eating disorder |
| 305.21 | Anorexia nervosa |
| 306 | Other mental disorder |

| | |
|---|---|
| 306.1 | Mental disorders during/after pregnancy |
| 306.9 | Tension headache |
| 293 | Symptoms involving head and neck |
| 293.1 | Swelling, mass, or lump in head and neck [Space-occupying lesion, intracranial NOS] |
| 313 | Pervasive developmental disorders |
| 313.1 | Attention deficit hyperactivity disorder |
| 313.2 | Tics and stuttering |
| 313.3 | Autism |
| 315 | Developmental delays and disorders |
| 315.1 | Learning disorder |
| 315.2 | Speech and language disorder |
| 315.3 | Intellectual disability |
| 316 | Substance addiction and disorders |
| 316.1 | Polyneuropathy due to drugs |
| 317 | Alcohol-related disorders |
| 317.1 | Alcoholism |
| 317.11 | Alcoholic liver damage |
| 318 | Tobacco use disorder |
| 312 | Conduct disorders |
| 312.3 | Impulse control disorder |

**Statistical methods for cohort comparisons**

Demographic categorical variables between the N3C entire and Wash U cohorts were compared using Chi-square tests[21]. Two separate Chi-square tests were performed: one comparing EPDS screening rates (positive vs negative, defined by total score ≥13 vs <13) and one comparing race/ethnicity distributions between cohorts. The Chi-square test statistic ($\chi^2$), degrees of freedom (df), and P-values are reported for both comparisons. Age distributions were compared using an unpaired, two-sided t-test, with results reported as mean difference and P-value. Statistical significance was set at $P<0.05$ for all comparisons. No adjustment for multiple comparisons was made for these descriptive demographic comparisons, as they were intended to characterize cohort differences rather than test primary hypotheses.

**Comparison of addition and ML approach performance - detailed methods**

Validated shortened assessments like the PHQ-2 use simple additive scoring, where responses to two questions are summed and compared against a threshold (typically ≥3 for positive screening)[24]. To evaluate our ML approach against this established method, we compared model performance using: 1) the optimal question combinations identified through our ML approach (Q4:anxious + Q8:sad and Q5:scared + Q8:sad), and 2) the EPDS equivalent of the PHQ-2 questions (Q2:enjoy + Q8:sad). We evaluated four classification strategies: 1) Simple addition with ≥3 threshold: Responses to the two questions are summed (range 0-6), and scores ≥3 are classified as positive screening, following the PHQ-2 convention[24]; 2) ML classification: Logistic regression models using the two question responses as predictors, with predicted probabilities >0.5 classified as positive. Models were trained using the clinical cutoff of ≥13 on the total EPDS score as the outcome; 3) Ensemble "OR" logic: Classified as positive if either the ML method OR the addition method predicts positive screening. This approach maximizes sensitivity at the potential cost of reduced specificity; and 4) Ensemble "AND" logic: Classified as positive only if both the ML method AND the addition method predict positive screening. This approach maximizes specificity at the potential cost of reduced sensitivity. Performance was assessed using confusion matrices with thresholds of ≥3 for the additive approach and ≥13 for defining true positive status based on total EPDS scores (standard cutoff for moderate to severe depression)[25,26]. We compared F1 score, kappa, precision, and recall across all four approaches for each question combination[16].

**Decision curve analysis - technical details**

We conducted decision curve analysis to evaluate the clinical utility of different EPDS screening strategies in the context of resource-constrained healthcare delivery systems (a challenge that became particularly acute during the COVID-19 pandemic and will remain relevant for future public health emergencies)[27,28]. Decision curve analysis compares the net benefit of prediction models across a range of threshold probabilities from 0.01 to 0.99[29,30]. Analyses were performed separately for three two-question combinations: 1) Q2:enjoy + Q8:sad (EPDS equivalent to PHQ-2); 2) Q4:anxious + Q8:sad (top-performing ML combination), 3) Q5:scared + Q8:sad (second top-performing ML combination). Depression was defined as a binary outcome based on a total EPDS score ≥13, as this threshold identifies moderate to severe depression and minimizes false positive results while maintaining acceptable sensitivity[26]. For each question pair, we computed a simple sum score (range 0-6) and generated binary indicators using a cutoff of ≥3[24]. The four prediction approaches included: 1) Simple addition method: Sum of responses scaled to probabilities (sum/6); 2) Logistic regression models: Predicted probabilities from ML models; 3) "OR" logic method: Maximum probability from the addition and ML models (takes the higher of the two predicted probabilities); and 4) "AND" logic method: Minimum probability from both models (takes the lower of the two predicted probabilities). Net benefit was calculated using the standard formula[29,30]:

$$\text{Net Benefit} = (TP/n) - (FP/n) \times (pt/(1-pt))$$

Where: TP = true positives (number correctly identified as having depression), FP = false positives (number incorrectly identified as having depression), n = total sample size, pt = threshold probability (the minimum probability at which a clinician would act). The term (pt/(1–pt)) represents the odds at the threshold probability and weights false positives relative to false negatives based on the chosen threshold. At higher thresholds, the model tolerates more false positives to avoid missing true cases; at lower thresholds, fewer false positives are acceptable.

Reference strategies included: "Treat all": Net benefit equal to the observed prevalence, assuming all individuals receive screening/treatment, "Treat none": Net benefit = 0, assuming no screening is performed.

Methods with net benefit exceeding both reference strategies across a range of threshold probabilities were considered to have clinical utility[29,30]. The threshold probability (pt) represents the minimum probability of disease at which a clinician would recommend treatment or further evaluation. By evaluating net benefit across a range of threshold probabilities, decision curve analysis accounts for varying clinical contexts and risk tolerances. For example, a clinician working in a setting with abundant mental health resources might use a lower threshold (treating more patients with lower predicted probabilities), while a resource-constrained setting might require a higher threshold (treating only those with higher predicted probabilities).

**Software**

All statistical analyses were performed using R (R Foundation for Statistical Computing[31]). ML models were developed using the Caret package[12].

#### Supplementary Results

##### EPDS testing frequency over time

Within the N3C database, EPDS data were contributed by a range of one to four data partner sites between January 2018 and October 2024. The frequency of EPDS administration demonstrated a substantial increase over time, reaching a peak of >15,000 administrations in early 2024 across four contributing sites. Notably, a marked escalation in EPDS utilization was observed coinciding with the onset of the COVID-19 pandemic in March 2020 and an additional one to two data partner sites contributing EPDS data (Figure S2). This increase likely reflects the expansion of data partner sites with item-level EPDS reporting capabilities rather than solely changes in clinical practice, with the majority of available data collected in the post-pandemic period.

**Figure S2: EPDS data availability in N3C over time.**

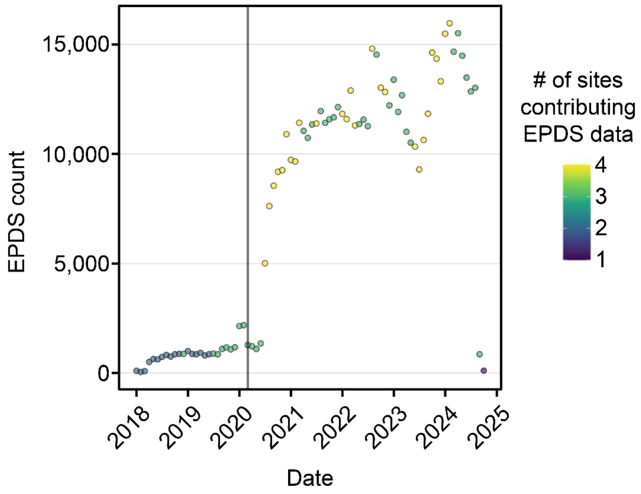


*The total number of EPDS assessments in N3C by month from January 2018 to October 2024 in N3C. Data points demonstrate an upward trend over time and are colored according to the number of contributing data partner sites. The vertical line at March 2020 represents the beginning of the COVID-19 pandemic.*

**Association between individual EPDS question responses and total scores**

To establish the conceptual foundation for our abbreviated screening approach, we first examined whether individual EPDS question responses were associated with total depression severity. For each of the 10 EPDS questions, we calculated mean total EPDS scores (with standard deviations) stratified by response category. We used linear regression to test whether total scores varied significantly across response levels. Figure S3 demonstrates a consistent dose-response relationship: women selecting more severe response categories for individual items had progressively higher total EPDS scores across all 10 questions, with all pairwise comparisons reaching statistical significance ($P<0.001$). For example, women responding "Yes, quite a lot" to Q4 (I have been anxious or worried for no good reason) had substantially higher mean total scores compared to those responding "No, not at all." One notable exception to the monotonic pattern was observed for Q1 (I have been able to laugh and see the funny side of things). While responses generally followed the expected gradient, women who responded "As much as I always could" (indicating preserved ability to laugh) had total scores similar to those responding "Not quite so much now" rather than showing the lowest scores. This deviation may reflect the positive framing of Q1 compared to the negatively-framed items, or could indicate that preserved humor does not necessarily exclude depressive symptoms in perinatal populations. Despite this single exception, the overall pattern strongly supported our hypothesis that individual question responses carry substantial information about total depression severity, providing empirical justification for developing abbreviated screening tools using question subsets rather than relying solely on new cutoff scores applied to the full 10-item instrument.

**Figure S3: Increased individual question responses are associated with increased EPDS total scores.**

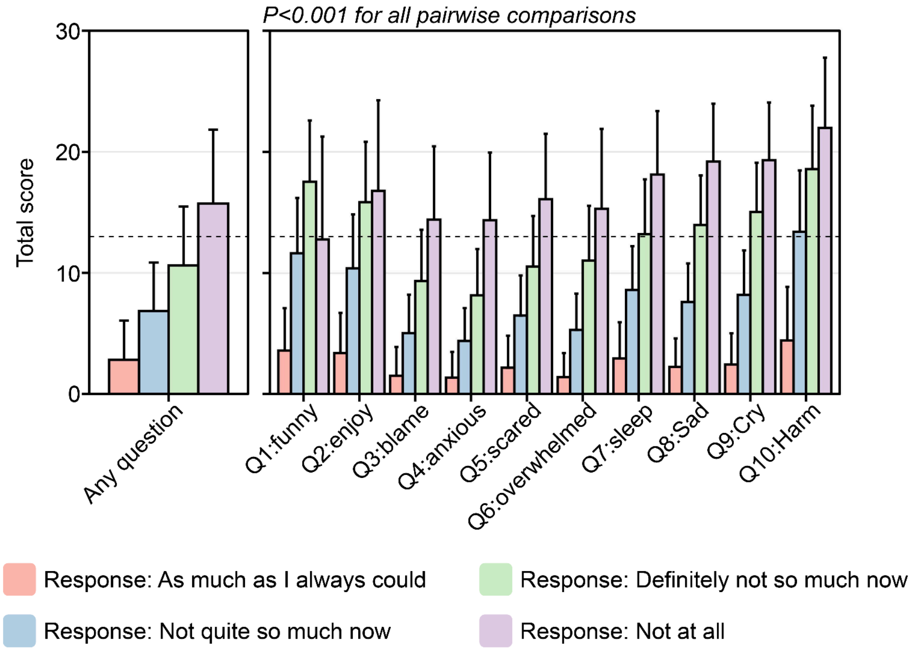


*The average EPDS total score based on responses to individual questions (Q1-Q10). The left panel shows overall scores by response to all questions, while the right panel breaks down scores by specific question. Four response categories are shown: "As much as I always could" (pink), "Not quite so much now" (blue), "Definitely not so much now" (green), and "Not at all" (purple). Data are expressed as mean and standard deviation. Statistical comparisons between each pair of response categories demonstrated significant differences in total EPDS scores across all comparisons (P<0.001). The horizontal dashed line represents a clinical cutoff score of 13 for positive depression screening.*

**Table S-III: ML model performance of all 2-item EPDS subsets.**

| Questions | $R^2$ | RMSE | MAE |
|---|---|---|---|
| 4,8 | 0.70 | 2.17 | 1.61 |
| 5,8 | 0.70 | 2.23 | 1.72 |
| 5,9 | 0.67 | 2.35 | 1.82 |
| 4,9 | 0.67 | 2.33 | 1.73 |
| 3,8 | 0.67 | 2.36 | 1.77 |
| 6,8 | 0.65 | 2.40 | 1.84 |
| 6,9 | 0.64 | 2.50 | 1.89 |
| 3,9 | 0.62 | 2.54 | 1.90 |
| 7,8 | 0.62 | 2.55 | 2.04 |
| 4,7 | 0.62 | 2.48 | 1.85 |
| 2,8 | 0.61 | 2.67 | 2.14 |
| 1,8 | 0.61 | 2.72 | 2.19 |
| 8,9 | 0.61 | 2.59 | 2.05 |
| 2,5 | 0.61 | 2.66 | 2.07 |
| 6,7 | 0.61 | 2.60 | 1.99 |
| 5,7 | 0.60 | 2.59 | 1.99 |
| 5,6 | 0.60 | 2.55 | 1.86 |
| 8,1 | 0.60 | 2.90 | 2.31 |
| 2,3 | 0.60 | 2.71 | 2.08 |
| 2,4 | 0.60 | 2.66 | 2.00 |
| 2,9 | 0.60 | 2.77 | 2.23 |
| 7,9 | 0.60 | 2.68 | 2.14 |
| 3,7 | 0.59 | 2.64 | 1.99 |
| 2,6 | 0.59 | 2.77 | 2.13 |
| 1,5 | 0.59 | 2.80 | 2.16 |
| 1,9 | 0.58 | 2.87 | 2.30 |
| 4,6 | 0.58 | 2.59 | 1.83 |
| 1,4 | 0.58 | 2.78 | 2.07 |

| | | | |
|---|---|---|---|
| 1,6 | 0.57 | 2.85 | 2.18 |
| 1,3 | 0.56 | 2.89 | 2.20 |
| 9,1 | 0.55 | 3.12 | 2.47 |
| 3,6 | 0.54 | 2.77 | 1.97 |
| 2,7 | 0.53 | 2.99 | 2.40 |
| 1,7 | 0.51 | 3.10 | 2.48 |
| 6,1 | 0.51 | 3.21 | 2.41 |
| 3,5 | 0.51 | 2.86 | 2.08 |
| 3,4 | 0.48 | 2.88 | 2.06 |
| 5,1 | 0.48 | 3.31 | 2.53 |
| 4,1 | 0.47 | 3.26 | 2.43 |
| 7,1 | 0.47 | 3.40 | 2.71 |
| 4,5 | 0.45 | 2.96 | 2.16 |
| 3,1 | 0.42 | 3.49 | 2.64 |
| 2,1 | 0.40 | 3.72 | 2.98 |
| 1,2 | 0.37 | 3.66 | 2.92 |

**EPDS question stability across COVID-19**

To assess whether the most predictive question combinations changed following the COVID-19 pandemic onset, we conducted separate analyses stratified by observation date (before versus after March 1, 2020). The most predictive question combinations assessed by $R^2$ remained largely consistent across the pandemic transition. Q4: anxiety appeared in top-performing pairs in both periods, paired with sadness-related items Q9:crying pre-pandemic and Q8:sadness post-pandemic, both capturing core depressive symptoms (Table S-IV).

**Table S-IV: Top five two-item EPDS questions most predictive of the total EPDS score pre- and post-COVID.**

| Pre-COVID | | Post-COVID | |
|---|---|---|---|
| **EPDS Questions** | **$R^2$** | **EPDS Questions** | **$R^2$** |
| 4,9 | 0.68 | 4,8 | 0.70 |
| 4,8 | 0.67 | 5,8 | 0.70 |
| 5,9 | 0.67 | 5,9 | 0.68 |
| 6,9 | 0.66 | 4,9 | 0.67 |
| 5,8 | 0.66 | 3,8 | 0.67 |

**Table S-V: Binary classification models using Q4:anxious + Q8sad and Q5:scared + Q8:sad can predict EPDS screening results.**

| Metric | Q4:anxious + Q8:sad | Q5:scared + Q8:sad |
|---|---|---|
| Accuracy | 0.95 | 0.95 |
| 95% confidence interval | (0.948, 0.953) | (0.949, 0.954) |
| Kappa | 0.74 | 0.74 |
| Sensitivity | 0.72 | 0.68 |
| Specificity | 0.98 | 0.99 |
| Positive predictive value | 0.83 | 0.86 |
| Negative predictive value | 0.96 | 0.96 |
| Prevalence | 0.11 | 0.11 |
| Detection rate | 0.08 | 0.08 |
| Detection prevalence | 0.10 | 0.09 |
| Balanced accuracy | 0.85 | 0.83 |
| Precision | 0.83 | 0.86 |
| Recall | 0.72 | 0.68 |
| F1 score | 0.77 | 0.76 |

**Table S-VI: Demographics of the postpartum cohort with EPDS data.**

| Group | N3C Entire cohort | N3C Postpartum cohort | Wash U cohort | Statistical comparison N3C Entire vs Wash U cohorts |
|---|---|---|---|---|
| **Total cohort** | **22,924** | **7,750** | **1,217** | |
| **EPDS screening:** Positive | 2,626 (11.5%) | 1,051 (13.6%) | 92 (7.6%) | $\chi^2$=17.17, df=1, *P*<0.001 |
| Negative | 20,298 (88.5%) | 6,699 (86.4%) | 1,125 (92.4%) | |
| **Race/ethnicity:** White NH | 11,190 (48.8%) | 3,322 (49.6%) | 495 (40.7%) | $\chi^2$=589.92, df=2, *P*<0.001 |
| Black NH | 5151 (22.5%) | 1,387 (20.7%) | 672 (55.2%) | |
| Hispanic/Latinx of any race | 4,226 (18.4%) | 1,172 (17.5%) | 50 (4.1%) | |
| Other NH | 1,318 (5.7%) | 512 (7.6%) | 0 (0%) | |
| Unknown | 1,039 (4.5%) | 306 (4.6%) | 0 (0%) | |
| **Age (years)** | 32 (27-37) | 30 (26-34) | 28 (24-32) | *P*<0.001 |

NH: non-Hispanic, df: degrees of freedom

Age is reported as median and interquartile range. Categorical variables between the N3C entire and Wash U cohorts were compared using Chi-square tests. Two separate tests were performed: one comparing EPDS screening rates and one comparing race/ethnicity distributions between cohorts. Age was compared using an unpaired, two-sided t-test.

**Figure S4: Increased individual question responses are associated with increased EPDS total scores in a postpartum cohort and external pregnancy cohort.**

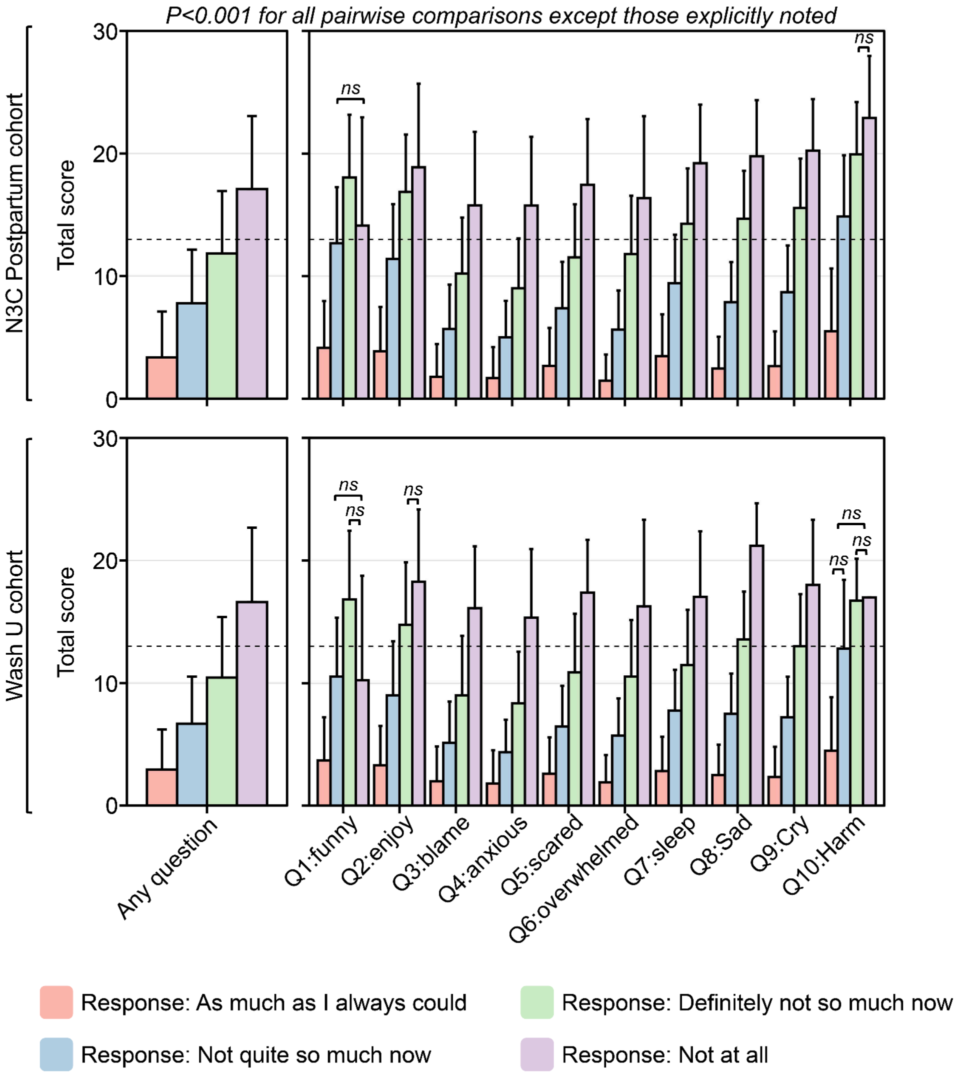


*The average EPDS total score among women in N3C in the postpartum period (between 4 weeks and 1 year after delivery) (top) and Stanford cohort (bottom) based on responses to individual questions (Q1-Q10). The left panel shows overall scores by response to all questions, while the right panel breaks down scores by specific question. Four response categories are shown: "As much as I always could" (pink), "Not quite so*

*much now" (blue), "Definitely not so much now" (green), and "Not at all" (purple). Data are expressed as mean and standard deviation. Statistical comparisons between each pair of response categories demonstrated significant differences in total EPDS scores across all comparisons (P<0.001) except for those specifically noted. The horizontal dashed line represents a clinical cutoff score of 13 for positive depression screening.*

Our ML method generalized to an assessment beyond the EPDS.

As methodological validation for our ML framework's generalizability beyond the EPDS, we applied it to the PHQ-9. We selected the PHQ-9 for validation because: 1) it provided a large sample size (n=398,606) for robust statistical testing, and 2) the existence of the established PHQ-2 allowed us to benchmark our ML-identified optimal questions against the current clinical standard[13]. This validation was designed to test whether our framework could consistently identify predictive question subsets across different psychometric instruments. The PHQ-9 cohort consisted of 398,606 distinct individuals with 151,106 (37.9%) individuals who screened positive (total score ≥10)[32–34]. The cohort was predominantly White Non-Hispanic (n=274,421, 68.8%) and the median age was 50 years old (IQR=31-68) (Table S-V).

Similar to our approach with the EPDS, we first evaluated the average total PHQ-9 score based on individual question responses and observed the same pattern as with the EPDS; total scores increased as the responses to individual questions increased (all pairwise comparisons were statistically significant: $P<0.001$) (Figure S5). We then performed our ML method and identified the optimal 2 questions to predict the remaining total PHQ-9 score assessed by $R^2$, RMSE, and MAE. The results indicated that Q2:sad + Q7:concentration was the best-performing model based on $R^2$ and RMSE, and ranked as the 4th-best combination when assessed by MAE (Figure S6 and Table S-VI). Interestingly, our ML approach identified Q2:sad + Q7:concentration rather than the PHQ-2's Q1:enjoy + Q2:sad. When we compared Q2:sad + Q7:concentration against the established PHQ-2 using binary classification with a threshold of 10, both models demonstrated comparable performance, with slightly better results observed for our ML-identified combination, validating the robustness of our methodology (Table S-VII).

**Table S-VII: Demographics of the N3C cohort with PHQ-9 data.**

| Group | Count (%) or Median (IQR) |
|---|---|
| **Total cohort** | **398,606** |
| ***PHQ-9 screening:*** Positive | 151,106 (37.9%) |
| Negative | 247,500 (62.1%) |
| ***Race/ethnicity:*** White NH | 171,896 (69.5%) |
| Black NH | 383,45 (15.5%) |
| Hispanic/Latinx of any race | 14,824 (6.0%) |
| Other NH | 14,711 (5.9%) |
| Unknown | 7,724 (3.1%) |
| ***Age (years)*** | 50 (31-68) |

NH: non-Hispanic

**Figure S5: Increased individual question responses are associated with increased PHQ-9 total scores.**

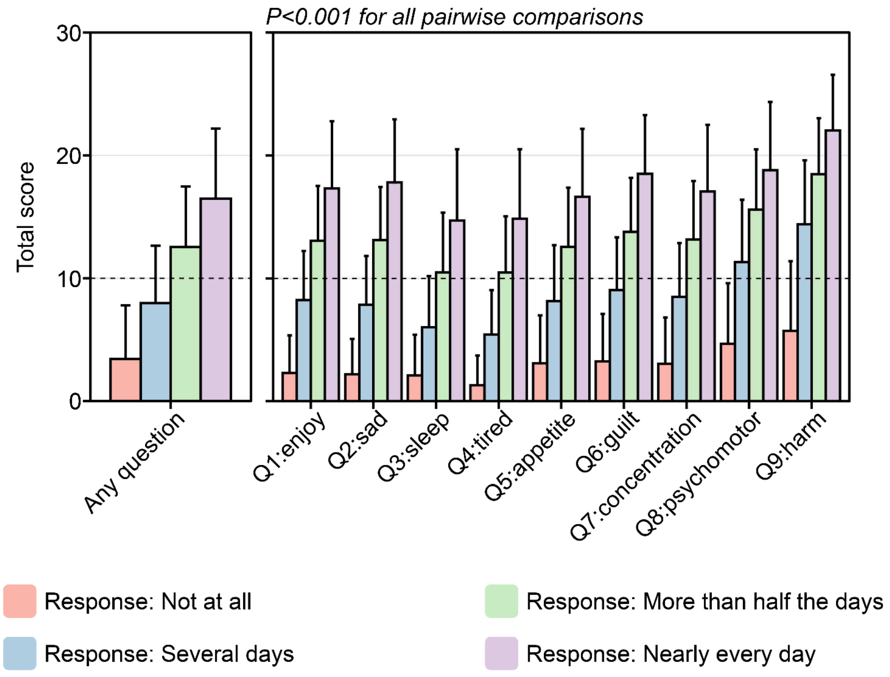


*The average PHQ-9 total score based on responses to individual questions (Q1-Q9). The left panel shows overall scores by response to all questions, while the right panel breaks down scores by specific question. Four response categories are shown: "Not at all" (pink), "Several days" (blue), "More than half the days" (green), and "Nearly every day" (purple). Data are expressed as mean and standard deviation. Statistical comparisons between each pair of response categories demonstrated significant differences in total PHQ-9 scores across all comparisons (P<0.001). The horizontal dashed line represents a clinical cutoff score of 10 for positive depression screening.*

**Figure S6: Q2+Q7 displayed the highest performance for predicting the remaining PHQ-9 score.**

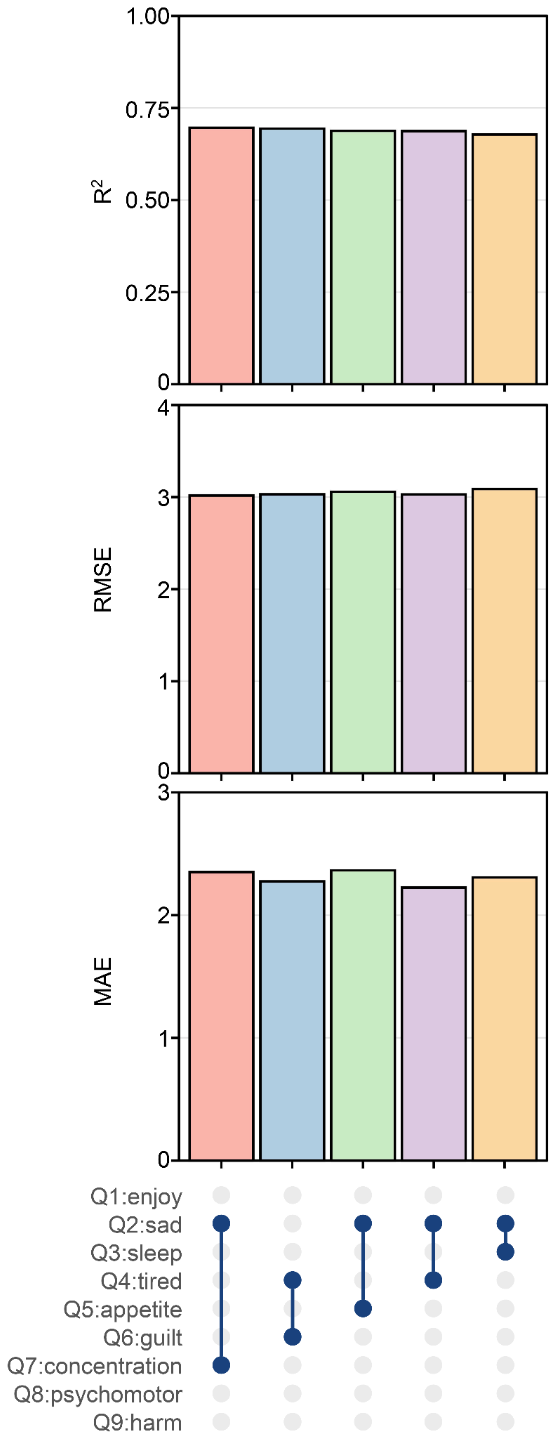

*Model performance metrics across the top five different models (represented by colored bars) using 2-item PHQ-9 question subsets to predict the PHQ-9 total score from the entire cohort of individuals who took the PHQ-9 in N3C. The top panel displays $R^2$ values (coefficient of determination), indicating similar predictive power across models. The middle panel shows RMSE, while the bottom panel shows MAE. The dot-and-line diagram at the bottom indicates which variables (Q1-Q9) were included in each model configuration, with blue dots representing included variables connected by vertical lines.*

**Table S-VIII: ML model performance of all 2-item PHQ-9 subsets.**

| Questions | $R^2$ | RMSE | MAE |
|---|---|---|---|
| 2,7 | 0.70 | 3.02 | 2.35 |
| 4,6 | 0.69 | 3.03 | 2.27 |
| 2,5 | 0.69 | 3.06 | 2.36 |
| 2,4 | 0.69 | 3.03 | 2.22 |
| 2,3 | 0.68 | 3.09 | 2.31 |
| 2,8 | 0.67 | 3.28 | 2.58 |
| 1,6 | 0.67 | 3.18 | 2.48 |
| 3,6 | 0.66 | 3.20 | 2.46 |
| 1,7 | 0.66 | 3.20 | 2.46 |
| 1,5 | 0.65 | 3.27 | 2.50 |
| 4,7 | 0.64 | 3.28 | 2.45 |
| 1,3 | 0.64 | 3.27 | 2.43 |
| 1,4 | 0.63 | 3.29 | 2.41 |
| 5,6 | 0.63 | 3.37 | 2.65 |
| 1,8 | 0.63 | 3.49 | 2.72 |
| 2,6 | 0.63 | 3.36 | 2.64 |
| 6,7 | 0.62 | 3.40 | 2.70 |
| 4,8 | 0.62 | 3.54 | 2.68 |
| 5,7 | 0.61 | 3.45 | 2.70 |
| 1,2 | 0.61 | 3.40 | 2.62 |
| 3,7 | 0.61 | 3.46 | 2.66 |
| 2,9 | 0.60 | 3.77 | 2.98 |
| 4,9 | 0.60 | 3.80 | 2.90 |
| 6,8 | 0.59 | 3.71 | 2.98 |
| 4,5 | 0.59 | 3.53 | 2.64 |
| 1,9 | 0.59 | 3.87 | 3.01 |
| 3,5 | 0.56 | 3.66 | 2.80 |
| 3,8 | 0.55 | 3.85 | 3.00 |

| | | | |
|---|---|---|---|
| 5,8 | 0.55 | 3.90 | 3.08 |
| 7,9 | 0.53 | 4.15 | 3.33 |
| 3,4 | 0.53 | 3.75 | 2.77 |
| 3,9 | 0.53 | 4.14 | 3.24 |
| 5,9 | 0.53 | 4.17 | 3.33 |
| 6,9 | 0.52 | 4.24 | 3.40 |
| 7,8 | 0.51 | 4.09 | 3.27 |
| 8,9 | 0.40 | 4.90 | 4.05 |

**Table S-IX: PHQ-9 binary classification model results using Q2+Q7 and Q1+Q2.**

| Metric | Q2+Q7 | Q1+Q2 |
|---|---|---|
| Accuracy | 0.89 | 0.88 |
| 95% confidence interval | (0.891, 0.893) | (0.877, 0.879) |
| Kappa | 0.77 | 0.74 |
| Sensitivity | 0.82 | 0.84 |
| Specificity | 0.94 | 0.90 |
| Positive predictive value | 0.89 | 0.84 |
| Negative predictive value | 0.89 | 0.90 |
| Prevalence | 0.38 | 0.38 |
| Detection rate | 0.31 | 0.32 |
| Detection prevalence | 0.35 | 0.38 |
| Balanced accuracy | 0.88 | 0.87 |
| Precision | 0.89 | 0.84 |
| Recall | 0.82 | 0.84 |
| F1 score | 0.85 | 0.84 |

**Table S-X: ML model performance of all 2, 3, 4, and 5-item EPDS and PHQ-9 subsets.**

| Assessment | Questions | # Questions | $R^2$ | RMSE | MAE |
|---|---|---|---|---|---|
| EPDS | 1,2 | 2 | 0.37 | 3.66 | 2.92 |
| EPDS | 1,3 | 2 | 0.56 | 2.89 | 2.20 |
| EPDS | 1,4 | 2 | 0.58 | 2.78 | 2.07 |
| EPDS | 1,5 | 2 | 0.59 | 2.80 | 2.16 |
| EPDS | 1,6 | 2 | 0.57 | 2.85 | 2.18 |
| EPDS | 1,7 | 2 | 0.51 | 3.10 | 2.48 |
| EPDS | 1,8 | 2 | 0.61 | 2.72 | 2.19 |
| EPDS | 1,9 | 2 | 0.58 | 2.87 | 2.30 |
| EPDS | 1,1 | 2 | 0.35 | 3.93 | 3.14 |
| EPDS | 2,3 | 2 | 0.60 | 2.71 | 2.08 |
| EPDS | 2,4 | 2 | 0.60 | 2.66 | 2.00 |
| EPDS | 2,5 | 2 | 0.61 | 2.66 | 2.07 |
| EPDS | 2,6 | 2 | 0.59 | 2.77 | 2.13 |
| EPDS | 2,7 | 2 | 0.53 | 2.99 | 2.40 |
| EPDS | 2,8 | 2 | 0.61 | 2.67 | 2.14 |
| EPDS | 2,9 | 2 | 0.60 | 2.77 | 2.23 |
| EPDS | 2,1 | 2 | 0.40 | 3.72 | 2.98 |
| EPDS | 3,4 | 2 | 0.48 | 2.88 | 2.06 |
| EPDS | 3,5 | 2 | 0.51 | 2.86 | 2.08 |
| EPDS | 3,6 | 2 | 0.54 | 2.77 | 1.97 |
| EPDS | 3,7 | 2 | 0.59 | 2.64 | 1.99 |
| EPDS | 3,8 | 2 | 0.67 | 2.36 | 1.77 |
| EPDS | 3,9 | 2 | 0.62 | 2.54 | 1.90 |
| EPDS | 3,1 | 2 | 0.42 | 3.49 | 2.64 |
| EPDS | 4,5 | 2 | 0.45 | 2.96 | 2.16 |
| EPDS | 4,6 | 2 | 0.58 | 2.59 | 1.83 |
| EPDS | 4,7 | 2 | 0.62 | 2.48 | 1.85 |
| EPDS | 4,8 | 2 | 0.70 | 2.17 | 1.61 |

| | | | | | |
|---|---|---|---|---|---|
| EPDS | 4,9 | 2 | 0.67 | 2.33 | 1.73 |
| EPDS | 4,1 | 2 | 0.47 | 3.26 | 2.43 |
| EPDS | 5,6 | 2 | 0.60 | 2.55 | 1.86 |
| EPDS | 5,7 | 2 | 0.60 | 2.59 | 1.99 |
| EPDS | 5,8 | 2 | 0.70 | 2.23 | 1.72 |
| EPDS | 5,9 | 2 | 0.67 | 2.35 | 1.82 |
| EPDS | 5,1 | 2 | 0.48 | 3.31 | 2.53 |
| EPDS | 6,7 | 2 | 0.61 | 2.60 | 1.99 |
| EPDS | 6,8 | 2 | 0.65 | 2.40 | 1.84 |
| EPDS | 6,9 | 2 | 0.64 | 2.50 | 1.89 |
| EPDS | 6,1 | 2 | 0.51 | 3.21 | 2.41 |
| EPDS | 7,8 | 2 | 0.62 | 2.55 | 2.04 |
| EPDS | 7,9 | 2 | 0.60 | 2.68 | 2.14 |
| EPDS | 7,1 | 2 | 0.47 | 3.40 | 2.71 |
| EPDS | 8,9 | 2 | 0.61 | 2.59 | 2.05 |
| EPDS | 8,1 | 2 | 0.60 | 2.90 | 2.31 |
| EPDS | 9,1 | 2 | 0.55 | 3.12 | 2.47 |
| EPDS | 1,2,3 | 3 | 0.59 | 2.55 | 1.96 |
| EPDS | 1,2,4 | 3 | 0.59 | 2.49 | 1.87 |
| EPDS | 1,2,5 | 3 | 0.60 | 2.51 | 1.96 |
| EPDS | 1,2,6 | 3 | 0.57 | 2.64 | 2.05 |
| EPDS | 1,2,7 | 3 | 0.50 | 2.89 | 2.33 |
| EPDS | 1,2,8 | 3 | 0.58 | 2.60 | 2.10 |
| EPDS | 1,2,9 | 3 | 0.57 | 2.69 | 2.17 |
| EPDS | 1,2,10 | 3 | 0.39 | 3.54 | 2.86 |
| EPDS | 1,3,4 | 3 | 0.64 | 2.20 | 1.59 |
| EPDS | 1,3,5 | 3 | 0.65 | 2.20 | 1.63 |
| EPDS | 1,3,6 | 3 | 0.64 | 2.24 | 1.65 |
| EPDS | 1,3,7 | 3 | 0.65 | 2.24 | 1.72 |
| EPDS | 1,3,8 | 3 | 0.70 | 2.06 | 1.57 |
| EPDS | 1,3,9 | 3 | 0.68 | 2.17 | 1.67 |
| EPDS | 1,3,10 | 3 | 0.57 | 2.79 | 2.14 |
| EPDS | 1,4,5 | 3 | 0.60 | 2.33 | 1.73 |

| | | | | | |
|---|---|---|---|---|---|
| EPDS | 1,4,6 | 3 | 0.67 | 2.10 | 1.51 |
| EPDS | 1,4,7 | 3 | 0.67 | 2.12 | 1.61 |
| EPDS | 1,4,8 | 3 | 0.73 | 1.91 | 1.44 |
| EPDS | 1,4,9 | 3 | 0.71 | 2.00 | 1.51 |
| EPDS | 1,4,10 | 3 | 0.60 | 2.64 | 2.00 |
| EPDS | 1,5,6 | 3 | 0.69 | 2.09 | 1.57 |
| EPDS | 1,5,7 | 3 | 0.66 | 2.23 | 1.76 |
| EPDS | 1,5,8 | 3 | 0.72 | 1.96 | 1.55 |
| EPDS | 1,5,9 | 3 | 0.71 | 2.03 | 1.60 |
| EPDS | 1,5,10 | 3 | 0.60 | 2.69 | 2.10 |
| EPDS | 1,6,7 | 3 | 0.63 | 2.32 | 1.82 |
| EPDS | 1,6,8 | 3 | 0.66 | 2.18 | 1.71 |
| EPDS | 1,6,9 | 3 | 0.66 | 2.24 | 1.74 |
| EPDS | 1,6,10 | 3 | 0.59 | 2.75 | 2.12 |
| EPDS | 1,7,8 | 3 | 0.62 | 2.38 | 1.93 |
| EPDS | 1,7,9 | 3 | 0.60 | 2.47 | 1.99 |
| EPDS | 1,7,10 | 3 | 0.52 | 3.02 | 2.44 |
| EPDS | 1,8,9 | 3 | 0.61 | 2.40 | 1.93 |
| EPDS | 1,8,10 | 3 | 0.61 | 2.66 | 2.14 |
| EPDS | 1,9,10 | 3 | 0.58 | 2.80 | 2.26 |
| EPDS | 2,3,4 | 3 | 0.66 | 2.09 | 1.53 |
| EPDS | 2,3,5 | 3 | 0.68 | 2.07 | 1.55 |
| EPDS | 2,3,6 | 3 | 0.66 | 2.15 | 1.59 |
| EPDS | 2,3,7 | 3 | 0.67 | 2.14 | 1.66 |
| EPDS | 2,3,8 | 3 | 0.70 | 2.00 | 1.54 |
| EPDS | 2,3,9 | 3 | 0.69 | 2.08 | 1.61 |
| EPDS | 2,3,10 | 3 | 0.61 | 2.63 | 2.04 |
| EPDS | 2,4,5 | 3 | 0.62 | 2.23 | 1.66 |
| EPDS | 2,4,6 | 3 | 0.67 | 2.05 | 1.48 |
| EPDS | 2,4,7 | 3 | 0.68 | 2.06 | 1.57 |
| EPDS | 2,4,8 | 3 | 0.72 | 1.88 | 1.42 |
| EPDS | 2,4,9 | 3 | 0.72 | 1.94 | 1.48 |
| EPDS | 2,4,10 | 3 | 0.62 | 2.54 | 1.93 |

| | | | | | |
|---|---|---|---|---|---|
| EPDS | 2,5,6 | 3 | 0.69 | 2.02 | 1.53 |
| EPDS | 2,5,7 | 3 | 0.67 | 2.15 | 1.71 |
| EPDS | 2,5,8 | 3 | 0.72 | 1.93 | 1.52 |
| EPDS | 2,5,9 | 3 | 0.72 | 1.96 | 1.56 |
| EPDS | 2,5,10 | 3 | 0.62 | 2.58 | 2.02 |
| EPDS | 2,6,7 | 3 | 0.64 | 2.27 | 1.79 |
| EPDS | 2,6,8 | 3 | 0.66 | 2.16 | 1.69 |
| EPDS | 2,6,9 | 3 | 0.66 | 2.19 | 1.72 |
| EPDS | 2,6,10 | 3 | 0.60 | 2.67 | 2.07 |
| EPDS | 2,7,8 | 3 | 0.62 | 2.34 | 1.90 |
| EPDS | 2,7,9 | 3 | 0.61 | 2.41 | 1.96 |
| EPDS | 2,7,10 | 3 | 0.53 | 2.92 | 2.36 |
| EPDS | 2,8,9 | 3 | 0.61 | 2.36 | 1.90 |
| EPDS | 2,8,10 | 3 | 0.61 | 2.62 | 2.11 |
| EPDS | 2,9,10 | 3 | 0.60 | 2.72 | 2.19 |
| EPDS | 3,4,5 | 3 | 0.45 | 2.53 | 1.78 |
| EPDS | 3,4,6 | 3 | 0.56 | 2.26 | 1.64 |
| EPDS | 3,4,7 | 3 | 0.65 | 2.04 | 1.46 |
| EPDS | 3,4,8 | 3 | 0.70 | 1.83 | 1.29 |
| EPDS | 3,4,9 | 3 | 0.67 | 1.98 | 1.40 |
| EPDS | 3,4,10 | 3 | 0.52 | 2.69 | 1.94 |
| EPDS | 3,5,6 | 3 | 0.59 | 2.22 | 1.54 |
| EPDS | 3,5,7 | 3 | 0.64 | 2.11 | 1.55 |
| EPDS | 3,5,8 | 3 | 0.71 | 1.86 | 1.37 |
| EPDS | 3,5,9 | 3 | 0.68 | 1.98 | 1.47 |
| EPDS | 3,5,10 | 3 | 0.53 | 2.71 | 1.99 |
| EPDS | 3,6,7 | 3 | 0.65 | 2.11 | 1.54 |
| EPDS | 3,6,8 | 3 | 0.67 | 1.99 | 1.44 |
| EPDS | 3,6,9 | 3 | 0.65 | 2.09 | 1.52 |
| EPDS | 3,6,10 | 3 | 0.57 | 2.63 | 1.90 |
| EPDS | 3,7,8 | 3 | 0.69 | 1.98 | 1.51 |
| EPDS | 3,7,9 | 3 | 0.66 | 2.09 | 1.59 |
| EPDS | 3,7,10 | 3 | 0.60 | 2.55 | 1.94 |

| | | | | | |
|---|---|---|---|---|---|
| EPDS | 3,8,9 | 3 | 0.66 | 2.07 | 1.57 |
| EPDS | 3,8,10 | 3 | 0.67 | 2.29 | 1.74 |
| EPDS | 3,9,10 | 3 | 0.63 | 2.46 | 1.86 |
| EPDS | 4,5,6 | 3 | 0.56 | 2.24 | 1.55 |
| EPDS | 4,5,7 | 3 | 0.61 | 2.16 | 1.60 |
| EPDS | 4,5,8 | 3 | 0.69 | 1.87 | 1.39 |
| EPDS | 4,5,9 | 3 | 0.67 | 1.99 | 1.47 |
| EPDS | 4,5,10 | 3 | 0.49 | 2.77 | 2.05 |
| EPDS | 4,6,7 | 3 | 0.68 | 1.94 | 1.38 |
| EPDS | 4,6,8 | 3 | 0.71 | 1.80 | 1.25 |
| EPDS | 4,6,9 | 3 | 0.70 | 1.89 | 1.32 |
| EPDS | 4,6,10 | 3 | 0.61 | 2.42 | 1.71 |
| EPDS | 4,7,8 | 3 | 0.72 | 1.81 | 1.35 |
| EPDS | 4,7,9 | 3 | 0.71 | 1.90 | 1.42 |
| EPDS | 4,7,10 | 3 | 0.64 | 2.37 | 1.78 |
| EPDS | 4,8,9 | 3 | 0.70 | 1.87 | 1.40 |
| EPDS | 4,8,10 | 3 | 0.71 | 2.08 | 1.56 |
| EPDS | 4,9,10 | 3 | 0.68 | 2.23 | 1.67 |
| EPDS | 5,6,7 | 3 | 0.68 | 1.99 | 1.49 |
| EPDS | 5,6,8 | 3 | 0.72 | 1.82 | 1.36 |
| EPDS | 5,6,9 | 3 | 0.71 | 1.88 | 1.40 |
| EPDS | 5,6,10 | 3 | 0.63 | 2.42 | 1.79 |
| EPDS | 5,7,8 | 3 | 0.70 | 1.91 | 1.51 |
| EPDS | 5,7,9 | 3 | 0.69 | 1.98 | 1.56 |
| EPDS | 5,7,10 | 3 | 0.61 | 2.50 | 1.93 |
| EPDS | 5,8,9 | 3 | 0.70 | 1.92 | 1.50 |
| EPDS | 5,8,10 | 3 | 0.70 | 2.15 | 1.68 |
| EPDS | 5,9,10 | 3 | 0.68 | 2.27 | 1.77 |
| EPDS | 6,7,8 | 3 | 0.66 | 2.08 | 1.62 |
| EPDS | 6,7,9 | 3 | 0.65 | 2.13 | 1.66 |
| EPDS | 6,7,10 | 3 | 0.61 | 2.51 | 1.95 |
| EPDS | 6,8,9 | 3 | 0.64 | 2.14 | 1.65 |
| EPDS | 6,8,10 | 3 | 0.66 | 2.33 | 1.80 |

| | | | | | |
|---|---|---|---|---|---|
| EPDS | 6,9,10 | 3 | 0.64 | 2.42 | 1.85 |
| EPDS | 7,8,9 | 3 | 0.60 | 2.30 | 1.83 |
| EPDS | 7,8,10 | 3 | 0.62 | 2.50 | 2.01 |
| EPDS | 7,9,10 | 3 | 0.60 | 2.62 | 2.11 |
| EPDS | 8,9,10 | 3 | 0.61 | 2.54 | 2.02 |
| EPDS | 1,2,3,4 | 4 | 0.65 | 1.92 | 1.41 |
| EPDS | 1,2,3,5 | 4 | 0.67 | 1.91 | 1.44 |
| EPDS | 1,2,3,6 | 4 | 0.64 | 2.01 | 1.50 |
| EPDS | 1,2,3,7 | 4 | 0.65 | 2.04 | 1.59 |
| EPDS | 1,2,3,8 | 4 | 0.68 | 1.91 | 1.48 |
| EPDS | 1,2,3,9 | 4 | 0.67 | 1.98 | 1.54 |
| EPDS | 1,2,3,10 | 4 | 0.59 | 2.48 | 1.93 |
| EPDS | 1,2,4,5 | 4 | 0.60 | 2.07 | 1.56 |
| EPDS | 1,2,4,6 | 4 | 0.66 | 1.91 | 1.39 |
| EPDS | 1,2,4,7 | 4 | 0.66 | 1.96 | 1.50 |
| EPDS | 1,2,4,8 | 4 | 0.70 | 1.79 | 1.36 |
| EPDS | 1,2,4,9 | 4 | 0.69 | 1.85 | 1.41 |
| EPDS | 1,2,4,10 | 4 | 0.60 | 2.39 | 1.81 |
| EPDS | 1,2,5,6 | 4 | 0.68 | 1.90 | 1.45 |
| EPDS | 1,2,5,7 | 4 | 0.64 | 2.06 | 1.64 |
| EPDS | 1,2,5,8 | 4 | 0.70 | 1.84 | 1.47 |
| EPDS | 1,2,5,9 | 4 | 0.70 | 1.88 | 1.50 |
| EPDS | 1,2,5,10 | 4 | 0.61 | 2.43 | 1.92 |
| EPDS | 1,2,6,7 | 4 | 0.60 | 2.19 | 1.74 |
| EPDS | 1,2,6,8 | 4 | 0.63 | 2.09 | 1.65 |
| EPDS | 1,2,6,9 | 4 | 0.63 | 2.12 | 1.68 |
| EPDS | 1,2,6,10 | 4 | 0.57 | 2.55 | 2.00 |
| EPDS | 1,2,7,8 | 4 | 0.57 | 2.29 | 1.87 |
| EPDS | 1,2,7,9 | 4 | 0.57 | 2.35 | 1.92 |
| EPDS | 1,2,7,10 | 4 | 0.50 | 2.83 | 2.30 |
| EPDS | 1,2,8,9 | 4 | 0.57 | 2.31 | 1.86 |
| EPDS | 1,2,8,10 | 4 | 0.58 | 2.55 | 2.07 |
| EPDS | 1,2,9,10 | 4 | 0.56 | 2.64 | 2.14 |

| | | | | | |
|---|---|---|---|---|---|
| EPDS | 1,3,4,5 | 4 | 0.62 | 1.90 | 1.36 |
| EPDS | 1,3,4,6 | 4 | 0.67 | 1.74 | 1.27 |
| EPDS | 1,3,4,7 | 4 | 0.72 | 1.65 | 1.21 |
| EPDS | 1,3,4,8 | 4 | 0.75 | 1.52 | 1.10 |
| EPDS | 1,3,4,9 | 4 | 0.73 | 1.61 | 1.17 |
| EPDS | 1,3,4,10 | 4 | 0.65 | 2.10 | 1.54 |
| EPDS | 1,3,5,6 | 4 | 0.70 | 1.72 | 1.23 |
| EPDS | 1,3,5,7 | 4 | 0.71 | 1.72 | 1.31 |
| EPDS | 1,3,5,8 | 4 | 0.75 | 1.55 | 1.17 |
| EPDS | 1,3,5,9 | 4 | 0.74 | 1.62 | 1.23 |
| EPDS | 1,3,5,10 | 4 | 0.66 | 2.12 | 1.58 |
| EPDS | 1,3,6,7 | 4 | 0.69 | 1.79 | 1.35 |
| EPDS | 1,3,6,8 | 4 | 0.70 | 1.72 | 1.29 |
| EPDS | 1,3,6,9 | 4 | 0.69 | 1.78 | 1.33 |
| EPDS | 1,3,6,10 | 4 | 0.65 | 2.16 | 1.60 |
| EPDS | 1,3,7,8 | 4 | 0.70 | 1.76 | 1.37 |
| EPDS | 1,3,7,9 | 4 | 0.69 | 1.84 | 1.42 |
| EPDS | 1,3,7,10 | 4 | 0.65 | 2.19 | 1.70 |
| EPDS | 1,3,8,9 | 4 | 0.68 | 1.84 | 1.42 |
| EPDS | 1,3,8,10 | 4 | 0.70 | 2.01 | 1.55 |
| EPDS | 1,3,9,10 | 4 | 0.67 | 2.12 | 1.64 |
| EPDS | 1,4,5,6 | 4 | 0.66 | 1.77 | 1.26 |
| EPDS | 1,4,5,7 | 4 | 0.66 | 1.80 | 1.37 |
| EPDS | 1,4,5,8 | 4 | 0.73 | 1.59 | 1.21 |
| EPDS | 1,4,5,9 | 4 | 0.72 | 1.66 | 1.25 |
| EPDS | 1,4,5,10 | 4 | 0.61 | 2.22 | 1.67 |
| EPDS | 1,4,6,7 | 4 | 0.72 | 1.65 | 1.21 |
| EPDS | 1,4,6,8 | 4 | 0.74 | 1.55 | 1.12 |
| EPDS | 1,4,6,9 | 4 | 0.74 | 1.60 | 1.15 |
| EPDS | 1,4,6,10 | 4 | 0.69 | 1.99 | 1.45 |
| EPDS | 1,4,7,8 | 4 | 0.73 | 1.61 | 1.24 |
| EPDS | 1,4,7,9 | 4 | 0.72 | 1.67 | 1.28 |
| EPDS | 1,4,7,10 | 4 | 0.68 | 2.05 | 1.57 |

| | | | | | |
|---|---|---|---|---|---|
| EPDS | 1,4,8,9 | 4 | 0.72 | 1.66 | 1.26 |
| EPDS | 1,4,8,10 | 4 | 0.73 | 1.84 | 1.39 |
| EPDS | 1,4,9,10 | 4 | 0.72 | 1.93 | 1.47 |
| EPDS | 1,5,6,7 | 4 | 0.71 | 1.70 | 1.31 |
| EPDS | 1,5,6,8 | 4 | 0.74 | 1.58 | 1.22 |
| EPDS | 1,5,6,9 | 4 | 0.75 | 1.60 | 1.23 |
| EPDS | 1,5,6,10 | 4 | 0.70 | 2.00 | 1.52 |
| EPDS | 1,5,7,8 | 4 | 0.71 | 1.72 | 1.38 |
| EPDS | 1,5,7,9 | 4 | 0.71 | 1.76 | 1.41 |
| EPDS | 1,5,7,10 | 4 | 0.66 | 2.17 | 1.72 |
| EPDS | 1,5,8,9 | 4 | 0.72 | 1.71 | 1.36 |
| EPDS | 1,5,8,10 | 4 | 0.72 | 1.91 | 1.52 |
| EPDS | 1,5,9,10 | 4 | 0.71 | 1.98 | 1.57 |
| EPDS | 1,6,7,8 | 4 | 0.65 | 1.93 | 1.53 |
| EPDS | 1,6,7,9 | 4 | 0.65 | 1.96 | 1.55 |
| EPDS | 1,6,7,10 | 4 | 0.63 | 2.26 | 1.79 |
| EPDS | 1,6,8,9 | 4 | 0.64 | 1.97 | 1.55 |
| EPDS | 1,6,8,10 | 4 | 0.66 | 2.13 | 1.67 |
| EPDS | 1,6,9,10 | 4 | 0.66 | 2.18 | 1.71 |
| EPDS | 1,7,8,9 | 4 | 0.58 | 2.16 | 1.74 |
| EPDS | 1,7,8,10 | 4 | 0.61 | 2.34 | 1.90 |
| EPDS | 1,7,9,10 | 4 | 0.60 | 2.43 | 1.97 |
| EPDS | 1,8,9,10 | 4 | 0.61 | 2.36 | 1.91 |
| EPDS | 2,3,4,5 | 4 | 0.64 | 1.79 | 1.30 |
| EPDS | 2,3,4,6 | 4 | 0.68 | 1.68 | 1.22 |
| EPDS | 2,3,4,7 | 4 | 0.73 | 1.58 | 1.17 |
| EPDS | 2,3,4,8 | 4 | 0.75 | 1.48 | 1.08 |
| EPDS | 2,3,4,9 | 4 | 0.74 | 1.54 | 1.13 |
| EPDS | 2,3,4,10 | 4 | 0.67 | 2.00 | 1.48 |
| EPDS | 2,3,5,6 | 4 | 0.71 | 1.64 | 1.19 |
| EPDS | 2,3,5,7 | 4 | 0.72 | 1.63 | 1.26 |
| EPDS | 2,3,5,8 | 4 | 0.76 | 1.50 | 1.14 |
| EPDS | 2,3,5,9 | 4 | 0.75 | 1.54 | 1.18 |

| | | | | | |
|---|---|---|---|---|---|
| EPDS | 2,3,5,10 | 4 | 0.68 | 2.00 | 1.51 |
| EPDS | 2,3,6,7 | 4 | 0.69 | 1.73 | 1.32 |
| EPDS | 2,3,6,8 | 4 | 0.70 | 1.69 | 1.27 |
| EPDS | 2,3,6,9 | 4 | 0.70 | 1.72 | 1.30 |
| EPDS | 2,3,6,10 | 4 | 0.66 | 2.08 | 1.55 |
| EPDS | 2,3,7,8 | 4 | 0.70 | 1.72 | 1.34 |
| EPDS | 2,3,7,9 | 4 | 0.70 | 1.77 | 1.38 |
| EPDS | 2,3,7,10 | 4 | 0.67 | 2.10 | 1.64 |
| EPDS | 2,3,8,9 | 4 | 0.68 | 1.79 | 1.38 |
| EPDS | 2,3,8,10 | 4 | 0.70 | 1.96 | 1.52 |
| EPDS | 2,3,9,10 | 4 | 0.69 | 2.04 | 1.59 |
| EPDS | 2,4,5,6 | 4 | 0.67 | 1.72 | 1.24 |
| EPDS | 2,4,5,7 | 4 | 0.67 | 1.74 | 1.33 |
| EPDS | 2,4,5,8 | 4 | 0.72 | 1.57 | 1.19 |
| EPDS | 2,4,5,9 | 4 | 0.72 | 1.60 | 1.22 |
| EPDS | 2,4,5,10 | 4 | 0.63 | 2.13 | 1.61 |
| EPDS | 2,4,6,7 | 4 | 0.72 | 1.61 | 1.19 |
| EPDS | 2,4,6,8 | 4 | 0.73 | 1.54 | 1.12 |
| EPDS | 2,4,6,9 | 4 | 0.73 | 1.57 | 1.14 |
| EPDS | 2,4,6,10 | 4 | 0.69 | 1.95 | 1.43 |
| EPDS | 2,4,7,8 | 4 | 0.73 | 1.59 | 1.22 |
| EPDS | 2,4,7,9 | 4 | 0.72 | 1.63 | 1.25 |
| EPDS | 2,4,7,10 | 4 | 0.69 | 1.99 | 1.53 |
| EPDS | 2,4,8,9 | 4 | 0.71 | 1.63 | 1.25 |
| EPDS | 2,4,8,10 | 4 | 0.73 | 1.81 | 1.38 |
| EPDS | 2,4,9,10 | 4 | 0.72 | 1.88 | 1.44 |
| EPDS | 2,5,6,7 | 4 | 0.71 | 1.66 | 1.28 |
| EPDS | 2,5,6,8 | 4 | 0.74 | 1.56 | 1.20 |
| EPDS | 2,5,6,9 | 4 | 0.75 | 1.56 | 1.21 |
| EPDS | 2,5,6,10 | 4 | 0.70 | 1.95 | 1.49 |
| EPDS | 2,5,7,8 | 4 | 0.71 | 1.69 | 1.36 |
| EPDS | 2,5,7,9 | 4 | 0.71 | 1.71 | 1.38 |
| EPDS | 2,5,7,10 | 4 | 0.67 | 2.10 | 1.67 |

| | | | | | |
|---|---|---|---|---|---|
| EPDS | 2,5,8,9 | 4 | 0.71 | 1.68 | 1.35 |
| EPDS | 2,5,8,10 | 4 | 0.72 | 1.88 | 1.49 |
| EPDS | 2,5,9,10 | 4 | 0.72 | 1.92 | 1.53 |
| EPDS | 2,6,7,8 | 4 | 0.64 | 1.91 | 1.52 |
| EPDS | 2,6,7,9 | 4 | 0.65 | 1.92 | 1.53 |
| EPDS | 2,6,7,10 | 4 | 0.63 | 2.22 | 1.76 |
| EPDS | 2,6,8,9 | 4 | 0.63 | 1.95 | 1.54 |
| EPDS | 2,6,8,10 | 4 | 0.66 | 2.11 | 1.66 |
| EPDS | 2,6,9,10 | 4 | 0.66 | 2.14 | 1.69 |
| EPDS | 2,7,8,9 | 4 | 0.58 | 2.13 | 1.72 |
| EPDS | 2,7,8,10 | 4 | 0.61 | 2.31 | 1.88 |
| EPDS | 2,7,9,10 | 4 | 0.60 | 2.37 | 1.94 |
| EPDS | 2,8,9,10 | 4 | 0.60 | 2.32 | 1.87 |
| EPDS | 3,4,5,6 | 4 | 0.50 | 2.00 | 1.41 |
| EPDS | 3,4,5,7 | 4 | 0.60 | 1.82 | 1.29 |
| EPDS | 3,4,5,8 | 4 | 0.67 | 1.60 | 1.12 |
| EPDS | 3,4,5,9 | 4 | 0.64 | 1.72 | 1.21 |
| EPDS | 3,4,5,10 | 4 | 0.49 | 2.37 | 1.69 |
| EPDS | 3,4,6,7 | 4 | 0.66 | 1.65 | 1.17 |
| EPDS | 3,4,6,8 | 4 | 0.69 | 1.55 | 1.10 |
| EPDS | 3,4,6,9 | 4 | 0.67 | 1.64 | 1.17 |
| EPDS | 3,4,6,10 | 4 | 0.59 | 2.11 | 1.53 |
| EPDS | 3,4,7,8 | 4 | 0.73 | 1.47 | 1.05 |
| EPDS | 3,4,7,9 | 4 | 0.71 | 1.57 | 1.12 |
| EPDS | 3,4,7,10 | 4 | 0.66 | 1.95 | 1.40 |
| EPDS | 3,4,8,9 | 4 | 0.69 | 1.57 | 1.12 |
| EPDS | 3,4,8,10 | 4 | 0.71 | 1.75 | 1.25 |
| EPDS | 3,4,9,10 | 4 | 0.68 | 1.89 | 1.36 |
| EPDS | 3,5,6,7 | 4 | 0.67 | 1.68 | 1.19 |
| EPDS | 3,5,6,8 | 4 | 0.70 | 1.56 | 1.09 |
| EPDS | 3,5,6,9 | 4 | 0.69 | 1.63 | 1.14 |
| EPDS | 3,5,6,10 | 4 | 0.61 | 2.10 | 1.48 |
| EPDS | 3,5,7,8 | 4 | 0.72 | 1.54 | 1.17 |

| | | | | | |
|---|---|---|---|---|---|
| EPDS | 3,5,7,9 | 4 | 0.70 | 1.62 | 1.22 |
| EPDS | 3,5,7,10 | 4 | 0.65 | 2.03 | 1.50 |
| EPDS | 3,5,8,9 | 4 | 0.70 | 1.60 | 1.19 |
| EPDS | 3,5,8,10 | 4 | 0.71 | 1.79 | 1.34 |
| EPDS | 3,5,9,10 | 4 | 0.69 | 1.91 | 1.42 |
| EPDS | 3,6,7,8 | 4 | 0.68 | 1.67 | 1.24 |
| EPDS | 3,6,7,9 | 4 | 0.67 | 1.73 | 1.28 |
| EPDS | 3,6,7,10 | 4 | 0.65 | 2.03 | 1.51 |
| EPDS | 3,6,8,9 | 4 | 0.64 | 1.77 | 1.30 |
| EPDS | 3,6,8,10 | 4 | 0.68 | 1.92 | 1.41 |
| EPDS | 3,6,9,10 | 4 | 0.66 | 2.02 | 1.48 |
| EPDS | 3,7,8,9 | 4 | 0.65 | 1.80 | 1.39 |
| EPDS | 3,7,8,10 | 4 | 0.68 | 1.93 | 1.48 |
| EPDS | 3,7,9,10 | 4 | 0.66 | 2.04 | 1.56 |
| EPDS | 3,8,9,10 | 4 | 0.66 | 2.02 | 1.53 |
| EPDS | 4,5,6,7 | 4 | 0.65 | 1.69 | 1.19 |
| EPDS | 4,5,6,8 | 4 | 0.69 | 1.54 | 1.07 |
| EPDS | 4,5,6,9 | 4 | 0.68 | 1.60 | 1.11 |
| EPDS | 4,5,6,10 | 4 | 0.59 | 2.10 | 1.47 |
| EPDS | 4,5,7,8 | 4 | 0.70 | 1.55 | 1.17 |
| EPDS | 4,5,7,9 | 4 | 0.69 | 1.62 | 1.22 |
| EPDS | 4,5,7,10 | 4 | 0.62 | 2.06 | 1.54 |
| EPDS | 4,5,8,9 | 4 | 0.69 | 1.58 | 1.18 |
| EPDS | 4,5,8,10 | 4 | 0.70 | 1.79 | 1.34 |
| EPDS | 4,5,9,10 | 4 | 0.68 | 1.90 | 1.42 |
| EPDS | 4,6,7,8 | 4 | 0.72 | 1.49 | 1.07 |
| EPDS | 4,6,7,9 | 4 | 0.72 | 1.54 | 1.09 |
| EPDS | 4,6,7,10 | 4 | 0.69 | 1.84 | 1.34 |
| EPDS | 4,6,8,9 | 4 | 0.70 | 1.57 | 1.11 |
| EPDS | 4,6,8,10 | 4 | 0.73 | 1.71 | 1.21 |
| EPDS | 4,6,9,10 | 4 | 0.71 | 1.79 | 1.27 |
| EPDS | 4,7,8,9 | 4 | 0.70 | 1.60 | 1.22 |
| EPDS | 4,7,8,10 | 4 | 0.73 | 1.74 | 1.32 |

| | | | | | |
|---|---|---|---|---|---|
| EPDS | 4,7,9,10 | 4 | 0.71 | 1.83 | 1.38 |
| EPDS | 4,8,9,10 | 4 | 0.71 | 1.80 | 1.36 |
| EPDS | 5,6,7,8 | 4 | 0.71 | 1.56 | 1.20 |
| EPDS | 5,6,7,9 | 4 | 0.71 | 1.59 | 1.21 |
| EPDS | 5,6,7,10 | 4 | 0.68 | 1.92 | 1.45 |
| EPDS | 5,6,8,9 | 4 | 0.70 | 1.58 | 1.21 |
| EPDS | 5,6,8,10 | 4 | 0.73 | 1.75 | 1.33 |
| EPDS | 5,6,9,10 | 4 | 0.72 | 1.81 | 1.35 |
| EPDS | 5,7,8,9 | 4 | 0.68 | 1.69 | 1.35 |
| EPDS | 5,7,8,10 | 4 | 0.70 | 1.86 | 1.48 |
| EPDS | 5,7,9,10 | 4 | 0.69 | 1.93 | 1.53 |
| EPDS | 5,8,9,10 | 4 | 0.70 | 1.86 | 1.47 |
| EPDS | 6,7,8,9 | 4 | 0.61 | 1.90 | 1.48 |
| EPDS | 6,7,8,10 | 4 | 0.66 | 2.03 | 1.59 |
| EPDS | 6,7,9,10 | 4 | 0.65 | 2.08 | 1.63 |
| EPDS | 6,8,9,10 | 4 | 0.64 | 2.08 | 1.62 |
| EPDS | 7,8,9,10 | 4 | 0.59 | 2.26 | 1.80 |
| EPDS | 1,2,3,4,5 | 5 | 0.63 | 1.63 | 1.19 |
| EPDS | 1,2,3,4,6 | 5 | 0.67 | 1.52 | 1.11 |
| EPDS | 1,2,3,4,7 | 5 | 0.71 | 1.46 | 1.09 |
| EPDS | 1,2,3,4,8 | 5 | 0.73 | 1.37 | 1.01 |
| EPDS | 1,2,3,4,9 | 5 | 0.72 | 1.43 | 1.05 |
| EPDS | 1,2,3,4,10 | 5 | 0.66 | 1.84 | 1.37 |
| EPDS | 1,2,3,5,6 | 5 | 0.70 | 1.50 | 1.10 |
| EPDS | 1,2,3,5,7 | 5 | 0.70 | 1.52 | 1.19 |
| EPDS | 1,2,3,5,8 | 5 | 0.74 | 1.40 | 1.08 |
| EPDS | 1,2,3,5,9 | 5 | 0.73 | 1.44 | 1.11 |
| EPDS | 1,2,3,5,10 | 5 | 0.67 | 1.86 | 1.41 |
| EPDS | 1,2,3,6,7 | 5 | 0.67 | 1.64 | 1.26 |
| EPDS | 1,2,3,6,8 | 5 | 0.67 | 1.60 | 1.21 |
| EPDS | 1,2,3,6,9 | 5 | 0.67 | 1.63 | 1.24 |
| EPDS | 1,2,3,6,10 | 5 | 0.64 | 1.95 | 1.47 |
| EPDS | 1,2,3,7,8 | 5 | 0.66 | 1.65 | 1.30 |

| | | | | | |
|---|---|---|---|---|---|
| EPDS | 1,2,3,7,9 | 5 | 0.66 | 1.70 | 1.33 |
| EPDS | 1,2,3,7,10 | 5 | 0.64 | 2.00 | 1.57 |
| EPDS | 1,2,3,8,9 | 5 | 0.64 | 1.72 | 1.33 |
| EPDS | 1,2,3,8,10 | 5 | 0.67 | 1.88 | 1.46 |
| EPDS | 1,2,3,9,10 | 5 | 0.66 | 1.95 | 1.52 |
| EPDS | 1,2,4,5,6 | 5 | 0.65 | 1.58 | 1.16 |
| EPDS | 1,2,4,5,7 | 5 | 0.64 | 1.64 | 1.27 |
| EPDS | 1,2,4,5,8 | 5 | 0.69 | 1.47 | 1.13 |
| EPDS | 1,2,4,5,9 | 5 | 0.69 | 1.50 | 1.16 |
| EPDS | 1,2,4,5,10 | 5 | 0.61 | 1.98 | 1.51 |
| EPDS | 1,2,4,6,7 | 5 | 0.69 | 1.52 | 1.12 |
| EPDS | 1,2,4,6,8 | 5 | 0.70 | 1.46 | 1.07 |
| EPDS | 1,2,4,6,9 | 5 | 0.71 | 1.48 | 1.08 |
| EPDS | 1,2,4,6,10 | 5 | 0.67 | 1.82 | 1.35 |
| EPDS | 1,2,4,7,8 | 5 | 0.69 | 1.52 | 1.18 |
| EPDS | 1,2,4,7,9 | 5 | 0.69 | 1.56 | 1.20 |
| EPDS | 1,2,4,7,10 | 5 | 0.66 | 1.90 | 1.47 |
| EPDS | 1,2,4,8,9 | 5 | 0.68 | 1.56 | 1.21 |
| EPDS | 1,2,4,8,10 | 5 | 0.70 | 1.73 | 1.32 |
| EPDS | 1,2,4,9,10 | 5 | 0.70 | 1.79 | 1.38 |
| EPDS | 1,2,5,6,7 | 5 | 0.69 | 1.57 | 1.22 |
| EPDS | 1,2,5,6,8 | 5 | 0.71 | 1.48 | 1.16 |
| EPDS | 1,2,5,6,9 | 5 | 0.72 | 1.48 | 1.15 |
| EPDS | 1,2,5,6,10 | 5 | 0.68 | 1.83 | 1.41 |
| EPDS | 1,2,5,7,8 | 5 | 0.67 | 1.63 | 1.32 |
| EPDS | 1,2,5,7,9 | 5 | 0.68 | 1.64 | 1.34 |
| EPDS | 1,2,5,7,10 | 5 | 0.64 | 2.01 | 1.61 |
| EPDS | 1,2,5,8,9 | 5 | 0.68 | 1.61 | 1.30 |
| EPDS | 1,2,5,8,10 | 5 | 0.70 | 1.80 | 1.45 |
| EPDS | 1,2,5,9,10 | 5 | 0.70 | 1.84 | 1.48 |
| EPDS | 1,2,6,7,8 | 5 | 0.59 | 1.86 | 1.49 |
| EPDS | 1,2,6,7,9 | 5 | 0.60 | 1.87 | 1.50 |
| EPDS | 1,2,6,7,10 | 5 | 0.60 | 2.14 | 1.71 |

| | | | | | |
|---|---|---|---|---|---|
| EPDS | 1,2,6,8,9 | 5 | 0.58 | 1.89 | 1.50 |
| EPDS | 1,2,6,8,10 | 5 | 0.62 | 2.04 | 1.62 |
| EPDS | 1,2,6,9,10 | 5 | 0.62 | 2.07 | 1.65 |
| EPDS | 1,2,7,8,9 | 5 | 0.52 | 2.09 | 1.70 |
| EPDS | 1,2,7,8,10 | 5 | 0.56 | 2.26 | 1.85 |
| EPDS | 1,2,7,9,10 | 5 | 0.56 | 2.32 | 1.90 |
| EPDS | 1,2,8,9,10 | 5 | 0.56 | 2.27 | 1.84 |
| EPDS | 1,3,4,5,6 | 5 | 0.64 | 1.48 | 1.04 |
| EPDS | 1,3,4,5,7 | 5 | 0.68 | 1.41 | 1.04 |
| EPDS | 1,3,4,5,8 | 5 | 0.73 | 1.27 | 0.91 |
| EPDS | 1,3,4,5,9 | 5 | 0.71 | 1.34 | 0.97 |
| EPDS | 1,3,4,5,10 | 5 | 0.63 | 1.81 | 1.32 |
| EPDS | 1,3,4,6,7 | 5 | 0.73 | 1.30 | 0.94 |
| EPDS | 1,3,4,6,8 | 5 | 0.74 | 1.25 | 0.89 |
| EPDS | 1,3,4,6,9 | 5 | 0.73 | 1.30 | 0.94 |
| EPDS | 1,3,4,6,10 | 5 | 0.69 | 1.65 | 1.20 |
| EPDS | 1,3,4,7,8 | 5 | 0.76 | 1.22 | 0.90 |
| EPDS | 1,3,4,7,9 | 5 | 0.75 | 1.28 | 0.95 |
| EPDS | 1,3,4,7,10 | 5 | 0.72 | 1.59 | 1.18 |
| EPDS | 1,3,4,8,9 | 5 | 0.73 | 1.30 | 0.95 |
| EPDS | 1,3,4,8,10 | 5 | 0.75 | 1.45 | 1.07 |
| EPDS | 1,3,4,9,10 | 5 | 0.73 | 1.55 | 1.14 |
| EPDS | 1,3,5,6,7 | 5 | 0.73 | 1.34 | 0.99 |
| EPDS | 1,3,5,6,8 | 5 | 0.75 | 1.26 | 0.91 |
| EPDS | 1,3,5,6,9 | 5 | 0.75 | 1.29 | 0.94 |
| EPDS | 1,3,5,6,10 | 5 | 0.70 | 1.64 | 1.19 |
| EPDS | 1,3,5,7,8 | 5 | 0.74 | 1.30 | 1.01 |
| EPDS | 1,3,5,7,9 | 5 | 0.74 | 1.34 | 1.04 |
| EPDS | 1,3,5,7,10 | 5 | 0.71 | 1.67 | 1.28 |
| EPDS | 1,3,5,8,9 | 5 | 0.73 | 1.33 | 1.03 |
| EPDS | 1,3,5,8,10 | 5 | 0.75 | 1.50 | 1.15 |
| EPDS | 1,3,5,9,10 | 5 | 0.74 | 1.57 | 1.20 |
| EPDS | 1,3,6,7,8 | 5 | 0.68 | 1.47 | 1.12 |

| | | | | | |
|---|---|---|---|---|---|
| EPDS | 1,3,6,7,9 | 5 | 0.68 | 1.51 | 1.15 |
| EPDS | 1,3,6,7,10 | 5 | 0.69 | 1.74 | 1.33 |
| EPDS | 1,3,6,8,9 | 5 | 0.65 | 1.55 | 1.17 |
| EPDS | 1,3,6,8,10 | 5 | 0.70 | 1.67 | 1.26 |
| EPDS | 1,3,6,9,10 | 5 | 0.69 | 1.73 | 1.31 |
| EPDS | 1,3,7,8,9 | 5 | 0.64 | 1.61 | 1.27 |
| EPDS | 1,3,7,8,10 | 5 | 0.69 | 1.73 | 1.35 |
| EPDS | 1,3,7,9,10 | 5 | 0.68 | 1.81 | 1.41 |
| EPDS | 1,3,8,9,10 | 5 | 0.67 | 1.80 | 1.39 |
| EPDS | 1,4,5,6,7 | 5 | 0.70 | 1.37 | 1.00 |
| EPDS | 1,4,5,6,8 | 5 | 0.73 | 1.27 | 0.91 |
| EPDS | 1,4,5,6,9 | 5 | 0.73 | 1.29 | 0.93 |
| EPDS | 1,4,5,6,10 | 5 | 0.68 | 1.67 | 1.22 |
| EPDS | 1,4,5,7,8 | 5 | 0.71 | 1.33 | 1.03 |
| EPDS | 1,4,5,7,9 | 5 | 0.71 | 1.37 | 1.06 |
| EPDS | 1,4,5,7,10 | 5 | 0.67 | 1.73 | 1.33 |
| EPDS | 1,4,5,8,9 | 5 | 0.71 | 1.35 | 1.04 |
| EPDS | 1,4,5,8,10 | 5 | 0.73 | 1.53 | 1.17 |
| EPDS | 1,4,5,9,10 | 5 | 0.72 | 1.59 | 1.22 |
| EPDS | 1,4,6,7,8 | 5 | 0.73 | 1.29 | 0.96 |
| EPDS | 1,4,6,7,9 | 5 | 0.73 | 1.32 | 0.97 |
| EPDS | 1,4,6,7,10 | 5 | 0.73 | 1.57 | 1.18 |
| EPDS | 1,4,6,8,9 | 5 | 0.71 | 1.36 | 0.99 |
| EPDS | 1,4,6,8,10 | 5 | 0.75 | 1.48 | 1.08 |
| EPDS | 1,4,6,9,10 | 5 | 0.74 | 1.53 | 1.11 |
| EPDS | 1,4,7,8,9 | 5 | 0.69 | 1.43 | 1.11 |
| EPDS | 1,4,7,8,10 | 5 | 0.73 | 1.56 | 1.20 |
| EPDS | 1,4,7,9,10 | 5 | 0.72 | 1.62 | 1.25 |
| EPDS | 1,4,8,9,10 | 5 | 0.72 | 1.60 | 1.23 |
| EPDS | 1,5,6,7,8 | 5 | 0.72 | 1.37 | 1.08 |
| EPDS | 1,5,6,7,9 | 5 | 0.73 | 1.38 | 1.09 |
| EPDS | 1,5,6,7,10 | 5 | 0.72 | 1.65 | 1.27 |
| EPDS | 1,5,6,8,9 | 5 | 0.72 | 1.38 | 1.08 |

| | | | | | |
|---|---|---|---|---|---|
| EPDS | 1,5,6,8,10 | 5 | 0.74 | 1.52 | 1.18 |
| EPDS | 1,5,6,9,10 | 5 | 0.75 | 1.55 | 1.20 |
| EPDS | 1,5,7,8,9 | 5 | 0.68 | 1.53 | 1.24 |
| EPDS | 1,5,7,8,10 | 5 | 0.71 | 1.68 | 1.36 |
| EPDS | 1,5,7,9,10 | 5 | 0.70 | 1.72 | 1.39 |
| EPDS | 1,5,8,9,10 | 5 | 0.71 | 1.67 | 1.34 |
| EPDS | 1,6,7,8,9 | 5 | 0.58 | 1.77 | 1.41 |
| EPDS | 1,6,7,8,10 | 5 | 0.64 | 1.88 | 1.50 |
| EPDS | 1,6,7,9,10 | 5 | 0.64 | 1.92 | 1.53 |
| EPDS | 1,6,8,9,10 | 5 | 0.63 | 1.93 | 1.53 |
| EPDS | 1,7,8,9,10 | 5 | 0.57 | 2.13 | 1.73 |
| EPDS | 2,3,4,5,6 | 5 | 0.65 | 1.42 | 1.00 |
| EPDS | 2,3,4,5,7 | 5 | 0.69 | 1.34 | 1.00 |
| EPDS | 2,3,4,5,8 | 5 | 0.73 | 1.23 | 0.89 |
| EPDS | 2,3,4,5,9 | 5 | 0.73 | 1.27 | 0.93 |
| EPDS | 2,3,4,5,10 | 5 | 0.65 | 1.71 | 1.26 |
| EPDS | 2,3,4,6,7 | 5 | 0.73 | 1.25 | 0.90 |
| EPDS | 2,3,4,6,8 | 5 | 0.73 | 1.23 | 0.87 |
| EPDS | 2,3,4,6,9 | 5 | 0.73 | 1.26 | 0.90 |
| EPDS | 2,3,4,6,10 | 5 | 0.70 | 1.59 | 1.15 |
| EPDS | 2,3,4,7,8 | 5 | 0.76 | 1.19 | 0.88 |
| EPDS | 2,3,4,7,9 | 5 | 0.75 | 1.23 | 0.92 |
| EPDS | 2,3,4,7,10 | 5 | 0.73 | 1.52 | 1.14 |
| EPDS | 2,3,4,8,9 | 5 | 0.73 | 1.27 | 0.93 |
| EPDS | 2,3,4,8,10 | 5 | 0.75 | 1.42 | 1.05 |
| EPDS | 2,3,4,9,10 | 5 | 0.74 | 1.49 | 1.10 |
| EPDS | 2,3,5,6,7 | 5 | 0.73 | 1.28 | 0.96 |
| EPDS | 2,3,5,6,8 | 5 | 0.74 | 1.23 | 0.90 |
| EPDS | 2,3,5,6,9 | 5 | 0.75 | 1.24 | 0.91 |
| EPDS | 2,3,5,6,10 | 5 | 0.71 | 1.58 | 1.16 |
| EPDS | 2,3,5,7,8 | 5 | 0.74 | 1.26 | 0.98 |
| EPDS | 2,3,5,7,9 | 5 | 0.75 | 1.28 | 1.00 |
| EPDS | 2,3,5,7,10 | 5 | 0.72 | 1.59 | 1.24 |

| | | | | | |
|---|---|---|---|---|---|
| EPDS | 2,3,5,8,9 | 5 | 0.73 | 1.29 | 1.00 |
| EPDS | 2,3,5,8,10 | 5 | 0.75 | 1.46 | 1.12 |
| EPDS | 2,3,5,9,10 | 5 | 0.75 | 1.50 | 1.16 |
| EPDS | 2,3,6,7,8 | 5 | 0.68 | 1.44 | 1.11 |
| EPDS | 2,3,6,7,9 | 5 | 0.69 | 1.46 | 1.12 |
| EPDS | 2,3,6,7,10 | 5 | 0.69 | 1.69 | 1.30 |
| EPDS | 2,3,6,8,9 | 5 | 0.65 | 1.52 | 1.15 |
| EPDS | 2,3,6,8,10 | 5 | 0.70 | 1.64 | 1.24 |
| EPDS | 2,3,6,9,10 | 5 | 0.70 | 1.68 | 1.28 |
| EPDS | 2,3,7,8,9 | 5 | 0.64 | 1.57 | 1.24 |
| EPDS | 2,3,7,8,10 | 5 | 0.69 | 1.69 | 1.32 |
| EPDS | 2,3,7,9,10 | 5 | 0.69 | 1.74 | 1.37 |
| EPDS | 2,3,8,9,10 | 5 | 0.67 | 1.75 | 1.36 |
| EPDS | 2,4,5,6,7 | 5 | 0.69 | 1.34 | 0.99 |
| EPDS | 2,4,5,6,8 | 5 | 0.72 | 1.26 | 0.91 |
| EPDS | 2,4,5,6,9 | 5 | 0.73 | 1.26 | 0.92 |
| EPDS | 2,4,5,6,10 | 5 | 0.68 | 1.63 | 1.20 |
| EPDS | 2,4,5,7,8 | 5 | 0.71 | 1.31 | 1.02 |
| EPDS | 2,4,5,7,9 | 5 | 0.71 | 1.33 | 1.04 |
| EPDS | 2,4,5,7,10 | 5 | 0.67 | 1.68 | 1.30 |
| EPDS | 2,4,5,8,9 | 5 | 0.70 | 1.32 | 1.02 |
| EPDS | 2,4,5,8,10 | 5 | 0.72 | 1.50 | 1.16 |
| EPDS | 2,4,5,9,10 | 5 | 0.72 | 1.55 | 1.19 |
| EPDS | 2,4,6,7,8 | 5 | 0.72 | 1.29 | 0.96 |
| EPDS | 2,4,6,7,9 | 5 | 0.73 | 1.30 | 0.96 |
| EPDS | 2,4,6,7,10 | 5 | 0.72 | 1.55 | 1.15 |
| EPDS | 2,4,6,8,9 | 5 | 0.70 | 1.35 | 0.99 |
| EPDS | 2,4,6,8,10 | 5 | 0.74 | 1.48 | 1.08 |
| EPDS | 2,4,6,9,10 | 5 | 0.74 | 1.50 | 1.10 |
| EPDS | 2,4,7,8,9 | 5 | 0.69 | 1.41 | 1.10 |
| EPDS | 2,4,7,8,10 | 5 | 0.73 | 1.54 | 1.19 |
| EPDS | 2,4,7,9,10 | 5 | 0.72 | 1.58 | 1.22 |
| EPDS | 2,4,8,9,10 | 5 | 0.71 | 1.58 | 1.22 |

| | | | | | |
|---|---|---|---|---|---|
| EPDS | 2,5,6,7,8 | 5 | 0.71 | 1.35 | 1.07 |
| EPDS | 2,5,6,7,9 | 5 | 0.73 | 1.35 | 1.07 |
| EPDS | 2,5,6,7,10 | 5 | 0.71 | 1.61 | 1.25 |
| EPDS | 2,5,6,8,9 | 5 | 0.71 | 1.36 | 1.07 |
| EPDS | 2,5,6,8,10 | 5 | 0.74 | 1.51 | 1.18 |
| EPDS | 2,5,6,9,10 | 5 | 0.75 | 1.52 | 1.18 |
| EPDS | 2,5,7,8,9 | 5 | 0.67 | 1.50 | 1.22 |
| EPDS | 2,5,7,8,10 | 5 | 0.70 | 1.65 | 1.34 |
| EPDS | 2,5,7,9,10 | 5 | 0.71 | 1.68 | 1.36 |
| EPDS | 2,5,8,9,10 | 5 | 0.71 | 1.64 | 1.33 |
| EPDS | 2,6,7,8,9 | 5 | 0.57 | 1.76 | 1.40 |
| EPDS | 2,6,7,8,10 | 5 | 0.63 | 1.87 | 1.50 |
| EPDS | 2,6,7,9,10 | 5 | 0.64 | 1.89 | 1.52 |
| EPDS | 2,6,8,9,10 | 5 | 0.62 | 1.91 | 1.51 |
| EPDS | 2,7,8,9,10 | 5 | 0.56 | 2.11 | 1.70 |
| EPDS | 3,4,5,6,7 | 5 | 0.59 | 1.45 | 0.99 |
| EPDS | 3,4,5,6,8 | 5 | 0.63 | 1.34 | 0.90 |
| EPDS | 3,4,5,6,9 | 5 | 0.62 | 1.41 | 0.96 |
| EPDS | 3,4,5,6,10 | 5 | 0.53 | 1.86 | 1.31 |
| EPDS | 3,4,5,7,8 | 5 | 0.68 | 1.27 | 0.90 |
| EPDS | 3,4,5,7,9 | 5 | 0.66 | 1.35 | 0.95 |
| EPDS | 3,4,5,7,10 | 5 | 0.61 | 1.73 | 1.25 |
| EPDS | 3,4,5,8,9 | 5 | 0.64 | 1.34 | 0.94 |
| EPDS | 3,4,5,8,10 | 5 | 0.68 | 1.52 | 1.08 |
| EPDS | 3,4,5,9,10 | 5 | 0.65 | 1.64 | 1.17 |
| EPDS | 3,4,6,7,8 | 5 | 0.70 | 1.22 | 0.85 |
| EPDS | 3,4,6,7,9 | 5 | 0.69 | 1.28 | 0.89 |
| EPDS | 3,4,6,7,10 | 5 | 0.67 | 1.56 | 1.11 |
| EPDS | 3,4,6,8,9 | 5 | 0.65 | 1.34 | 0.93 |
| EPDS | 3,4,6,8,10 | 5 | 0.70 | 1.47 | 1.04 |
| EPDS | 3,4,6,9,10 | 5 | 0.68 | 1.56 | 1.11 |
| EPDS | 3,4,7,8,9 | 5 | 0.68 | 1.29 | 0.92 |
| EPDS | 3,4,7,8,10 | 5 | 0.73 | 1.41 | 1.03 |

| | | | | | |
|---|---|---|---|---|---|
| EPDS | 3,4,7,9,10 | 5 | 0.71 | 1.50 | 1.09 |
| EPDS | 3,4,8,9,10 | 5 | 0.70 | 1.51 | 1.08 |
| EPDS | 3,5,6,7,8 | 5 | 0.69 | 1.28 | 0.91 |
| EPDS | 3,5,6,7,9 | 5 | 0.69 | 1.32 | 0.94 |
| EPDS | 3,5,6,7,10 | 5 | 0.67 | 1.61 | 1.16 |
| EPDS | 3,5,6,8,9 | 5 | 0.67 | 1.34 | 0.95 |
| EPDS | 3,5,6,8,10 | 5 | 0.71 | 1.49 | 1.06 |
| EPDS | 3,5,6,9,10 | 5 | 0.69 | 1.56 | 1.10 |
| EPDS | 3,5,7,8,9 | 5 | 0.67 | 1.36 | 1.03 |
| EPDS | 3,5,7,8,10 | 5 | 0.71 | 1.50 | 1.14 |
| EPDS | 3,5,7,9,10 | 5 | 0.70 | 1.58 | 1.19 |
| EPDS | 3,5,8,9,10 | 5 | 0.70 | 1.55 | 1.17 |
| EPDS | 3,6,7,8,9 | 5 | 0.61 | 1.53 | 1.14 |
| EPDS | 3,6,7,8,10 | 5 | 0.67 | 1.62 | 1.22 |
| EPDS | 3,6,7,9,10 | 5 | 0.66 | 1.69 | 1.26 |
| EPDS | 3,6,8,9,10 | 5 | 0.64 | 1.72 | 1.27 |
| EPDS | 3,7,8,9,10 | 5 | 0.64 | 1.76 | 1.36 |
| EPDS | 4,5,6,7,8 | 5 | 0.68 | 1.25 | 0.89 |
| EPDS | 4,5,6,7,9 | 5 | 0.68 | 1.29 | 0.91 |
| EPDS | 4,5,6,7,10 | 5 | 0.66 | 1.59 | 1.14 |
| EPDS | 4,5,6,8,9 | 5 | 0.66 | 1.30 | 0.91 |
| EPDS | 4,5,6,8,10 | 5 | 0.70 | 1.45 | 1.03 |
| EPDS | 4,5,6,9,10 | 5 | 0.69 | 1.52 | 1.07 |
| EPDS | 4,5,7,8,9 | 5 | 0.66 | 1.34 | 1.02 |
| EPDS | 4,5,7,8,10 | 5 | 0.70 | 1.48 | 1.14 |
| EPDS | 4,5,7,9,10 | 5 | 0.69 | 1.56 | 1.18 |
| EPDS | 4,5,8,9,10 | 5 | 0.69 | 1.52 | 1.15 |
| EPDS | 4,6,7,8,9 | 5 | 0.67 | 1.32 | 0.95 |
| EPDS | 4,6,7,8,10 | 5 | 0.73 | 1.42 | 1.03 |
| EPDS | 4,6,7,9,10 | 5 | 0.72 | 1.47 | 1.06 |
| EPDS | 4,6,8,9,10 | 5 | 0.70 | 1.49 | 1.08 |
| EPDS | 4,7,8,9,10 | 5 | 0.70 | 1.55 | 1.19 |
| EPDS | 5,6,7,8,9 | 5 | 0.67 | 1.38 | 1.07 |

| | | | | | |
|---|---|---|---|---|---|
| EPDS | 5,6,7,8,10 | 5 | 0.71 | 1.50 | 1.17 |
| EPDS | 5,6,7,9,10 | 5 | 0.71 | 1.54 | 1.19 |
| EPDS | 5,6,8,9,10 | 5 | 0.70 | 1.53 | 1.18 |
| EPDS | 5,7,8,9,10 | 5 | 0.67 | 1.65 | 1.32 |
| EPDS | 6,7,8,9,10 | 5 | 0.60 | 1.86 | 1.46 |
| PHQ-9 | 1,2 | 2 | 0.61 | 3.40 | 2.62 |
| PHQ-9 | 1,3 | 2 | 0.64 | 3.27 | 2.43 |
| PHQ-9 | 1,4 | 2 | 0.63 | 3.29 | 2.41 |
| PHQ-9 | 1,5 | 2 | 0.65 | 3.27 | 2.50 |
| PHQ-9 | 1,6 | 2 | 0.67 | 3.18 | 2.48 |
| PHQ-9 | 1,7 | 2 | 0.66 | 3.20 | 2.46 |
| PHQ-9 | 1,8 | 2 | 0.63 | 3.49 | 2.72 |
| PHQ-9 | 1,9 | 2 | 0.59 | 3.87 | 3.01 |
| PHQ-9 | 2,3 | 2 | 0.68 | 3.09 | 2.31 |
| PHQ-9 | 2,4 | 2 | 0.69 | 3.03 | 2.22 |
| PHQ-9 | 2,5 | 2 | 0.69 | 3.06 | 2.36 |
| PHQ-9 | 2,6 | 2 | 0.63 | 3.36 | 2.64 |
| PHQ-9 | 2,7 | 2 | 0.70 | 3.02 | 2.35 |
| PHQ-9 | 2,8 | 2 | 0.67 | 3.28 | 2.58 |
| PHQ-9 | 2,9 | 2 | 0.60 | 3.77 | 2.98 |
| PHQ-9 | 3,4 | 2 | 0.53 | 3.75 | 2.77 |
| PHQ-9 | 3,5 | 2 | 0.56 | 3.66 | 2.80 |
| PHQ-9 | 3,6 | 2 | 0.66 | 3.20 | 2.46 |
| PHQ-9 | 3,7 | 2 | 0.61 | 3.46 | 2.66 |
| PHQ-9 | 3,8 | 2 | 0.55 | 3.85 | 3.00 |
| PHQ-9 | 3,9 | 2 | 0.53 | 4.14 | 3.24 |
| PHQ-9 | 4,5 | 2 | 0.59 | 3.53 | 2.64 |
| PHQ-9 | 4,6 | 2 | 0.69 | 3.03 | 2.27 |
| PHQ-9 | 4,7 | 2 | 0.64 | 3.28 | 2.45 |
| PHQ-9 | 4,8 | 2 | 0.62 | 3.54 | 2.68 |
| PHQ-9 | 4,9 | 2 | 0.60 | 3.80 | 2.90 |
| PHQ-9 | 5,6 | 2 | 0.63 | 3.37 | 2.65 |
| PHQ-9 | 5,7 | 2 | 0.61 | 3.45 | 2.70 |

| | | | | | |
|---|---|---|---|---|---|
| PHQ-9 | 5,8 | 2 | 0.55 | 3.90 | 3.08 |
| PHQ-9 | 5,9 | 2 | 0.53 | 4.17 | 3.33 |
| PHQ-9 | 6,7 | 2 | 0.62 | 3.40 | 2.70 |
| PHQ-9 | 6,8 | 2 | 0.59 | 3.71 | 2.98 |
| PHQ-9 | 6,9 | 2 | 0.52 | 4.24 | 3.40 |
| PHQ-9 | 7,8 | 2 | 0.51 | 4.09 | 3.27 |
| PHQ-9 | 7,9 | 2 | 0.53 | 4.15 | 3.33 |
| PHQ-9 | 8,9 | 2 | 0.40 | 4.90 | 4.05 |
| PHQ-9 | 1,2,3 | 3 | 0.67 | 2.65 | 1.96 |
| PHQ-9 | 1,2,4 | 3 | 0.65 | 2.70 | 1.96 |
| PHQ-9 | 1,2,5 | 3 | 0.67 | 2.65 | 2.03 |
| PHQ-9 | 1,2,6 | 3 | 0.62 | 2.85 | 2.23 |
| PHQ-9 | 1,2,7 | 3 | 0.68 | 2.60 | 2.02 |
| PHQ-9 | 1,2,8 | 3 | 0.67 | 2.78 | 2.17 |
| PHQ-9 | 1,2,9 | 3 | 0.62 | 3.18 | 2.47 |
| PHQ-9 | 1,3,4 | 3 | 0.62 | 2.83 | 2.00 |
| PHQ-9 | 1,3,5 | 3 | 0.68 | 2.64 | 1.94 |
| PHQ-9 | 1,3,6 | 3 | 0.73 | 2.39 | 1.79 |
| PHQ-9 | 1,3,7 | 3 | 0.71 | 2.51 | 1.85 |
| PHQ-9 | 1,3,8 | 3 | 0.70 | 2.67 | 2.00 |
| PHQ-9 | 1,3,9 | 3 | 0.68 | 2.90 | 2.18 |
| PHQ-9 | 1,4,5 | 3 | 0.66 | 2.71 | 1.97 |
| PHQ-9 | 1,4,6 | 3 | 0.72 | 2.42 | 1.79 |
| PHQ-9 | 1,4,7 | 3 | 0.70 | 2.54 | 1.85 |
| PHQ-9 | 1,4,8 | 3 | 0.70 | 2.66 | 1.96 |
| PHQ-9 | 1,4,9 | 3 | 0.68 | 2.89 | 2.14 |
| PHQ-9 | 1,5,6 | 3 | 0.71 | 2.53 | 1.96 |
| PHQ-9 | 1,5,7 | 3 | 0.71 | 2.52 | 1.93 |
| PHQ-9 | 1,5,8 | 3 | 0.69 | 2.74 | 2.11 |
| PHQ-9 | 1,5,9 | 3 | 0.68 | 2.95 | 2.28 |
| PHQ-9 | 1,6,7 | 3 | 0.70 | 2.55 | 2.00 |
| PHQ-9 | 1,6,8 | 3 | 0.70 | 2.69 | 2.11 |
| PHQ-9 | 1,6,9 | 3 | 0.65 | 3.07 | 2.40 |

| | | | | | |
|---|---|---|---|---|---|
| PHQ-9 | 1,7,8 | 3 | 0.66 | 2.86 | 2.22 |
| PHQ-9 | 1,7,9 | 3 | 0.68 | 2.93 | 2.28 |
| PHQ-9 | 1,8,9 | 3 | 0.64 | 3.25 | 2.56 |
| PHQ-9 | 2,3,4 | 3 | 0.68 | 2.61 | 1.86 |
| PHQ-9 | 2,3,5 | 3 | 0.72 | 2.46 | 1.82 |
| PHQ-9 | 2,3,6 | 3 | 0.70 | 2.53 | 1.90 |
| PHQ-9 | 2,3,7 | 3 | 0.74 | 2.35 | 1.75 |
| PHQ-9 | 2,3,8 | 3 | 0.73 | 2.50 | 1.88 |
| PHQ-9 | 2,3,9 | 3 | 0.69 | 2.84 | 2.15 |
| PHQ-9 | 2,4,5 | 3 | 0.71 | 2.48 | 1.81 |
| PHQ-9 | 2,4,6 | 3 | 0.71 | 2.48 | 1.84 |
| PHQ-9 | 2,4,7 | 3 | 0.74 | 2.33 | 1.70 |
| PHQ-9 | 2,4,8 | 3 | 0.75 | 2.42 | 1.79 |
| PHQ-9 | 2,4,9 | 3 | 0.71 | 2.76 | 2.05 |
| PHQ-9 | 2,5,6 | 3 | 0.68 | 2.63 | 2.05 |
| PHQ-9 | 2,5,7 | 3 | 0.74 | 2.35 | 1.81 |
| PHQ-9 | 2,5,8 | 3 | 0.73 | 2.54 | 1.98 |
| PHQ-9 | 2,5,9 | 3 | 0.69 | 2.86 | 2.22 |
| PHQ-9 | 2,6,7 | 3 | 0.68 | 2.65 | 2.08 |
| PHQ-9 | 2,6,8 | 3 | 0.67 | 2.82 | 2.23 |
| PHQ-9 | 2,6,9 | 3 | 0.61 | 3.27 | 2.58 |
| PHQ-9 | 2,7,8 | 3 | 0.70 | 2.68 | 2.10 |
| PHQ-9 | 2,7,9 | 3 | 0.70 | 2.86 | 2.24 |
| PHQ-9 | 2,8,9 | 3 | 0.66 | 3.15 | 2.49 |
| PHQ-9 | 3,4,5 | 3 | 0.56 | 3.10 | 2.25 |
| PHQ-9 | 3,4,6 | 3 | 0.70 | 2.56 | 1.86 |
| PHQ-9 | 3,4,7 | 3 | 0.63 | 2.84 | 2.06 |
| PHQ-9 | 3,4,8 | 3 | 0.62 | 3.03 | 2.24 |
| PHQ-9 | 3,4,9 | 3 | 0.62 | 3.20 | 2.38 |
| PHQ-9 | 3,5,6 | 3 | 0.69 | 2.60 | 1.97 |
| PHQ-9 | 3,5,7 | 3 | 0.65 | 2.77 | 2.09 |
| PHQ-9 | 3,5,8 | 3 | 0.62 | 3.05 | 2.34 |
| PHQ-9 | 3,5,9 | 3 | 0.63 | 3.19 | 2.46 |

| | | | | | |
|---|---|---|---|---|---|
| PHQ-9 | 3,6,7 | 3 | 0.70 | 2.55 | 1.95 |
| PHQ-9 | 3,6,8 | 3 | 0.70 | 2.71 | 2.09 |
| PHQ-9 | 3,6,9 | 3 | 0.66 | 3.04 | 2.35 |
| PHQ-9 | 3,7,8 | 3 | 0.61 | 3.10 | 2.39 |
| PHQ-9 | 3,7,9 | 3 | 0.65 | 3.08 | 2.38 |
| PHQ-9 | 3,8,9 | 3 | 0.59 | 3.48 | 2.73 |
| PHQ-9 | 4,5,6 | 3 | 0.70 | 2.54 | 1.90 |
| PHQ-9 | 4,5,7 | 3 | 0.66 | 2.70 | 2.01 |
| PHQ-9 | 4,5,8 | 3 | 0.65 | 2.91 | 2.19 |
| PHQ-9 | 4,5,9 | 3 | 0.65 | 3.05 | 2.30 |
| PHQ-9 | 4,6,7 | 3 | 0.73 | 2.44 | 1.83 |
| PHQ-9 | 4,6,8 | 3 | 0.73 | 2.53 | 1.91 |
| PHQ-9 | 4,6,9 | 3 | 0.70 | 2.85 | 2.15 |
| PHQ-9 | 4,7,8 | 3 | 0.65 | 2.90 | 2.19 |
| PHQ-9 | 4,7,9 | 3 | 0.69 | 2.88 | 2.17 |
| PHQ-9 | 4,8,9 | 3 | 0.66 | 3.16 | 2.42 |
| PHQ-9 | 5,6,7 | 3 | 0.68 | 2.69 | 2.11 |
| PHQ-9 | 5,6,8 | 3 | 0.66 | 2.91 | 2.31 |
| PHQ-9 | 5,6,9 | 3 | 0.62 | 3.22 | 2.55 |
| PHQ-9 | 5,7,8 | 3 | 0.61 | 3.14 | 2.46 |
| PHQ-9 | 5,7,9 | 3 | 0.65 | 3.12 | 2.46 |
| PHQ-9 | 5,8,9 | 3 | 0.58 | 3.57 | 2.86 |
| PHQ-9 | 6,7,8 | 3 | 0.61 | 3.11 | 2.48 |
| PHQ-9 | 6,7,9 | 3 | 0.61 | 3.29 | 2.61 |
| PHQ-9 | 6,8,9 | 3 | 0.57 | 3.63 | 2.91 |
| PHQ-9 | 7,8,9 | 3 | 0.53 | 3.79 | 3.06 |
| PHQ-9 | 1,2,3,4 | 4 | 0.62 | 2.31 | 1.64 |
| PHQ-9 | 1,2,3,5 | 4 | 0.68 | 2.12 | 1.54 |
| PHQ-9 | 1,2,3,6 | 4 | 0.68 | 2.15 | 1.60 |
| PHQ-9 | 1,2,3,7 | 4 | 0.71 | 2.02 | 1.48 |
| PHQ-9 | 1,2,3,8 | 4 | 0.72 | 2.12 | 1.58 |
| PHQ-9 | 1,2,3,9 | 4 | 0.68 | 2.42 | 1.81 |
| PHQ-9 | 1,2,4,5 | 4 | 0.65 | 2.21 | 1.59 |

| | | | | | |
|---|---|---|---|---|---|
| PHQ-9 | 1,2,4,6 | 4 | 0.66 | 2.19 | 1.61 |
| PHQ-9 | 1,2,4,7 | 4 | 0.69 | 2.07 | 1.50 |
| PHQ-9 | 1,2,4,8 | 4 | 0.71 | 2.14 | 1.57 |
| PHQ-9 | 1,2,4,9 | 4 | 0.67 | 2.44 | 1.80 |
| PHQ-9 | 1,2,5,6 | 4 | 0.65 | 2.25 | 1.74 |
| PHQ-9 | 1,2,5,7 | 4 | 0.71 | 2.03 | 1.56 |
| PHQ-9 | 1,2,5,8 | 4 | 0.71 | 2.17 | 1.68 |
| PHQ-9 | 1,2,5,9 | 4 | 0.68 | 2.45 | 1.90 |
| PHQ-9 | 1,2,6,7 | 4 | 0.65 | 2.26 | 1.78 |
| PHQ-9 | 1,2,6,8 | 4 | 0.66 | 2.37 | 1.88 |
| PHQ-9 | 1,2,6,9 | 4 | 0.60 | 2.76 | 2.16 |
| PHQ-9 | 1,2,7,8 | 4 | 0.68 | 2.30 | 1.80 |
| PHQ-9 | 1,2,7,9 | 4 | 0.68 | 2.44 | 1.91 |
| PHQ-9 | 1,2,8,9 | 4 | 0.66 | 2.65 | 2.08 |
| PHQ-9 | 1,3,4,5 | 4 | 0.62 | 2.36 | 1.66 |
| PHQ-9 | 1,3,4,6 | 4 | 0.72 | 2.01 | 1.41 |
| PHQ-9 | 1,3,4,7 | 4 | 0.67 | 2.17 | 1.52 |
| PHQ-9 | 1,3,4,8 | 4 | 0.68 | 2.26 | 1.60 |
| PHQ-9 | 1,3,4,9 | 4 | 0.68 | 2.43 | 1.74 |
| PHQ-9 | 1,3,5,6 | 4 | 0.74 | 1.94 | 1.44 |
| PHQ-9 | 1,3,5,7 | 4 | 0.72 | 2.02 | 1.46 |
| PHQ-9 | 1,3,5,8 | 4 | 0.71 | 2.16 | 1.60 |
| PHQ-9 | 1,3,5,9 | 4 | 0.72 | 2.30 | 1.72 |
| PHQ-9 | 1,3,6,7 | 4 | 0.75 | 1.89 | 1.42 |
| PHQ-9 | 1,3,6,8 | 4 | 0.76 | 1.96 | 1.49 |
| PHQ-9 | 1,3,6,9 | 4 | 0.73 | 2.25 | 1.70 |
| PHQ-9 | 1,3,7,8 | 4 | 0.71 | 2.20 | 1.64 |
| PHQ-9 | 1,3,7,9 | 4 | 0.74 | 2.22 | 1.67 |
| PHQ-9 | 1,3,8,9 | 4 | 0.72 | 2.42 | 1.83 |
| PHQ-9 | 1,4,5,6 | 4 | 0.72 | 2.01 | 1.48 |
| PHQ-9 | 1,4,5,7 | 4 | 0.70 | 2.09 | 1.51 |
| PHQ-9 | 1,4,5,8 | 4 | 0.70 | 2.20 | 1.61 |
| PHQ-9 | 1,4,5,9 | 4 | 0.70 | 2.35 | 1.74 |

| | | | | | |
|---|---|---|---|---|---|
| PHQ-9 | 1,4,6,7 | 4 | 0.74 | 1.93 | 1.43 |
| PHQ-9 | 1,4,6,8 | 4 | 0.76 | 1.97 | 1.48 |
| PHQ-9 | 1,4,6,9 | 4 | 0.72 | 2.27 | 1.69 |
| PHQ-9 | 1,4,7,8 | 4 | 0.70 | 2.21 | 1.63 |
| PHQ-9 | 1,4,7,9 | 4 | 0.73 | 2.23 | 1.65 |
| PHQ-9 | 1,4,8,9 | 4 | 0.72 | 2.39 | 1.79 |
| PHQ-9 | 1,5,6,7 | 4 | 0.72 | 2.02 | 1.57 |
| PHQ-9 | 1,5,6,8 | 4 | 0.73 | 2.14 | 1.68 |
| PHQ-9 | 1,5,6,9 | 4 | 0.70 | 2.41 | 1.88 |
| PHQ-9 | 1,5,7,8 | 4 | 0.70 | 2.26 | 1.75 |
| PHQ-9 | 1,5,7,9 | 4 | 0.73 | 2.28 | 1.77 |
| PHQ-9 | 1,5,8,9 | 4 | 0.70 | 2.52 | 1.97 |
| PHQ-9 | 1,6,7,8 | 4 | 0.69 | 2.29 | 1.81 |
| PHQ-9 | 1,6,7,9 | 4 | 0.69 | 2.45 | 1.93 |
| PHQ-9 | 1,6,8,9 | 4 | 0.68 | 2.62 | 2.06 |
| PHQ-9 | 1,7,8,9 | 4 | 0.67 | 2.66 | 2.08 |
| PHQ-9 | 2,3,4,5 | 4 | 0.67 | 2.15 | 1.53 |
| PHQ-9 | 2,3,4,6 | 4 | 0.69 | 2.08 | 1.47 |
| PHQ-9 | 2,3,4,7 | 4 | 0.72 | 1.98 | 1.40 |
| PHQ-9 | 2,3,4,8 | 4 | 0.73 | 2.05 | 1.45 |
| PHQ-9 | 2,3,4,9 | 4 | 0.70 | 2.33 | 1.67 |
| PHQ-9 | 2,3,5,6 | 4 | 0.71 | 2.03 | 1.51 |
| PHQ-9 | 2,3,5,7 | 4 | 0.76 | 1.85 | 1.35 |
| PHQ-9 | 2,3,5,8 | 4 | 0.76 | 1.98 | 1.48 |
| PHQ-9 | 2,3,5,9 | 4 | 0.73 | 2.23 | 1.67 |
| PHQ-9 | 2,3,6,7 | 4 | 0.72 | 1.99 | 1.49 |
| PHQ-9 | 2,3,6,8 | 4 | 0.73 | 2.08 | 1.58 |
| PHQ-9 | 2,3,6,9 | 4 | 0.69 | 2.42 | 1.84 |
| PHQ-9 | 2,3,7,8 | 4 | 0.74 | 2.05 | 1.54 |
| PHQ-9 | 2,3,7,9 | 4 | 0.75 | 2.16 | 1.63 |
| PHQ-9 | 2,3,8,9 | 4 | 0.73 | 2.35 | 1.78 |
| PHQ-9 | 2,4,5,6 | 4 | 0.71 | 2.05 | 1.50 |
| PHQ-9 | 2,4,5,7 | 4 | 0.75 | 1.88 | 1.36 |

| | | | | | |
|---|---|---|---|---|---|
| PHQ-9 | 2,4,5,8 | 4 | 0.75 | 1.97 | 1.45 |
| PHQ-9 | 2,4,5,9 | 4 | 0.73 | 2.23 | 1.65 |
| PHQ-9 | 2,4,6,7 | 4 | 0.73 | 1.96 | 1.45 |
| PHQ-9 | 2,4,6,8 | 4 | 0.75 | 2.00 | 1.50 |
| PHQ-9 | 2,4,6,9 | 4 | 0.70 | 2.35 | 1.76 |
| PHQ-9 | 2,4,7,8 | 4 | 0.75 | 2.00 | 1.48 |
| PHQ-9 | 2,4,7,9 | 4 | 0.76 | 2.11 | 1.57 |
| PHQ-9 | 2,4,8,9 | 4 | 0.75 | 2.25 | 1.68 |
| PHQ-9 | 2,5,6,7 | 4 | 0.71 | 2.07 | 1.62 |
| PHQ-9 | 2,5,6,8 | 4 | 0.71 | 2.21 | 1.74 |
| PHQ-9 | 2,5,6,9 | 4 | 0.67 | 2.53 | 1.98 |
| PHQ-9 | 2,5,7,8 | 4 | 0.74 | 2.08 | 1.62 |
| PHQ-9 | 2,5,7,9 | 4 | 0.74 | 2.19 | 1.70 |
| PHQ-9 | 2,5,8,9 | 4 | 0.72 | 2.41 | 1.90 |
| PHQ-9 | 2,6,7,8 | 4 | 0.67 | 2.37 | 1.88 |
| PHQ-9 | 2,6,7,9 | 4 | 0.66 | 2.58 | 2.03 |
| PHQ-9 | 2,6,8,9 | 4 | 0.64 | 2.77 | 2.19 |
| PHQ-9 | 2,7,8,9 | 4 | 0.69 | 2.57 | 2.02 |
| PHQ-9 | 3,4,5,6 | 4 | 0.68 | 2.16 | 1.56 |
| PHQ-9 | 3,4,5,7 | 4 | 0.62 | 2.37 | 1.70 |
| PHQ-9 | 3,4,5,8 | 4 | 0.61 | 2.54 | 1.86 |
| PHQ-9 | 3,4,5,9 | 4 | 0.64 | 2.63 | 1.93 |
| PHQ-9 | 3,4,6,7 | 4 | 0.71 | 2.04 | 1.48 |
| PHQ-9 | 3,4,6,8 | 4 | 0.73 | 2.10 | 1.54 |
| PHQ-9 | 3,4,6,9 | 4 | 0.70 | 2.37 | 1.74 |
| PHQ-9 | 3,4,7,8 | 4 | 0.63 | 2.50 | 1.83 |
| PHQ-9 | 3,4,7,9 | 4 | 0.68 | 2.44 | 1.80 |
| PHQ-9 | 3,4,8,9 | 4 | 0.66 | 2.67 | 2.00 |
| PHQ-9 | 3,5,6,7 | 4 | 0.71 | 2.07 | 1.57 |
| PHQ-9 | 3,5,6,8 | 4 | 0.71 | 2.20 | 1.69 |
| PHQ-9 | 3,5,6,9 | 4 | 0.69 | 2.44 | 1.87 |
| PHQ-9 | 3,5,7,8 | 4 | 0.64 | 2.49 | 1.89 |
| PHQ-9 | 3,5,7,9 | 4 | 0.69 | 2.43 | 1.86 |

| | | | | | |
|---|---|---|---|---|---|
| PHQ-9 | 3,5,8,9 | 4 | 0.65 | 2.74 | 2.13 |
| PHQ-9 | 3,6,7,8 | 4 | 0.69 | 2.29 | 1.77 |
| PHQ-9 | 3,6,7,9 | 4 | 0.70 | 2.41 | 1.86 |
| PHQ-9 | 3,6,8,9 | 4 | 0.68 | 2.61 | 2.02 |
| PHQ-9 | 3,7,8,9 | 4 | 0.63 | 2.81 | 2.19 |
| PHQ-9 | 4,5,6,7 | 4 | 0.72 | 2.03 | 1.52 |
| PHQ-9 | 4,5,6,8 | 4 | 0.73 | 2.12 | 1.60 |
| PHQ-9 | 4,5,6,9 | 4 | 0.71 | 2.36 | 1.78 |
| PHQ-9 | 4,5,7,8 | 4 | 0.66 | 2.40 | 1.80 |
| PHQ-9 | 4,5,7,9 | 4 | 0.71 | 2.35 | 1.77 |
| PHQ-9 | 4,5,8,9 | 4 | 0.68 | 2.58 | 1.97 |
| PHQ-9 | 4,6,7,8 | 4 | 0.72 | 2.16 | 1.64 |
| PHQ-9 | 4,6,7,9 | 4 | 0.73 | 2.28 | 1.73 |
| PHQ-9 | 4,6,8,9 | 4 | 0.73 | 2.41 | 1.83 |
| PHQ-9 | 4,7,8,9 | 4 | 0.68 | 2.59 | 1.98 |
| PHQ-9 | 5,6,7,8 | 4 | 0.65 | 2.46 | 1.94 |
| PHQ-9 | 5,6,7,9 | 4 | 0.67 | 2.57 | 2.02 |
| PHQ-9 | 5,6,8,9 | 4 | 0.64 | 2.82 | 2.24 |
| PHQ-9 | 5,7,8,9 | 4 | 0.62 | 2.89 | 2.30 |
| PHQ-9 | 6,7,8,9 | 4 | 0.59 | 3.03 | 2.41 |
| PHQ-9 | 1,2,3,4,5 | 5 | 0.59 | 1.89 | 1.33 |
| PHQ-9 | 1,2,3,4,6 | 5 | 0.62 | 1.80 | 1.28 |
| PHQ-9 | 1,2,3,4,7 | 5 | 0.64 | 1.73 | 1.22 |
| PHQ-9 | 1,2,3,4,8 | 5 | 0.68 | 1.77 | 1.25 |
| PHQ-9 | 1,2,3,4,9 | 5 | 0.65 | 2.03 | 1.44 |
| PHQ-9 | 1,2,3,5,6 | 5 | 0.66 | 1.70 | 1.25 |
| PHQ-9 | 1,2,3,5,7 | 5 | 0.71 | 1.56 | 1.13 |
| PHQ-9 | 1,2,3,5,8 | 5 | 0.72 | 1.66 | 1.23 |
| PHQ-9 | 1,2,3,5,9 | 5 | 0.70 | 1.89 | 1.40 |
| PHQ-9 | 1,2,3,6,7 | 5 | 0.68 | 1.65 | 1.23 |
| PHQ-9 | 1,2,3,6,8 | 5 | 0.71 | 1.71 | 1.30 |
| PHQ-9 | 1,2,3,6,9 | 5 | 0.66 | 2.03 | 1.53 |
| PHQ-9 | 1,2,3,7,8 | 5 | 0.71 | 1.71 | 1.28 |

| | | | | | |
|---|---|---|---|---|---|
| PHQ-9 | 1,2,3,7,9 | 5 | 0.72 | 1.82 | 1.36 |
| PHQ-9 | 1,2,3,8,9 | 5 | 0.71 | 1.96 | 1.48 |
| PHQ-9 | 1,2,4,5,6 | 5 | 0.63 | 1.79 | 1.31 |
| PHQ-9 | 1,2,4,5,7 | 5 | 0.68 | 1.66 | 1.19 |
| PHQ-9 | 1,2,4,5,8 | 5 | 0.70 | 1.72 | 1.26 |
| PHQ-9 | 1,2,4,5,9 | 5 | 0.67 | 1.96 | 1.44 |
| PHQ-9 | 1,2,4,6,7 | 5 | 0.66 | 1.71 | 1.26 |
| PHQ-9 | 1,2,4,6,8 | 5 | 0.70 | 1.73 | 1.29 |
| PHQ-9 | 1,2,4,6,9 | 5 | 0.65 | 2.06 | 1.54 |
| PHQ-9 | 1,2,4,7,8 | 5 | 0.69 | 1.74 | 1.28 |
| PHQ-9 | 1,2,4,7,9 | 5 | 0.71 | 1.85 | 1.37 |
| PHQ-9 | 1,2,4,8,9 | 5 | 0.71 | 1.96 | 1.46 |
| PHQ-9 | 1,2,5,6,7 | 5 | 0.66 | 1.75 | 1.37 |
| PHQ-9 | 1,2,5,6,8 | 5 | 0.67 | 1.85 | 1.46 |
| PHQ-9 | 1,2,5,6,9 | 5 | 0.63 | 2.15 | 1.68 |
| PHQ-9 | 1,2,5,7,8 | 5 | 0.70 | 1.76 | 1.37 |
| PHQ-9 | 1,2,5,7,9 | 5 | 0.71 | 1.86 | 1.45 |
| PHQ-9 | 1,2,5,8,9 | 5 | 0.70 | 2.04 | 1.60 |
| PHQ-9 | 1,2,6,7,8 | 5 | 0.62 | 2.00 | 1.59 |
| PHQ-9 | 1,2,6,7,9 | 5 | 0.62 | 2.19 | 1.73 |
| PHQ-9 | 1,2,6,8,9 | 5 | 0.62 | 2.32 | 1.84 |
| PHQ-9 | 1,2,7,8,9 | 5 | 0.66 | 2.18 | 1.72 |
| PHQ-9 | 1,3,4,5,6 | 5 | 0.69 | 1.66 | 1.15 |
| PHQ-9 | 1,3,4,5,7 | 5 | 0.64 | 1.78 | 1.23 |
| PHQ-9 | 1,3,4,5,8 | 5 | 0.66 | 1.87 | 1.31 |
| PHQ-9 | 1,3,4,5,9 | 5 | 0.67 | 1.99 | 1.41 |
| PHQ-9 | 1,3,4,6,7 | 5 | 0.72 | 1.55 | 1.09 |
| PHQ-9 | 1,3,4,6,8 | 5 | 0.75 | 1.56 | 1.12 |
| PHQ-9 | 1,3,4,6,9 | 5 | 0.72 | 1.83 | 1.31 |
| PHQ-9 | 1,3,4,7,8 | 5 | 0.66 | 1.85 | 1.30 |
| PHQ-9 | 1,3,4,7,9 | 5 | 0.71 | 1.84 | 1.30 |
| PHQ-9 | 1,3,4,8,9 | 5 | 0.71 | 1.97 | 1.43 |
| PHQ-9 | 1,3,5,6,7 | 5 | 0.75 | 1.48 | 1.09 |

| | | | | | |
|---|---|---|---|---|---|
| PHQ-9 | 1,3,5,6,8 | 5 | 0.76 | 1.55 | 1.17 |
| PHQ-9 | 1,3,5,6,9 | 5 | 0.74 | 1.79 | 1.34 |
| PHQ-9 | 1,3,5,7,8 | 5 | 0.71 | 1.75 | 1.29 |
| PHQ-9 | 1,3,5,7,9 | 5 | 0.75 | 1.73 | 1.28 |
| PHQ-9 | 1,3,5,8,9 | 5 | 0.74 | 1.91 | 1.44 |
| PHQ-9 | 1,3,6,7,8 | 5 | 0.74 | 1.63 | 1.24 |
| PHQ-9 | 1,3,6,7,9 | 5 | 0.74 | 1.76 | 1.34 |
| PHQ-9 | 1,3,6,8,9 | 5 | 0.75 | 1.87 | 1.42 |
| PHQ-9 | 1,3,7,8,9 | 5 | 0.72 | 1.97 | 1.50 |
| PHQ-9 | 1,4,5,6,7 | 5 | 0.72 | 1.56 | 1.15 |
| PHQ-9 | 1,4,5,6,8 | 5 | 0.75 | 1.60 | 1.20 |
| PHQ-9 | 1,4,5,6,9 | 5 | 0.72 | 1.85 | 1.38 |
| PHQ-9 | 1,4,5,7,8 | 5 | 0.69 | 1.80 | 1.32 |
| PHQ-9 | 1,4,5,7,9 | 5 | 0.73 | 1.79 | 1.31 |
| PHQ-9 | 1,4,5,8,9 | 5 | 0.73 | 1.94 | 1.45 |
| PHQ-9 | 1,4,6,7,8 | 5 | 0.73 | 1.65 | 1.24 |
| PHQ-9 | 1,4,6,7,9 | 5 | 0.74 | 1.78 | 1.34 |
| PHQ-9 | 1,4,6,8,9 | 5 | 0.75 | 1.86 | 1.40 |
| PHQ-9 | 1,4,7,8,9 | 5 | 0.72 | 1.96 | 1.48 |
| PHQ-9 | 1,5,6,7,8 | 5 | 0.70 | 1.79 | 1.41 |
| PHQ-9 | 1,5,6,7,9 | 5 | 0.71 | 1.91 | 1.49 |
| PHQ-9 | 1,5,6,8,9 | 5 | 0.71 | 2.06 | 1.62 |
| PHQ-9 | 1,5,7,8,9 | 5 | 0.70 | 2.06 | 1.62 |
| PHQ-9 | 1,6,7,8,9 | 5 | 0.66 | 2.22 | 1.75 |
| PHQ-9 | 2,3,4,5,6 | 5 | 0.66 | 1.71 | 1.20 |
| PHQ-9 | 2,3,4,5,7 | 5 | 0.71 | 1.59 | 1.10 |
| PHQ-9 | 2,3,4,5,8 | 5 | 0.72 | 1.66 | 1.15 |
| PHQ-9 | 2,3,4,5,9 | 5 | 0.70 | 1.88 | 1.33 |
| PHQ-9 | 2,3,4,6,7 | 5 | 0.70 | 1.60 | 1.13 |
| PHQ-9 | 2,3,4,6,8 | 5 | 0.73 | 1.61 | 1.15 |
| PHQ-9 | 2,3,4,6,9 | 5 | 0.68 | 1.94 | 1.39 |
| PHQ-9 | 2,3,4,7,8 | 5 | 0.72 | 1.65 | 1.16 |
| PHQ-9 | 2,3,4,7,9 | 5 | 0.74 | 1.74 | 1.23 |

| | | | | | |
|---|---|---|---|---|---|
| PHQ-9 | 2,3,4,8,9 | 5 | 0.74 | 1.85 | 1.33 |
| PHQ-9 | 2,3,5,6,7 | 5 | 0.73 | 1.54 | 1.14 |
| PHQ-9 | 2,3,5,6,8 | 5 | 0.74 | 1.63 | 1.23 |
| PHQ-9 | 2,3,5,6,9 | 5 | 0.70 | 1.91 | 1.44 |
| PHQ-9 | 2,3,5,7,8 | 5 | 0.75 | 1.58 | 1.17 |
| PHQ-9 | 2,3,5,7,9 | 5 | 0.77 | 1.65 | 1.23 |
| PHQ-9 | 2,3,5,8,9 | 5 | 0.76 | 1.82 | 1.38 |
| PHQ-9 | 2,3,6,7,8 | 5 | 0.71 | 1.72 | 1.31 |
| PHQ-9 | 2,3,6,7,9 | 5 | 0.71 | 1.88 | 1.43 |
| PHQ-9 | 2,3,6,8,9 | 5 | 0.71 | 2.01 | 1.53 |
| PHQ-9 | 2,3,7,8,9 | 5 | 0.74 | 1.90 | 1.44 |
| PHQ-9 | 2,4,5,6,7 | 5 | 0.71 | 1.57 | 1.16 |
| PHQ-9 | 2,4,5,6,8 | 5 | 0.74 | 1.62 | 1.21 |
| PHQ-9 | 2,4,5,6,9 | 5 | 0.70 | 1.91 | 1.42 |
| PHQ-9 | 2,4,5,7,8 | 5 | 0.75 | 1.59 | 1.17 |
| PHQ-9 | 2,4,5,7,9 | 5 | 0.76 | 1.66 | 1.23 |
| PHQ-9 | 2,4,5,8,9 | 5 | 0.76 | 1.80 | 1.34 |
| PHQ-9 | 2,4,6,7,8 | 5 | 0.72 | 1.67 | 1.25 |
| PHQ-9 | 2,4,6,7,9 | 5 | 0.72 | 1.84 | 1.38 |
| PHQ-9 | 2,4,6,8,9 | 5 | 0.73 | 1.92 | 1.44 |
| PHQ-9 | 2,4,7,8,9 | 5 | 0.75 | 1.83 | 1.37 |
| PHQ-9 | 2,5,6,7,8 | 5 | 0.68 | 1.84 | 1.45 |
| PHQ-9 | 2,5,6,7,9 | 5 | 0.69 | 1.99 | 1.56 |
| PHQ-9 | 2,5,6,8,9 | 5 | 0.68 | 2.15 | 1.69 |
| PHQ-9 | 2,5,7,8,9 | 5 | 0.73 | 1.96 | 1.54 |
| PHQ-9 | 2,6,7,8,9 | 5 | 0.63 | 2.33 | 1.84 |
| PHQ-9 | 3,4,5,6,7 | 5 | 0.68 | 1.69 | 1.21 |
| PHQ-9 | 3,4,5,6,8 | 5 | 0.70 | 1.75 | 1.28 |
| PHQ-9 | 3,4,5,6,9 | 5 | 0.69 | 1.97 | 1.44 |
| PHQ-9 | 3,4,5,7,8 | 5 | 0.60 | 2.08 | 1.51 |
| PHQ-9 | 3,4,5,7,9 | 5 | 0.68 | 2.00 | 1.47 |
| PHQ-9 | 3,4,5,8,9 | 5 | 0.66 | 2.20 | 1.65 |
| PHQ-9 | 3,4,6,7,8 | 5 | 0.70 | 1.77 | 1.30 |

| | | | | | |
|---|---|---|---|---|---|
| PHQ-9 | 3,4,6,7,9 | 5 | 0.71 | 1.87 | 1.37 |
| PHQ-9 | 3,4,6,8,9 | 5 | 0.72 | 1.97 | 1.46 |
| PHQ-9 | 3,4,7,8,9 | 5 | 0.66 | 2.18 | 1.63 |
| PHQ-9 | 3,5,6,7,8 | 5 | 0.69 | 1.84 | 1.41 |
| PHQ-9 | 3,5,6,7,9 | 5 | 0.71 | 1.92 | 1.47 |
| PHQ-9 | 3,5,6,8,9 | 5 | 0.70 | 2.09 | 1.61 |
| PHQ-9 | 3,5,7,8,9 | 5 | 0.66 | 2.22 | 1.72 |
| PHQ-9 | 3,6,7,8,9 | 5 | 0.67 | 2.19 | 1.69 |
| PHQ-9 | 4,5,6,7,8 | 5 | 0.70 | 1.78 | 1.35 |
| PHQ-9 | 4,5,6,7,9 | 5 | 0.72 | 1.87 | 1.41 |
| PHQ-9 | 4,5,6,8,9 | 5 | 0.72 | 1.99 | 1.51 |
| PHQ-9 | 4,5,7,8,9 | 5 | 0.69 | 2.12 | 1.62 |
| PHQ-9 | 4,6,7,8,9 | 5 | 0.71 | 2.04 | 1.55 |
| PHQ-9 | 5,6,7,8,9 | 5 | 0.63 | 2.37 | 1.87 |

Adding covariates did not improve EPDS model performance.

Covariates, such as demographics (i.e., age and race/ethnicity), pregnancy outcomes, and prior history of mental disorders, are known to increase risk for PPD[35]. To investigate whether these covariates impacted ML model performance for EPDS total score prediction using a subset of questions, we built ML models using each permutation of 2-5 EPDS questions in our N3C postpartum cohort adding covariates to models to assess their impact on model performance: 1) no covariates, 2) demographics (i.e., age and race/ethnicity), 3) pregnancy outcome (i.e., live birth, stillbirth, spontaneous abortion, induced abortion, ectopic pregnancy, delivery record only, and missing pregnancy outcome), 4) prior mental health history (i.e., any mental health diagnosis prior to the EPDS assessment date), and 5) all 3 (i.e., demographics, pregnancy outcome, and prior mental health history). The results revealed models that included covariates did not perform better than those without covariates *($P>0.05$)* (Figure S7).

**Figure S7: Incorporating covariates in the model did not improve its performance.**

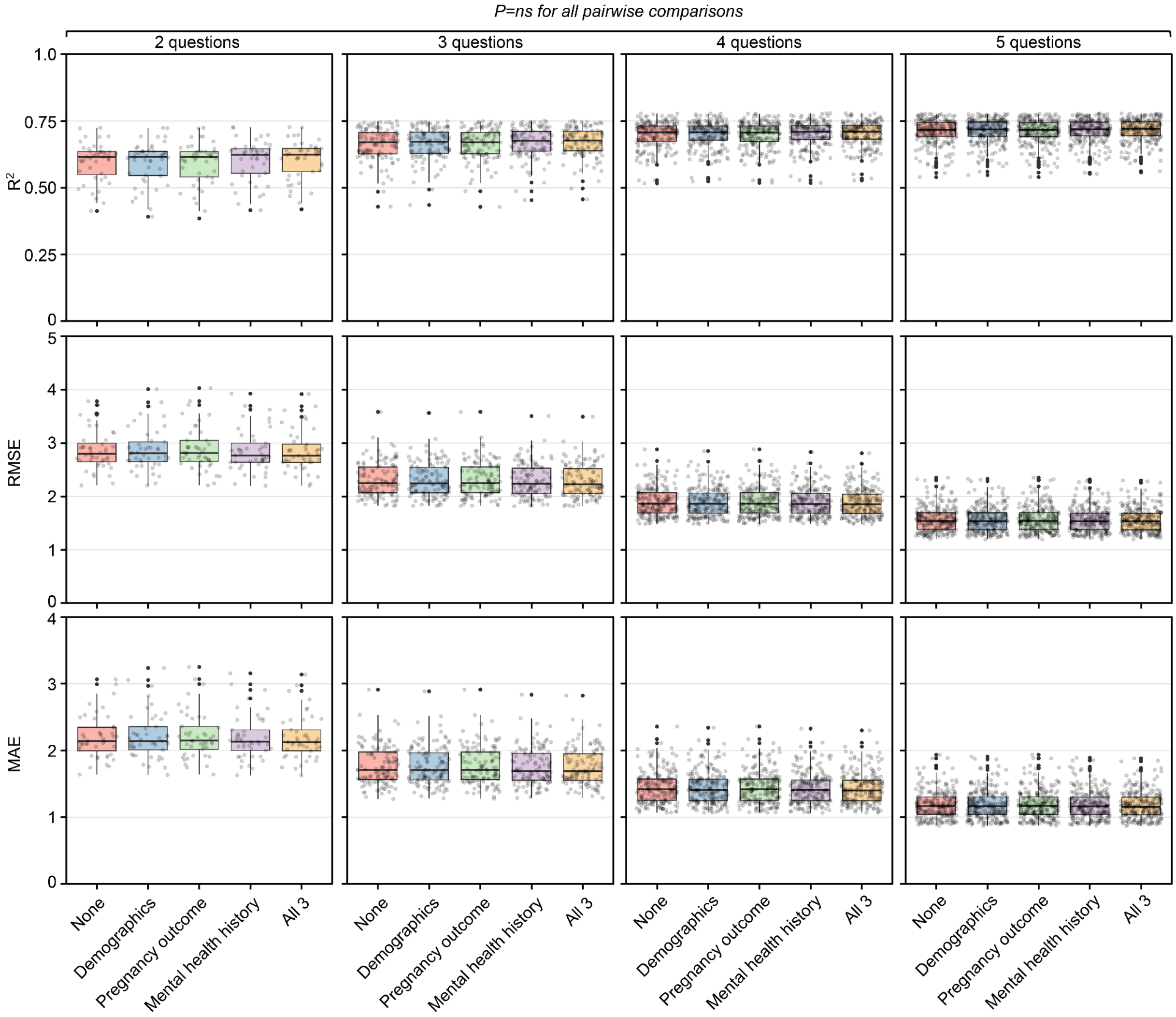


ns = not significant

*Box plots showing model performance metrics (R², RMSE, MAE) across all 2, 3, 4, and 5-question EPDS combinations in the N3C postpartum cohort. Models were evaluated with: no covariates (None), demographics only, pregnancy outcomes only, mental health history only, and all covariates combined (All 3). Each gray dot represents an individual n-question model. Results demonstrate that predictive performance was statistically equivalent across all covariate combinations, with no significant pairwise differences observed (all P=ns).*

**Figure S8: Our ML approach exhibited the best performance.**

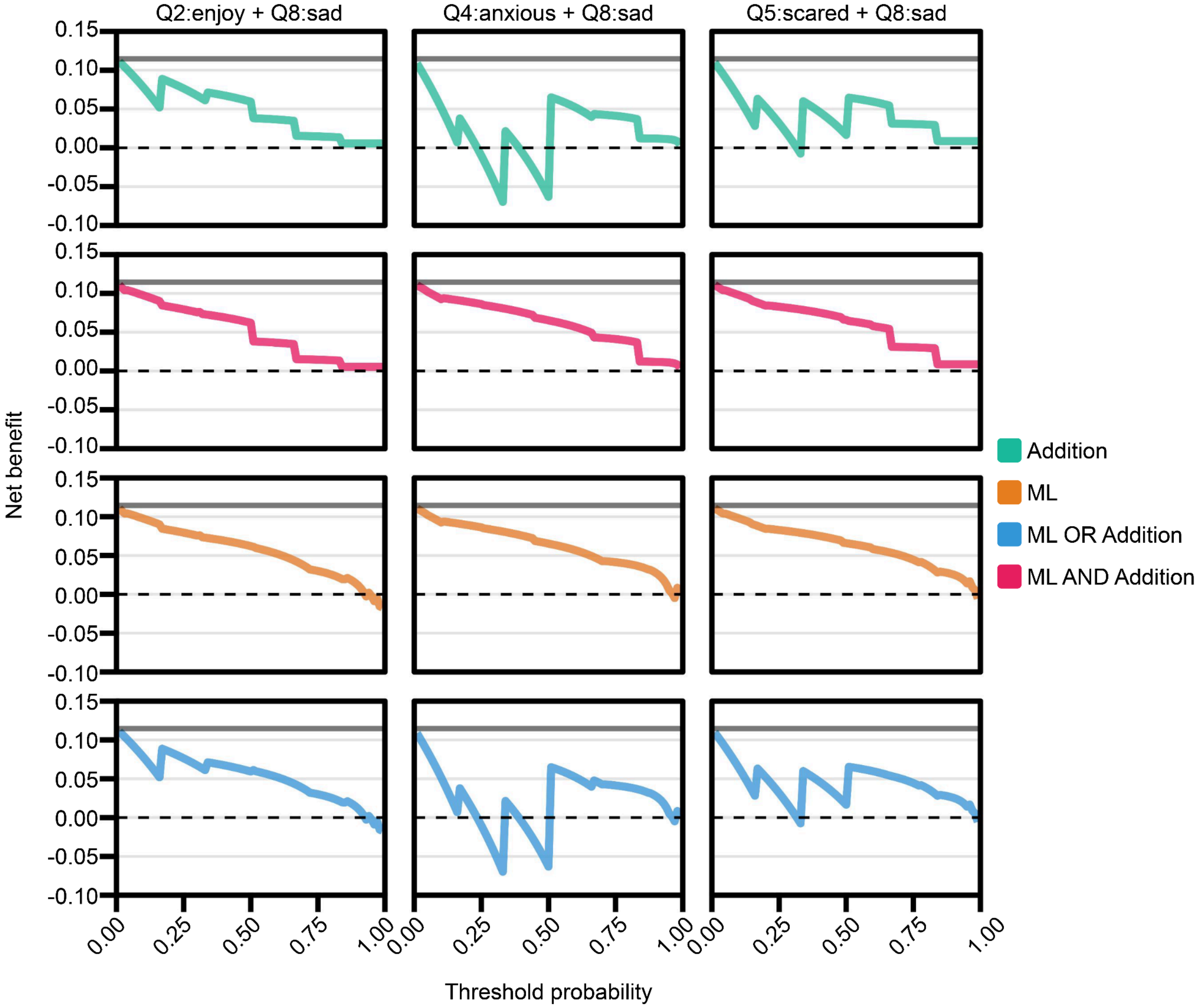


*Decision curve analysis evaluating clinical utility of different EPDS question combinations and screening approaches. Our ML method demonstrated superior net benefit over traditional approaches, with the greatest advantage observed at lower threshold probabilities.*
